# Phase 3, multicentre, double-blind, randomised, parallel-group, placebo-controlled study of camostat mesilate (FOY-305) for the treatment of COVID-19 (CANDLE study)

**DOI:** 10.1101/2022.03.27.22271988

**Authors:** Taku Kinoshita, Masahiro Shinoda, Yasuhiro Nishizaki, Katsuya Shiraki, Yuji Hirai, Yoshiko Kichikawa, Kenji Tsushima, Masaharu Sinkai, Naoyuki Komura, Kazuo Yoshida, Yasutoshi Kido, Hiroshi Kakeya, Naoto Uemura, Junichi Kadota

**Affiliations:** Respiratory Medicine, International University of Health and Welfare Narita Hospital, Narita, Japan; Department of Respiratory Medicine, Tokyo Shinagawa Hospital, Shinagawa, Tokyo, Japan; Tokai University Tokyo Hospital, Tokyo, Japan; Department of General and Laboratory Medicine, Mie Prefectural General Medical Center, Yokkaichi, Japan; Department of Infectious Diseases, Tokyo Medical University Hachioji Medical Center, Hachioji, Japan; Department of Respiratory Medicine, Mishuku Hospital, Tokyo, Japan; Department of Statistical Analysis, Ono Pharmaceutical Co., Ltd., Osaka, Japan; Clinical Development Planning, Ono Pharmaceutical Co., Ltd., Osaka, Japan; Department of Parasitology and Research Center for Infectious Disease Sciences, Graduate School of Medicine, Osaka City University, Osaka, Japan; Department of Infection Control Science, Graduate School of Medicine, Osaka City University, Osaka, Japan; Department of Clinical Pharmacology and Therapeutics and Department of Respiratory Medicine and Infectious Diseases, Faculty of Medicine, Oita University, Oita, Japan; Nagasaki Harbor Medical Center, Nagasaki, Japan

**Author notes:** **Corresponding author** Naoto Uemura, Department of Clinical Pharmacology and Therapeutics, Faculty of Medicine, Oita University, 1-1 Idaigaoka, Hasama-machi, Yufu-shi 879-5593, Japan. **Prior publication** None.

**Keywords:** camostat mesilate, COVID-19, randomised controlled trial, SARS-CoV-2

## Abstract

**Background:** *In vitro* drug-screening studies have indicated that camostat mesilate (FOY-305) may prevent SARS-CoV-2 infection into human airway epithelial cells. This study was conducted to investigate whether camostat mesilate is an effective treatment for SARS-CoV-2 infection (COVID-19).

**Methods:** This was a phase 3, multicentre, double-blind, randomised, parallel-group, placebo-controlled study. Patients were enrolled if they were admitted to a hospital within 5 days of onset of COVID-19 symptoms or within 5 days of a positive test for asymptomatic patients. Severe cases (e.g., those requiring oxygenation/ventilation) were excluded. Patients were administered camostat mesilate (600 mg qid; four to eight times higher than the clinical doses in Japan) or placebo for up to 14 days. The primary efficacy endpoint was the time to the first two consecutive negative tests for SARS-CoV-2.

**Findings:** One-hundred and fifty-five patients were randomised to receive camostat mesilate (n=78) or placebo (n=77). The median time to the first test was 11 days in both groups, and conversion to negative status was observed in 60·8% and 63·5% of patients in the camostat mesilate and placebo groups, respectively. The primary (Bayesian) and secondary (frequentist) analyses found no significant differences in the primary endpoint between the two groups. No additional safety concerns beyond those already known for camostat mesilate were identified.

**Interpretation:** Camostat mesilate is no more effective, based on upper airway viral clearance, than placebo for treating patients with mild to moderate SARS-CoV-2 infection with or without symptoms.

**Funding:** Ono Pharmaceutical Co., Ltd.

**RESEARCH IN CONTEXT PANEL:** *Evidence before this study:* SARS-CoV-2 infection (COVID-19), as a significant global health threat, is characterised by broad symptoms and varying disease severity. At the time of planning this study, there were no specific treatments for COVID-19 beyond the use of antiviral drugs, steroids and, in severe cases, ventilation with oxygen. Pre-clinical screening studies revealed the spike (S) protein of SARS-CoV-2 bind to angiotensin converting enzyme II (ACE2) on the host cell membrane. The S protein is then cleaved by a type II transmembrane serine protease (TMPRSS2) as an essential enzyme for the viral entry into host cells. *In vitro* drug-screening studies have shown that drugs that block binding of the S protein to ACE2 can prevent viral entry into a cell line derived from human airway epithelium. The studies identified 4-(4-guanidinobenzoyloxy)phenylacetic acid, the active metabolite of a serine protease inhibitor (camostat mesilate, FOY-305), as a candidate inhibitor of SARS-CoV-2 entry into humans. A retrospective study of critically ill COVID-19 patients with organ failure revealed a decline in disease activity within 8 days of admission among patients treated with camostat mesilate. In consideration of the preclinical and early clinical evidence, it was hypothesised that camostat mesilate is an effective treatment for patients with COVID-19. Therefore, we planned and executed a phase 3, randomised, double-blind, placebo-controlled study to investigate the efficacy and safety of camostat mesilate for the treatment of patients with mild to moderate COVID-19 infection with or without symptoms. The primary endpoint was the time to the first two consecutive negative tests for SARS-CoV-2. No controlled clinical studies of camostat mesilate had been conducted at the time of planning this study.

*Added value of this study:* The results of this randomised controlled trial revealed that camostat mesilate, administered at a dose of 600 mg qid for up to 14 days, was no more effective than placebo, based on upper airway viral clearance in patients with mild to moderate SARS-CoV-2 infection with or without symptoms. Furthermore, there were no differences between the study groups in terms of other efficacy endpoints. This study used a dose that was four to eight times higher than the clinical doses of camostat mesilate used in Japan for the acute symptoms of chronic pancreatitis and postoperative reflux oesophagitis. The study identified no additional safety concerns beyond those already known for camostat mesilate.

*Implications of all available evidence:* After starting this study, another randomised, placebo-controlled study reported the efficacy and safety of camostat mesilate for the treatment of patients with COVID-19, albeit at a lower dose of 200 mg three times daily. That study also found no difference between camostat mesilate and placebo for the primary endpoint (the time to discharge or a clinical improvement in clinical severity of at least two points on a seven-point ordinal scale). Along with this evidence, our study did not support the use of camostat mesilate as a treatment option for COVID-19. However, since the administration of camostat mesilate was started after the onset of symptoms and presumably the peak viral load, we cannot exclude the possibility that camostat mesilate may be effective if administration is started earlier in the course of infection, or perhaps as prophylactic use in close contacts.

## INTRODUCTION

SARS-CoV-2 is a highly transmissible virus that causes a potentially severe infection (COVID-19), which is continuing to spread worldwide and thus represents a significant global health threat.^1,2^ The symptoms and severity of COVID-19 vary considerably. Some patients develop advanced disease within about 10 days of onset, with life-threatening symptoms, including severe inflammatory reactions, dyspnoea, and severe acute pneumonia.^3,4^ Another challenge is the emergence of novel variants displaying altered transmissibility, infectiveness, disease severity, and mortality risk.

Currently, severe cases are generally treated with remdesivir, dexamethasone and symptomatic therapies. However, appropriate treatments have not been established for asymptomatic patients or patients with moderate symptoms who do not require oxygen therapy. Convenient oral drugs that can be prescribed to patients recuperating at home or other outpatient settings are also needed.

As one potential therapeutic target, it was discovered that the spike protein (S protein) of SARS-CoV-2 binds to angiotensin converting enzyme II (ACE2) on the host cell membrane as a functional receptor.^5,6^ The S protein is then cleaved into S1 and S2 by host-derived protease activity. The S1 fragment binds to ACE2 and the S2 fragment is cleaved by a type II transmembrane serine protease (TMPRSS2) expressed on the host cell membrane. These steps promote fusion of the viral envelope (outer membrane) with the cell membrane. Therefore, ACE2 and TMPRSS2, which are expressed on airway epithelial cells, are key factors in SARS-CoV-2 infection. *In vitro* drug-screening studies have indicated that 4-(4-guanidinobenzoyloxy)phenylacetic acid (GBPA), the active metabolite of the serine protease inhibitor camostat mesilate (FOY-305), inhibits TMPRSS2 and prevents SARS-CoV-2 infection of a human airway epithelial cell-derived cell line (Calu-3 cells).^6–13^

Repurposing drugs that have already been approved for other indications may help facilitate the drug development process and shorten the development time.^14,15^ In Japan, camostat mesilate is an oral drug that has been used to treat the acute symptoms of chronic pancreatitis and postoperative reflux oesophagitis for more than 30 years, and has shown a good safety profile over this period of time.^16^

Based on the preclinical evidence, it has been postulated that camostat mesilate may also be useful for treating COVID-19. In support of this hypothesis, one retrospective study of critically ill COVID-19 patients with organ failure treated in an intensive care unit revealed a decline in disease activity within 8 days of admission among patients treated with camostat mesilate but not in patients treated with hydroxychloroquine.^17^ This clinical improvement was accompanied by a decline in inflammatory markers, such as C-reactive protein and interleukin-6, and increased oxygenation.

This double-blind Phase 3 study was conducted to evaluate the efficacy and safety of camostat mesilate and hence explore the role of TMPRSS2 as a potential treatment target for mild to moderate SARS-CoV-2 infection with or without symptoms.

## METHODS

Further information about the design of this study, including patient eligibility, is available in the English version of the **study protocol** (**appendix**).

### Ethics

This study adhered to the Declaration of Helsinki, Good Clinical Practice, and relevant local/international guidelines. The protocol and patient consent forms were approved by the ethics committees or institutional review boards at all participating institutions (**appendix p 1**).

### Patients

Patients aged at least 18 years were eligible for this study if they were admitted to the participating hospitals within 5 days of onset of SARS-CoV-2 symptoms or within 5 days of a positive test for asymptomatic patients. SARS-CoV-2 infection must have been tested using a standard method at the time the study was conducted (e.g., reverse transcriptase-polymerase chain reaction [RT-PCR] test, loop-mediated isothermal amplification [LAMP] test, or antigen test). Only patients with asymptomatic/mild or moderate infection were eligible. Patients with severe infection, such as those requiring oxygenation, ventilation, or admission to an intensive care unit were excluded. Major exclusion criteria were prior history of SARS-CoV-2 infection, history of vaccination for SARS-CoV-2, and history of treatment with camostat mesilate or nafamostat mesilate. Further eligibility criteria are described in the **study protocol** (**appendix**). All patients provided informed consent.

### Study design

The study comprised a double-blind phase (up to 14 days) in which they were randomised to receive camostat mesilate or placebo, and a 2-week follow-up period after the last dose of the study drug.

Randomisation was performed using a minimisation method with the following randomisation factors: medical institution, age (at least 65 *vs* less than 65 years), and absence/presence of underlying diseases (chronic respiratory disease, chronic kidney disease, diabetes mellitus, hypertension, cardiovascular diseases, and obesity [body mass index, BMI, at least 30 kg/m^2^]). Patients were enrolled, randomised, and allocated to the appropriate treatments by the investigators/subinvestigators using an interactive web response system, which was managed by the sponsor.

The length of the double-blind period (up to 14 days) was chosen because of the typical clinical course.^18^

There were no changes to the study that were implemented after commencing enrolment.

### Interventions

Eligible patients were allocated to either camostat mesilate or placebo film-coated tablets, which were visually indistinguishable in appearance and packaging, to be administered at a dose of 600 mg four times daily (qid; before breakfast, before lunch, before dinner, and bedtime) for up to 14 days. The administration status was confirmed by clinical study staff, such as the principal investigator. The dose of camostat mesilate was chosen based on (1) preclinical EC_50_ values, which determined the clinical target exposures; (2) modelling and simulation to predict high dose exposure in the clinic; and (3) a results of a Phase 1 study of the safety and pharmacokinetics of camostat mesilate at 600 mg qid in healthy Japanese volunteers.^19^

During the study, it was prohibited to administer drugs with antiviral effects (e.g., remdesivir, favipiravir, ciclesonide, nafamostat mesilate, hydroxychloroquine, ivermectin, combination drug of lopinavir and ritonavir, povidone-iodine) and drugs with anticytokine effects (e.g., tocilizumab, Janus kinase inhibitors) from the day of onset of symptoms until completion of the study. However, these drugs could be continued at the same dose in patients already using these to treat a pre-existing comorbidity. The use of other unapproved drugs or camostat mesilate as a commercial product was prohibited.

In the randomised period, the allocated treatment was to be discontinued in accordance with the study criteria listed in **appendix p 4** (**table S1)**, which included patient request, emergence of an adverse event that made it difficult to continue the study, negative test for SARS-CoV-2 on two consecutive occasions, and increasing disease severity (exacerbation of pneumonia and SpO_2_ of 93% or less despite oxygen therapy). Efficacy evaluations were not conducted after confirmation of SARS-CoV-2 negativity. Treatments beyond day 14 were at the attending physician’s discretion or institutional policies and were not recorded.

### Endpoints

The primary efficacy endpoint was the time to the first two consecutive negative SARS-CoV-2 tests performed at the hospital’s local laboratory. The local tests were used for the primary endpoint in consideration of the time involved to send and analyse samples at the central laboratory and the potential difficulty of hospitalizing patients until the central laboratory had processed the tests.

Secondary efficacy endpoints were the time to the first two consecutive negative SARS-CoV-2 tests performed at the central laboratory, the proportion of patients testing negative for SARS-CoV-2, ordinal scale for disease severity (**appendix p 5 [table S2]**),^20^ the proportion of patients requiring mechanical ventilation, and survival.

Exploratory efficacy endpoints were the SARS-CoV-2 viral load (measured at the central laboratory), presence of lung lesions on chest imaging, time to resolution of clinical symptoms, proportion of patients in whom the clinical symptoms resolved, and antibody responses (IgM and IgG; measured at the central laboratory).

IgM and IgG targeting the spike protein of the virus were detected using a lateral flow immunofluorescence assay kit (SARS-CoV-2 IgM and IgG Quantum Dot Immunoassay, Mokobio Biotechnology R&D, Rockville, MD, USA). The fluorescence signal was semiquantified by an immunofluorescence analyser (Mokosensor-Q100, Mokobio Biotechnology R&D).

SARS-CoV-2 tests were performed daily throughout the treatment period at each hospital’s laboratory using the locally available methods. For the central evaluations, samples were sent to a central laboratory and analysed using a standardised RT-PCR method.

Safety evaluations included monitoring of adverse events, laboratory tests, vital signs, and 12-lead electrocardiography throughout the study.

### Statistical analyses

The objective of this study was to establish superiority of camostat mesilate over placebo in patients with COVID-19 using the time to negative SARS-CoV-2 test as the primary endpoint.

The criteria for effectiveness were a Bayesian posterior probability of at least 92% with a hazard ratio exceeding 1.0. The criteria for ineffectiveness were a Bayesian posterior probability of 8% or less with a hazard ratio exceeding 1.0.

The time to a negative SARS-CoV-2 test was assumed to follow an exponential distribution, with a median time of 14 days in the placebo group and a median time of 7 to 8 days in the camostat mesilate group. The probabilities of meeting the assessment of effectiveness or ineffectiveness with 50 patients per group (100 in total) were calculated at various analysis time-points by applying numerical simulations in SAS version 9.4. Because the timing of the interim analysis was dependent on the status of the COVID-19 outbreak, numerical simulations were performed, assuming the time-points when 40 to 80 subjects would have been randomised. The prior distribution of the regression coefficient was assumed to be uniform. Based on the numerical experiment, irrespective of which time-point the interim analysis was performed, the probability that treatment would be effective was 75% to 90% if the median duration was 7 or 8 days in the camostat mesilate group. Moreover, if the median duration was 14 days, the probability that treatment was effective was controlled within 10%. (**appendix p 2 [sample size calculation]**). From these data, we therefore considered it was possible to demonstrate superiority of camostat mesilate over placebo with a sample size of 100 patients (50 patients per group).

A modified intention-to-treat analysis set was used for efficacy analyses by excluding any patients who tested negative for SARS-CoV-2 on day 1 (local laboratory tests). All analyses were performed on an as-randomised basis. The safety analysis set comprised all patients who received at least one dose of the allocated drug.

Baseline characteristics were analysed using descriptive statistics, including numbers (proportions) of patients and summary statistics, as appropriate.

For the primary endpoint, the time to SARS-CoV-2 negativity (in days) was calculated as the date of the first (of two consecutive) SARS-CoV-2 negativity test minus the date of randomisation plus one. The events and reasons for censoring patients in this analysis are defined in **appendix p 6** (**table S3**). Cox proportional hazards model stratified by the randomisation factors (age and underlying disease) was used to determine the posterior mean hazard ratio with two-sided 95% credible intervals for the camostat mesilate group relative to the placebo group. The distribution of the regression coefficients was assumed to be uniform. In a secondary analysis, we applied the log-rank test stratified by age and underlying disease and plotted Kaplan–Meier curves for both groups to calculate the median time to SARS-CoV-2 negativity and 95% confidence intervals were calculated using the Brookmeyer–Crowley method with double log transformation. As a sensitivity analysis, we investigated the influence of patients who tested positive for SARS-CoV-2 *after* conversion to a SARS-CoV-2 negative status. The events and reasons for censoring patients in this analysis are defined in **appendix p 7** (**table S4**).

The proportions of patients negative for SARS-CoV-2, and the distribution of disease severity were compared between the two groups using the Mantel–Haenszel test stratified by the randomisation factors. The actual values for the ordinal scale of severity were compared between the treatment groups using the proportional odds model, which included treatment group and randomisation factors as factors. The median time to the resolution of clinical symptoms was estimated using the Kaplan–Meier method and 95% confidence intervals were calculated using the Brookmeyer–Crowley method with double log transformation. Changes in viral load, antibody responses (IgG and IgM), and safety outcomes were analysed descriptively in terms of the number and percentage of patients or summary statistics, as appropriate.

All tests were two-sided with a significance level of 5%. Because the primary analysis of the primary efficacy endpoint was based on Bayesian interim monitoring, a significance level was not applied. No adjustment for multiplicity between other endpoints or time-points was made.

Some changes in the statistical analyses were implemented before unblinding of the data. These additional analyses were performed to further evaluate efficacy, and are summarized in the **appendix p 8** (**table S5**).

SAS version 9.4 (SAS Institute, Cary, NC, USA) was used for all statistical analyses.

The study was registered on ClinicalTrials.gov (NCT04657497) and the Japan Registry for Clinical Trials (jRCT2031200198).

## RESULTS

### Patients

Because patient enrolment progressed as planned, no interim analyses were conducted. Between November 2020 and March 2021, a total of 161 patients provided consent and 155 patients were randomised across 21 participating institutions: 78 to camostat mesilate and 77 to placebo (**figure 1, appendix p 9** [**table S6**]). Four patients in the camostat mesilate group and three in the placebo group were excluded from the modified intention-to-treat population due to negative SARS-CoV-2 tests on day 1; thus, the modified intention-to-treat analysis set comprised 74 patients in each group. Because one patient in each group did not receive the allocated treatment, the safety analysis set comprised 77 patients in the camostat mesilate group and 76 in the placebo group.

**Figure 1.**
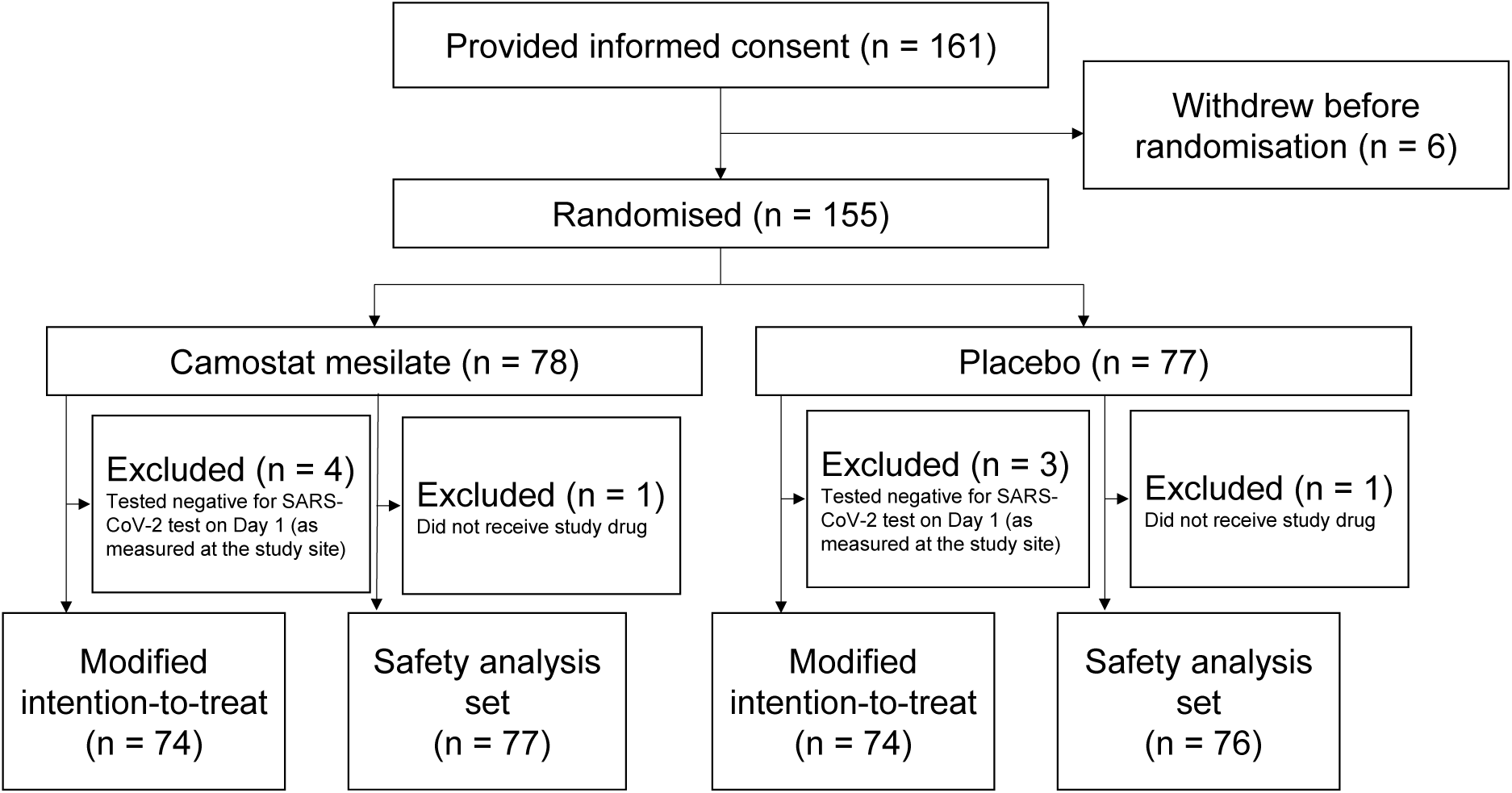
Patient disposition

Among 155 patients, 78 (50·3%) were male and 77 (49·7%) were female, 59 (38·1%) were at least 65 years old, and 71 (45·8%) had at least one underlying disease, the most common being hypertension in 44 patients (28·4%) (**table 1**). The median interval between the onset of symptoms (date of positive test for asymptomatic patients) to date of registration was 4 days (range 0–5 days). RT-PCR was the predominant testing methods, being used for 142 patients (91·6%). Nasopharyngeal swabs were used in 108 patients (69·7%), nasal swabs in 16 patents (10·3%), and saliva samples in 30 patients (19·4%). The median viral load was 6·91 log_10_ copies/mL (range 3·40–9·40 log_10_ copies/mL). All of the patients were hospitalised without requiring oxygen therapy (i.e. ordinal scale of 3). One hundred and eight patients had symptoms at registration. Both groups were very similar, demonstrating the robustness of the randomisation scheme.

**Table 1.** Patient characteristics

|  | **Camostat mesilate** | **Placebo** | **Total** |
| --- | --- | --- | --- |
|  | **(N = 78)** | **(N = 77)** | **(N = 155)** |
| Sex |  |  |  |
| Male | 35 (44·9) | 43 (55·8) | 78 (50·3) |
| Female | 43 (55·1) | 34 (44·2) | 77 (49·7) |
| Race |  |  |  |
| Asian | 78 (100·0) | 77 (100·0) | 155 (100·0) |
| Age at the time of consent (years) |  |  |  |
| ≥65 | 30 (38·5) | 29 (37·7) | 59 (38·1) |
| <65 | 48 (61·5) | 48 (62·3) | 96 (61·9) |
| Mean ± standard deviation | 55·7 ± 18·8 | 56·1 ± 18·2 | 55·9 ± 18·4 |
| Median (range) | 59·0 (21–89) | 56·0 (21–94) | 58·0 (21–94) |
| BMI (kg/m<sup>2</sup>) |  |  |  |
| Mean ± standard deviation | 24·5 ± 5·2 | 23·9 ± 3·7 | 24·2 ± 4·5 |
| Median (range) | 23·9 (14·1–46·2) | 23·2 (18·4–33·5) | 23·8 (14·1–46·2) |
| Underlying diseases<sup>a</sup> | 36 (46·2) | 35 (45·5) | 71 (45·8) |
| Chronic respiratory disease | 11 (14·1) | 14 (18·2) | 25 (16·1) |
| Chronic kidney disease | 5 (6·4) | 4 (5·2) | 9 (5·8) |
| Diabetes mellitus | 15 (19·2) | 12 (15·6) | 27 (17·4) |
| Hypertension | 24 (30·8) | 20 (26·0) | 44 (28·4) |
| Cardiovascular disease | 4 (5·1) | 4 (5·2) | 8 (5·2) |
|  | **(N = 78)** | **(N = 77)** | **(N = 155)** |
| Obesity (BMI $\geq 30$ kg/m<sup>2</sup>) | 9 (11·5) | 6 (7·8) | 15 (9·7) |
| Duration from the onset date<sup>b</sup> (days) |  |  |  |
| <4 | 43 (55·1) | 34 (44·2) | 77 (49·7) |
| $\geq 4$ | 35 (44·9) | 43 (55·8) | 78 (50·3) |
| Mean ± standard deviation | 3·3 ± 1·2 | 3·5 ± 1·1 | 3·4 ± 1·2 |
| Median (range) | 3·0 (0–5) | 4·0 (1–5) | 4·0 (0–5) |
| Test method used at the study site to<br>measure COVID-19 viral load |  |  |  |
| RT-PCR | 71 (91·0) | 71 (92·2) | 142 (91·6) |
| LAMP test | 7 (9·0) | 5 (6·5) | 12 (7·7) |
| Missing | 0 | 1 (1·3) | 1 (0·6) |
| Sample type used for negative/positive<br>determination |  |  |  |
| Nasopharyngeal swab | 56 (71·8) | 52 (67·5) | 108 (69·7) |
| Nasal swab | 9 (11·5) | 7 (9·1) | 16 (10·3) |
| Saliva | 13 (16·7) | 17 (22·1) | 30 (19·4) |
| Missing | 0 | 1 (1·3) | 1 (0·6) |
| SARS-CoV-2 viral load (central<br>laboratory) (log<sub>10</sub> copies/mL) |  |  |  |
| <7 | 39 (50·0) | 45 (58·4) | 84 (54·2) |
| $\geq 7$ | 39 (50·0) | 32 (41·6) | 71 (45·8) |
|  | **(N = 78)** | **(N = 77)** | **(N = 155)** |
| Mean ± standard deviation | 6.69 ± 1.48 | 6.41 ± 1.69 | 6.55 ± 1.59 |
| Median (range) | 6.98 (3.40–9.08) | 6.61 (3.40–9.40) | 6.91 (3.40–9.40) |
| Ordinal scale for severity |  |  |  |
| 3: Hospitalized, no oxygen therapy | 78 (100.0) | 77 (100.0) | 155 (100.0) |
| Presence of lung lesions | 38 (48.7) | 42 (54.5) | 80 (51.6) |
| IgM antibody test (central laboratory) |  |  |  |
| Positive | 1 (1.3) | 2 (2.6) | 3 (1.9) |
| Negative | 77 (98.7) | 75 (97.4) | 152 (98.1) |
| Mean ± standard deviation | 0.023 ± 0.081 | 0.042 ± 0.180 | 0.033 ± 0.139 |
| Median (range) | 0.010 (0.00–0.72) | 0.010 (0.00–1.14) | 0.010 (0.00–1.14) |
| IgG antibody test (central laboratory) |  |  |  |
| Positive | 2 (2.6) | 0 | 2 (1.3) |
| Negative | 76 (97.4) | 77 (100.0) | 153 (98.7) |
| Mean ± standard deviation | 0.078 ± 0.470 | 0.011 ± 0.009 | 0.045 ± 0.334 |
| Median (range) | 0.010 (0.00–3.91) | 0.010 (0.00–0.08) | 0.010 (0.00–3.91) |
| Presence of clinical symptoms | 55 (70.5) | 53 (68.8) | 108 (69.7) |

Values are n (%) unless otherwise stated
BMI, body mass index; RT-PCR, reverse transcriptase-polymerase chain reaction; LAMP, loop-mediated isothermal amplification; Ig, immunoglobulin
<sup>a</sup>Includes the following: chronic respiratory disease, chronic kidney disease, diabetes mellitus, hypertension, cardiovascular disease, obesity (BMI $\geq 30$ kg/m<sup>2</sup>) are included
<sup>b</sup>Duration from COVID-19 symptoms onset date (for asymptomatic patients, the collection date of sample
with positive confirmation) to the registration date

During the treatment period, 134 of 155 patients discontinued treatment or dropped out from the study: 68 of 78 (87·2%) in the camostat mesilate group and 66 of 77 (85·7%) in the placebo group (**appendix p 9** [**table S6**]). The most frequent reason for discontinuation of treatment was two consecutive negative SARS-CoV-2 tests in accordance with the study protocol in the camostat mesilate (45 patients, 57·7%) and placebo (43 patients, 55·8%) groups. Seven of 78 (9·0%) patients in the camostat mesilate group and six of 77 (7·8%) in the placebo group withdrew at the patient’s request.

### Time to SARS-CoV-2 negative test

The median time to the first two consecutive SARS-CoV-2 negative tests (local laboratory) was 11 days in both groups (**figure 2**), with conversion to negative status by day 14 in 45 of 74 patients (60·8%) in the camostat mesilate group and 47 of 74 patients (63·5%) in the placebo groups. The primary (Bayesian) and secondary (frequentist) analyses confirmed there was no significant difference in the primary endpoint between the two groups. Similar results were obtained in the results of the sensitivity analysis and central laboratory tests (**appendix p 10–11** [**tables S7 and S8**]). Subgroup analyses were also conducted to evaluate the potential influence of patient characteristics, such as underlying diseases and antibodies. However, there were no differences in the efficacy of camostat mesilate or placebo in terms of the time to a negative SARS-CoV-2 test among any of the subgroups evaluated, including the baseline viral load (**appendix p 12–13** [**table S9**]).

**Figure 2.**
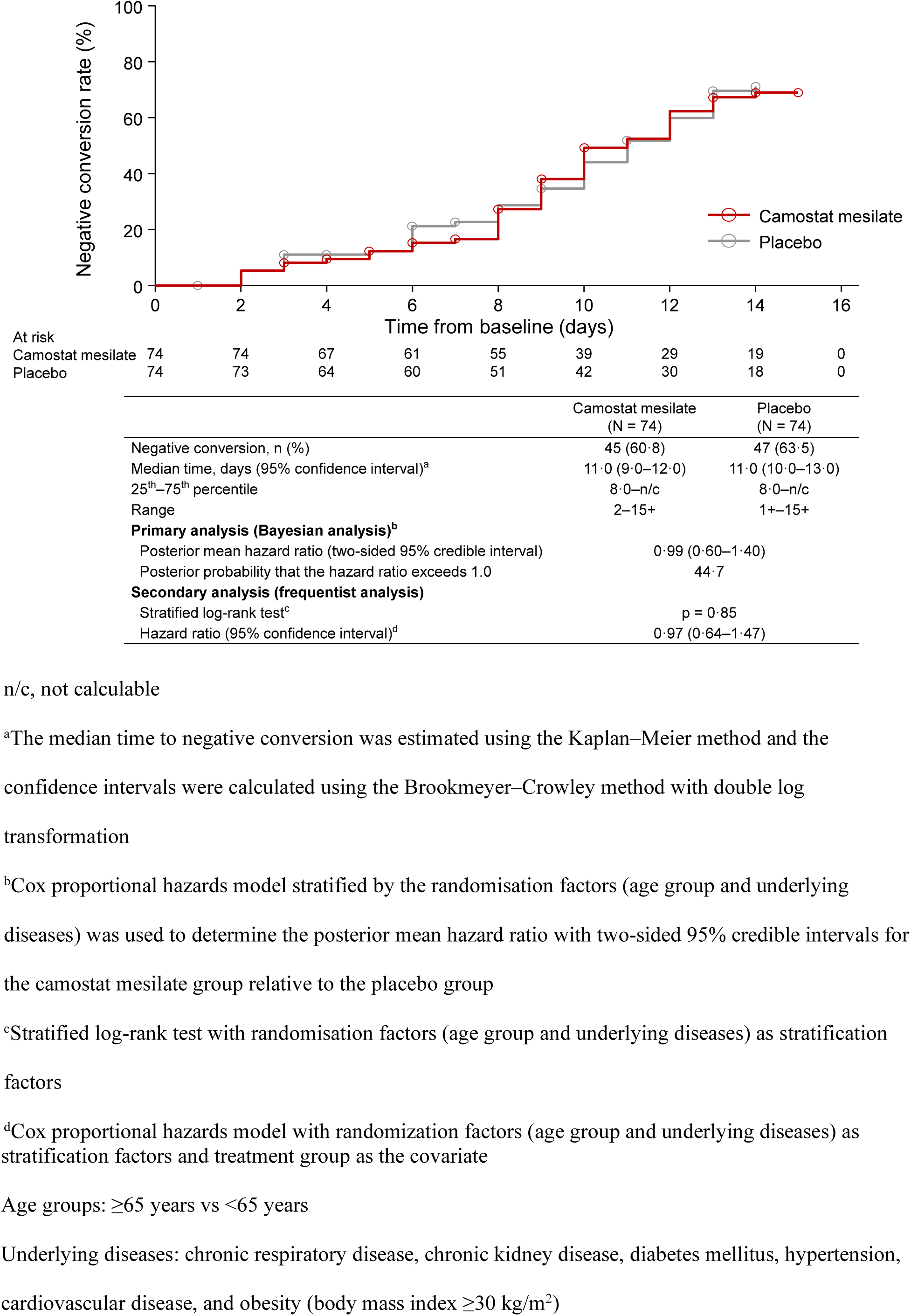
Time to SARS-CoV-2 negative conversion (local laboratory tests) n/c, not calculable ^a^The median time to negative conversion was estimated using the Kaplan–Meier method and the confidence intervals were calculated using the Brookmeyer–Crowley method with double log transformation ^b^Cox proportional hazards model stratified by the randomisation factors (age group and underlying diseases) was used to determine the posterior mean hazard ratio with two-sided 95% credible intervals for the camostat mesilate group relative to the placebo group ^c^Stratified log-rank test with randomisation factors (age group and underlying diseases) as stratification factors ^d^Cox proportional hazards model with randomization factors (age group and underlying diseases) as stratification factors and treatment group as the covariate Age groups: ≥65 years vs <65 years Underlying diseases: chronic respiratory disease, chronic kidney disease, diabetes mellitus, hypertension, cardiovascular disease, and obesity (body mass index ≥30 kg/m^2^)

### Viral load

The viral load was monitored daily in all patients. As illustrated in **figure 3**, the changes in viral load over time were comparable in both groups and there were no apparent differences at any time-point.

**Figure 3.**
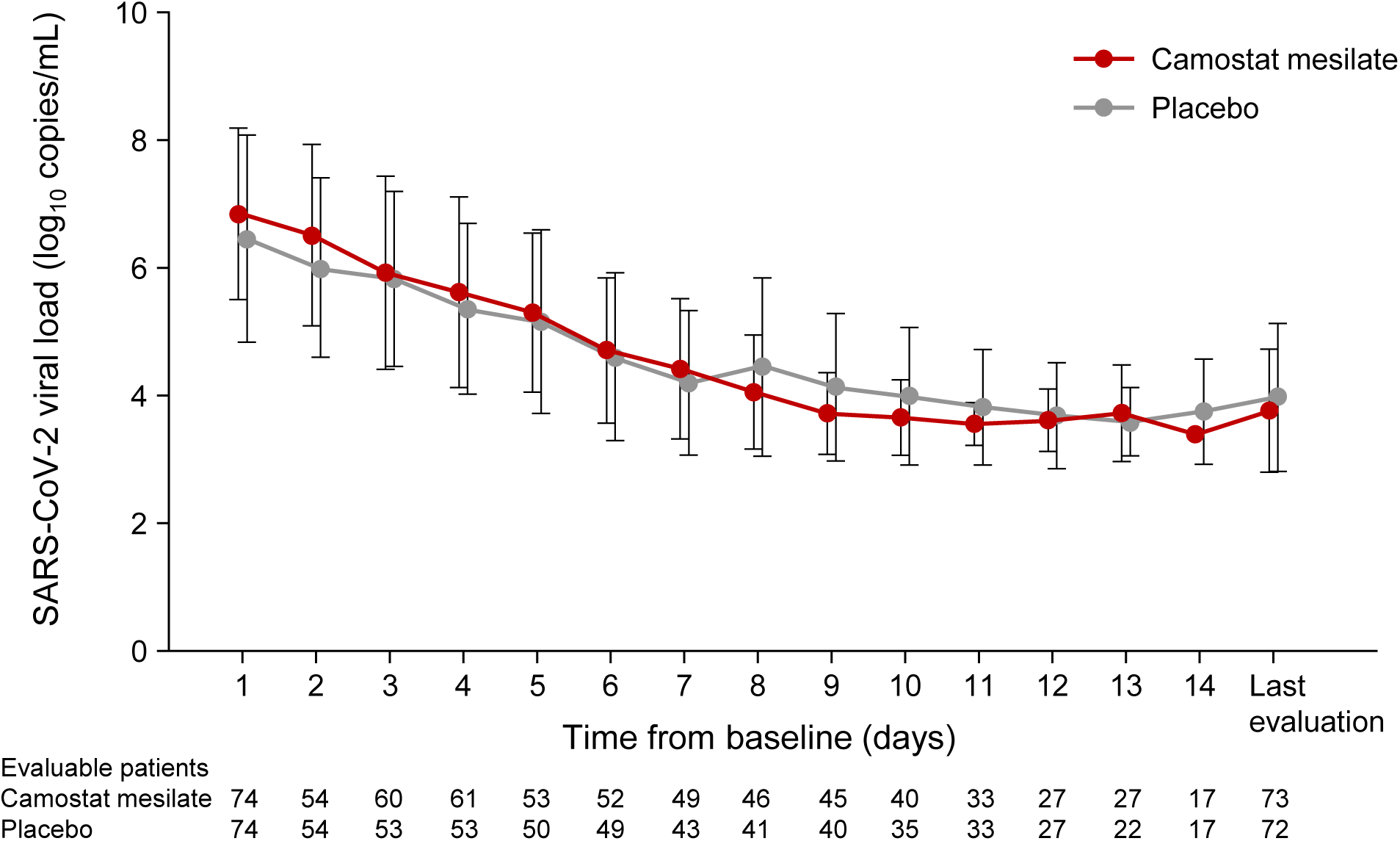
Change in SARS-CoV-2 viral load over time Values are mean ± standard deviation

### Ordinal scale of severity

The distribution of the ordinal scale of severity was comparable in both groups, with no clear differences at any time (**figure 4**). The ordinal scale was grade 3 (hospitalised, no oxygen therapy) in most patients during the study period because all patients were hospitalised for SARS-CoV-2 testing; outpatients were not enrolled due to the risk of transmission. Nevertheless, none of the patients in either group required intubation/mechanical ventilation or ventilation plus additional organ support, and there were no deaths.

**Figure 4.**
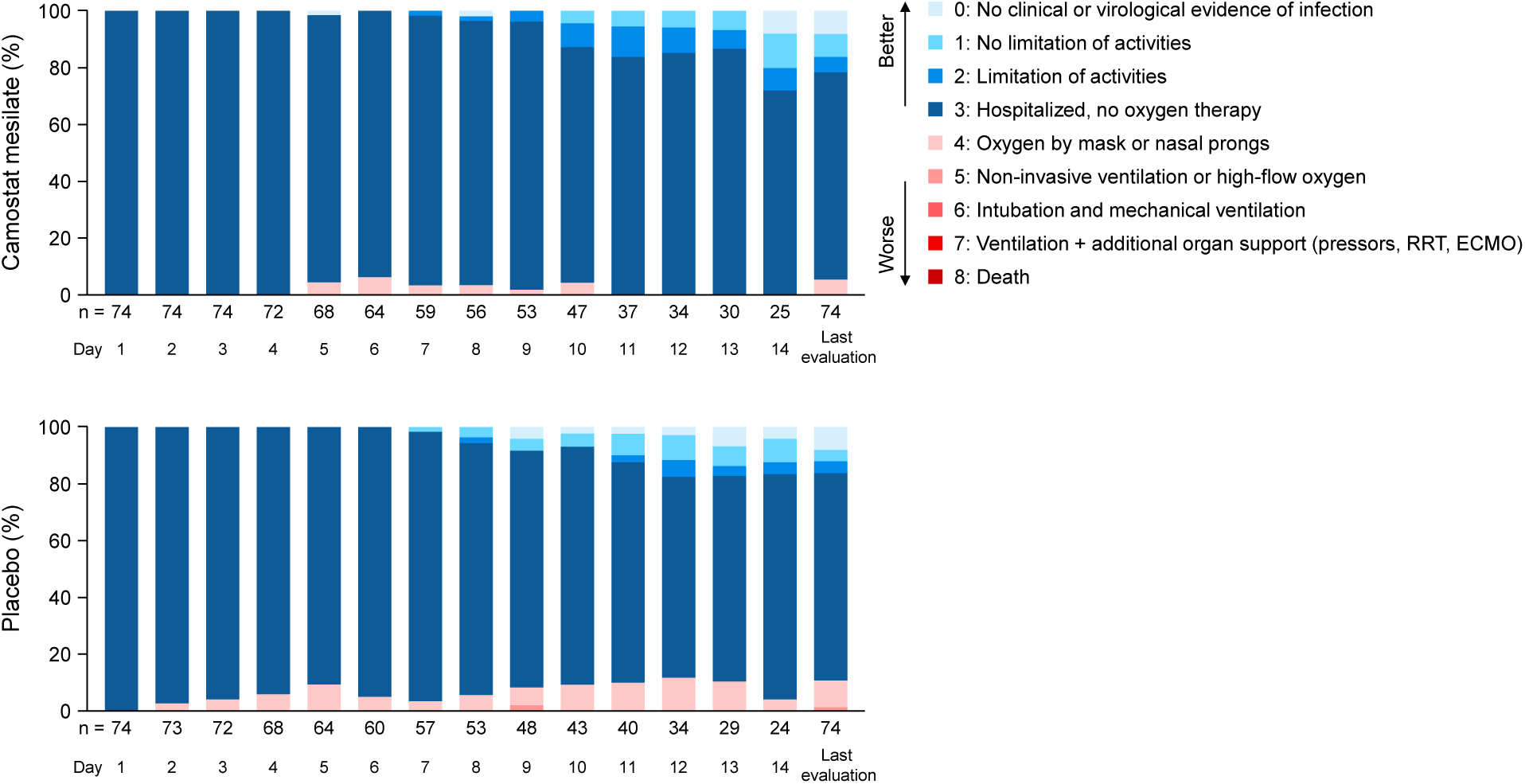
Ordinal scale for severity The vertical axes show the cumulative percentages of patients RRT, renal replacement therapy; ECMO, extracorporeal membrane oxygenation

The most severe case was a patient in the placebo group whose severity was classified as grade 5 (requiring non-invasive ventilation or high-flow oxygen therapy) on day 9. Other than the patient classified as grade 5 on day 9, none of the other patients in either group experienced a worsening in the ordinal scale by at least two categories at any time during the study.

### Resolution of clinical symptoms

The median time to resolution of clinical symptoms was 13 days in the camostat mesilate and 12 days in the placebo group (**table 2**).

**Table 2.** Time to resolution of clinical symptoms

|  | **Camostat mesylate** | **Placebo** |
| --- | --- | --- |
|  | **(N = 53)** | **(N = 52)** |
| Events, n (%) | 23 (43·4) | 23 (44·2) |
| Median (95% CI) (days) | 13·0 (10·0–n/c) | 12·0 (10·0–n/c) |
| 25<sup>th</sup> to 75<sup>th</sup> percentile | 7·0–15·0 | 9·0–n/c |
| Range | 2–16+ | 2–14+ |

+, censored, CI, confidence interval; n/c, not calculable

### Safety

Adverse events occurred in 25 of 77 patients in the camostat mesilate group (32·5%) and in 31 of 76 patients in the placebo group (40·8%) (**table 3**). A serious adverse event (prinzmetal angina) occurred in one patient in the camostat mesilate group, but it was not considered related to the study drug. Adverse drug reactions were reported in nine patients in the camostat mesilate group and in seven patients in the placebo group. Two patients in the camostat mesilate group discontinued treatment due to adverse drug reactions (hepatic function abnormal and drug eruption). The most common types of adverse drug reactions in the camostat mesilate group were gastrointestinal disorders (**table 3**).

**Table 3.** Safety

|  | **Camostat mesilate** | **Placebo** | **Total** |
| --- | --- | --- | --- |
|  | **(N = 77)** | **(N = 76)** | **(N = 153)** |
| Number of patients with |  |  |  |
| Any adverse events | 25 (32·5) | 31 (40·8) | 56 (36·6) |
| Any serious adverse events | 1 (1·3) | 0 | 1 (0·7) |
| Any adverse events that led to<br>discontinuation of treatment | 2 (2·6) | 0 | 2 (1·3) |
| Any adverse drug reactions | 9 (11·7) | 7 (9·2) | 16 (10·5) |
| Serious adverse drug reactions | 0 | 0 | 0 |
| Adverse drug reactions that led to<br>discontinuation of treatment | 2 (2·6) | 0 | 2 (1·3) |
| Adverse events or adverse drug reactions<br>resulting in death | 0 | 0 | 0 |
| Adverse drug reactions by system organ<br>class/preferred term |  |  |  |
| Gastrointestinal disorder | 4 (5·2) | 5 (6·6) | 9 (5·9) |
| Abdominal discomfort | 1 (1·3) | 1 (1·3) | 2 (1·3) |
| Constipation | 1 (1·3) | 0 | 1 (0·7) |
| Diarrhoea | 2 (2·6) | 1 (1·3) | 3 (2·0) |
| Nausea | 0 | 1 (1·3) | 1 (0·7) |
| Stomatitis | 0 | 1 (1·3) | 1 (0·7) |
| Vomiting | 0 | 1 (1·3) | 1 (0·7) |
| Hepatobiliary disorders | 1 (1·3) | 1 (1·3) | 2 (1·3) |
| Hepatic function abnormal | 1 (1·3) | 1 (1·3) | 2 (1·3) |
|  | **(N = 77)** | **(N = 76)** | **(N = 153)** |
| Laboratory test | 3 (3·9) | 2 (2·6) | 5 (3·3) |
| Alanine aminotransferase increased | 1 (1·3) | 2 (2·6) | 3 (2·0) |
| Aspartate aminotransferase increased | 0 | 1 (1·3) | 1 (0·7) |
| Blood potassium increased | 2 (2·6) | 0 | 2 (1·3) |
| Blood alkaline phosphatase increased | 0 | 1 (1·3) | 1 (0·7) |
| Skin and subcutaneous tissue disorders | 1 (1·3) | 1 (1·3) | 2 (1·3) |
| Drug eruption | 1 (1·3) | 0 | 1 (0·7) |
| Rash papular | 0 | 1 (1·3) | 1 (0·7) |

Values are n (%)

## DISCUSSION

SARS-CoV-2 remains a clinically significant global health crisis and there remains an urgent, ongoing need to identify effective treatments. A number of preclinical studies have been conducted in the search for therapies for COVID-19. It was discovered that GBPA, the active metabolite of camostat mesilate, inhibits TMPRSS2 and prevents SARS-CoV-2 infection of human airway cells.^6–13^ Furthermore, a small-scale retrospective study suggested a potential therapeutic effect of camostat mesilate in patients admitted to an intensive care unit.^21^

Despite promising results of preclinical studies, as well as a small retrospective study, the results of our study indicate that camostat mesilate is no more effective than placebo for treating patients with mild to moderate SARS-CoV-2 infection with or without symptoms. The lack of antiviral effect was demonstrated based on the median time to the first two consecutive SARS-CoV-2 negative tests (the primary endpoint) and the other clinical outcomes, with no statistically significant or clinically relevant differences between the two groups. Although this study investigated a high dose of camostat mesilate (600 mg qid; four to eight times higher than the clinical doses in Japan), no new safety concerns were identified.

Another large-scale study conducted in Denmark and Sweden also reported no benefit of administering camostat mesilate at a dose of 200 mg three times daily (lower than the dose used in our study, 600 mg qid) or placebo for 5 days in hospitalized patients.^22^ The primary endpoint in that study was a composite of the time to discharge or a clinical improvement in clinical severity of at least two points on a seven-point ordinal scale. The median time to clinical improvement was 5 days in both groups.

From the data available to date, it is unclear why the findings of preclinical studies did not translate into a clinical effect. However, several factors related to the design of the study and the mechanism of action of camostat mesilate should be considered:

### Viral entry pathway

Although TMPRSS2 is one of the primary routes of viral entry, the viral particles might exploit other pathways, such as endocytosis, to compensate for reduced entry via TMPRSS2. Thus, effective treatments may require inhibition of multiple viral entry pathways and a combination of drugs with various mechanisms of action, including camostat mesilate, may be useful for treating COVID-19.^23^

### Inappropriate timing of administration

The peak viral load is typically reached within 2–3 days after the onset of symptoms and the administration of camostat mesilate was started approximately 3 days after the onset of symptoms in this study. The selected timing of administration of camostat mesilate might not have been best optimised to suppress viral activity. It has been suggested that therapies aimed at blocking infection or viral reproduction may be more effective if they were initiated before the peak viral load.^24^ Therefore, some efficacy may be observed if administration is started as early as possible in the course of infection, perhaps as prophylactic administration to close contacts of patients, such as household members, or immediately after a positive test result. In fact, antibody drugs such as casirivimab/imdeviimab have been shown to reduce the risk of onset in uninfected patients and to prevent aggravation.^25^

### Dosing

Prior to this study, we conducted a Phase 1 study to set the dosage and treatment regimen in this study. An important PK feature of camostat mesilate is that when orally administered, it is rapidly metabolized into an active metabolite (GBPA) by esterases.^26–28^ Those studies demonstrated that camostat mesilate is not detectable in the human plasma, and that GBPA is rapidly eliminated with a half-life of less than 2 h. Therefore, frequent dosing is required to maintain target plasma concentrations. Considering the adherence of the target patient population, we assumed that qid administration of camostat mesilate (morning, midday, evening, and before bedtime) would be acceptable. Specifically, in PK/PD simulations, the times above the EC_50_ of camostat mesilate at doses of 800 mg three times daily and 600 mg qid were 9·8 h and 11·5 h, respectively.^19^ The results of the PK/PD simulations also suggested an advantage of increasing the dosing frequency rather than increasing the dose per administration. Multiple administrations of camostat mesilate at 600 mg qid were well tolerated in a Phase 1 study. However, in a repeated-dose toxicity study in dogs, camostat 300 mg/kg decreased body weight and food intake, induced vomiting and effects on the gastrointestinal tract, including gastrointestinal injury, and caused death, with a no-observed adverse effect level (NOAEL) of 100 mg/kg.^19,29^ Converting the NOAEL in dogs to humans yielded an equivalent dose of 3333 mg.^19,30^ Thus, the dose used in this study had a safety margin of 1·4-fold. Based on the overall balance between the expected time above EC_50_ and safety risks, 600 mg qid was determined to be an appropriate dose for this study. The plasma concentration of GBPA was predicted to exceed the EC_50_ for at least 11·5 h at the dose used (2400 mg/day).^19^ However, the targeted time above EC_50_ might have been insufficient to inhibit TMPRSS2 and hence prevent viral entry, although the exact relationship between the exposure and antiviral activity is not clear in the clinic.

A limitation of this study is that the improvement of the ordinal scale of severity could not be evaluated correctly because most patients were hospitalised for daily viral testing regardless of the presence or absence of symptoms and were hence classified as grade 3. Another possible limitation is that the effects of camostat mesilate against SARS-CoV-2 were evaluated using nasopharyngeal and nasal swab samples in the majority of patients. However, the appropriateness of an index of upper airway viral load in asymptomatic to moderate cases remains questionable. It is considered that the epidemic strain at the time was a D614G strain, but no data on the type of strain were collected for this study. Efficacy against currently circulating variants is unknown.

There are some strengths of this study that should be mentioned. In particular, this was a double-blind, randomised, placebo-controlled study with robust randomisation as demonstrated by the high similarity of both groups. In addition, this study used a dose that was four to eight times higher than the clinical doses in Japan used for the acute symptoms of chronic pancreatitis and postoperative reflux oesophagitis based on the preclinical and early clinical evidence. Furthermore, the efficacy of camostat mesilate was assessed using multiple clinically relevant endpoints, including local and central laboratory tests for SARS-CoV-2 infection and viral load.

Although the study results were negative, there were several lessons and the study generated important new evidence. There are still some questions related to the development of clinical trials for emerging infectious diseases, including study design and patient segmentation. Even in a state of emergency, the doses in clinical trials should be carefully selected with consideration of clinical pharmacology, including PK/PD modelling and simulation, when planning clinical trials for a new drug candidate in settings such as this, in order to provide clear evidence supporting or halting ongoing development of the drug. Furthermore, collaboration between government, industry and academia is essential for the development of therapeutic agents in a pandemic.

## Conclusions

In conclusion, the results of this study found clear evidence for not using camostat mesilate to treat mild to moderate SARS-CoV-2 infection with or without symptoms. Of note, no new safety concerns were identified at the high dose used in this study, which exceeds the standard dose used in other indications. Overall, these findings highlight the continuing need to identify and develop alternative therapies for COVID-19, and the necessity of conducting well-designed studies to confirm whether preclinical findings translate into meaningful clinical efficacy.

## Supporting information

Protocol

## Data Availability

Qualified researchers may request Ono Pharma to disclose individual patient-level data from clinical studies through the following website: https://www.clinicalstudydatarequest.com/. For more information on Ono Pharma’s Policy for the Disclosure of Clinical Study Data, please see the following website: https://www.ono.co.jp/eng/rd/policy.html.

## Appendix

- IRB information
- Sample size calculation
- 9 Tables
- CONSORT Checklist
- Study protocol (separate file)

## Acknowledgements

This study was funded by Ono Pharmaceutical Co., Ltd. The authors express their gratitude to all of the patients, investigators, and healthcare professionals involved in this study, and Yu Nakagama MD, PhD (Specially Appointed Senior Lecturer, Department of Parasitology, Graduate School of Medicine, Osaka City University) for research assistance and advice. The authors also thank Nicholas D. Smith (EMC K.K.) for medical writing support, which was funded by Ono Pharmaceutical Co., Ltd.

## Authors’ contributions

**Conceptualisation:** Naoyuki Komura, Junichi Kadota

**Data curation:** not applicable

**Formal analysis:** Kazuo Yoshida

**Funding acquisition:** not applicable

**Investigation:** Taku Kinoshita, Masahiro Shinoda, Yasuhiro Nishizaki, Katsuya Shiraki, Yuji Hirai, Yoshiko Kichikawa, Kenji Tsushima, Masaharu Sinkai

**Methodology:** Naoyuki Komura, Kazuo Yoshida, Yasutoshi Kido, Hiroshi Kakeya, Naoto Uemura, Junichi Kadota

**Project administration:** Naoyuki Komura, Yasutoshi Kido, Hiroshi Kakeya, Naoto Uemura, Junichi Kadota

**Resources:** Not applicable

**Software:** Not applicable

**Supervision:** Yasutoshi Kido, Hiroshi Kakeya, Naoto Uemura, Junichi Kadota

**Validation:** Not applicable

**Visualisation:** Not applicable

**Writing – original draft:** all authors

**Writing – review and editing:** all authors

**Data verification:** Naoyuki Komura, Kazuo Yoshida, Yasutoshi Kido, Hiroshi Kakeya, Naoto Uemura, Junichi Kadota

