## APPENDIX for "Phase 3, multicentre, double-blind, randomised, parallel-group, placebo-controlled study of camostat mesilate (FOY-305) for the treatment of COVID-19 (CANDLE study)"

##### Contents


| | Page |
|---|---|
| IRB information | 1 |
| Sample size calculation | 2 |
| Table S1. Discontinuation criteria | 4 |
| Table S2. Ordinal scale of severity | 5 |
| Table S3. Events and reasons for censoring patients for the analysis of time to SARS-CoV-2 negativity | 6 |
| Table S4. Events and reasons for censoring patients in the sensitivity analysis of the time to SARS-CoV-2 negativity | 7 |
| Table S5. Changes to the statistical analyses plan from the protocol | 8 |
| Table S6. Patient disposition | 9 |
| Table S7. Results of the sensitivity analysis of the primary endpoint | 10 |
| Table S8. Time to negative SARS-CoV-2 status as measured by the central laboratory | 11 |
| Table S9. Results of the subgroup analyses of time to negative SARS-CoV-2 status | 12 |
| CONSORT Checklist | 14 |


##### IRB Information

The protocol and patient consent forms were reviewed and approved by the following institutional review boards or ethics committees at each participating centre: Haradoi Hospital Institutional Review Board, Higashiosaka City Medical Center Institutional Review Board, Institutional Review Board of Tokyo Medical University Hachioji Medical Center, International University of Health and Welfare Institutional Review Board, Juntendo University Hospital Institutional Review Board, Kantorosai Hospital Institutional Review Board, Kobori Central Clinical Research Ethics Committee, Koseiren Hospital Central Institutional Review Board, Mishuku Hospital Institutional Review Board, National Hospital Organization Kanazawa Medical Center Institutional Review Board, Nihon University Hospitals' Joint Institutional Review Board, Okayama City Hospital Institutional Review Board, Review Board of Human Rights and Ethics for Clinical Studies Institutional Review Board, Review Board of Human Rights and Ethics for Clinical Studies Institutional Review Board, Saiseikai Kurihashi Hospital Institutional Review Board, Saitama Medical Center Institutional Review Board, Saitama Prefectural Cardiovascular and Respiratory Center Institutional Review Board, Tachikawa Hospital Institutional Review Board, Tokai University Hospital Group Institutional Review Board, Tokushukai Group Institutional Review Board, Tokyo Medical and Dental University Hospital Institutional Review Board, Tokyo Metropolitan Hiroo Hospital Institutional Review Board, Tokyo Shinagawa Hospital Institutional Review Board, Yokohama Municipal Citizen's Hospital Institutional Review Board, and Yokosuka Kyosai Hospital Institutional Review Board.

**Sample size calculation**

The objective of this study was to establish superiority of camostat mesilate over placebo in patients with COVID-19 using the time to negative SARS-CoV-2 test as the primary endpoint. Patients were to be randomised 1:1 to either camostat mesilate or placebo using a dynamic allocation procedure with site, age (≥65 *vs* <65 years), and presence/absence of underlying diseases (chronic respiratory disease, chronic kidney disease, diabetes mellitus, hypertension, cardiovascular diseases, obesity [body mass index ≥30 kg/m$^2$]) as allocation factors.

If enrolment became difficult due to convergence of COVID-19, an interim analysis was planned to assess the effectiveness or ineffectiveness at the time by applying Bayesian interim monitoring[1] with the criteria for assessment in the interim and final analyses (**table A**). The criteria for the assessment in the final analysis were kept regardless of whether an interim analysis was performed.

**Table A. Criteria for assessing effectiveness/ineffectiveness using the Bayesian posterior probability method**

| Time of analysis | Criteria for effectiveness | Criteria for ineffectiveness |
|---|---|---|
| Interim analysis | Bayesian posterior probability ≥97·5% for hazard ratio >1·0 | Bayesian posterior probability ≤2.5% for hazard ratio >1·0 AND Bayesian posterior probability ≥80·0% for hazard ratio <1·4 |
| Final analysis | Bayesian posterior probability ≥92% for hazard ratio >1·0 | Bayesian posterior probability ≤8% for hazard ratio >1·0 |

The time to negative SARS-CoV-2 test was assumed to follow an exponential distribution, with a median time of 14 days in the placebo group and a median time of 7–8 days in the camostat mesilate group. The probabilities of meeting the assessment of effectiveness or ineffectiveness with 50 subjects per group (100 in total) were calculated at various analysis time-points by applying numerical simulations in SAS version 9.4. Because the timing of the interim analysis was dependent on the status of the COVID-19 outbreak, numerical simulations were performed, assuming the time-points when 40 to 80 subjects would have been randomised. The prior distribution of the regression coefficient was assumed to be uniform. The numerical experiments were repeated 1,000 times under these conditions, and the probabilities of meeting the assessment of effectiveness and ineffectiveness at various time points were estimated as shown in **table B**.

**Table B. Simulated effectiveness and ineffectiveness rates according to predicted time to time to negative SARS-CoV-2 and number of patients**

| Median time (days) in the camostat mesilate group | Number of patients at the time of the interim analysis | Assessment | Interim analysis (%) | Final analysis (%) | Total (%) |
|---|---|---|---|---|---|
| 7 | 40 | Effective | 36·6 | 52·2 | 88·8 |
| | | Ineffective | 4·3 | 6·9 | 11·2 |
| | 50 | Effective | 44·1 | 46·2 | 90·3 |
| | | Ineffective | 2·4 | 7·3 | 9·7 |
| | 60 | Effective | 51·1 | 39·1 | 90·2 |
| | | Ineffective | 2·2 | 7·6 | 9·8 |
| | 70 | Effective | 57·5 | 32·5 | 90·0 |
| | | Ineffective | 1·5 | 8·5 | 10·0 |
| | 80 | Effective | 64·3 | 25·9 | 90·2 |
| | | Ineffective | 1·5 | 8·3 | 9·8 |
| 8 | 40 | Effective | 23·1 | 52·2 | 75·3 |
| | | Ineffective | 7·6 | 17·1 | 24·7 |
| | 50 | Effective | 29·8 | 45·5 | 75·3 |
| | | Ineffective | 6·5 | 18·2 | 24·7 |
| | 60 | Effective | 36·1 | 40·6 | 76·7 |
| | | Ineffective | 6·1 | 17·2 | 23·3 |
| | 70 | Effective | 39·1 | 38·0 | 77·1 |
| | | Ineffective | 4·6 | 18·3 | 22·9 |
| | 80 | Effective | 45·5 | 31·1 | 76·6 |
| | | Ineffective | 4·9 | 18·5 | 23·4 |
| 14 | 40 | Effective | 2·4 | 6·9 | 9·3 |
| | | Ineffective | 43·6 | 47·1 | 90·7 |
| | 50 | Effective | 2·6 | 6·6 | 9·2 |
| | | Ineffective | 47·2 | 43·6 | 90·8 |
| | 60 | Effective | 2·8 | 6·4 | 9·2 |
| | | Ineffective | 49·2 | 41·6 | 90·8 |
| | 70 | Effective | 2·2 | 6·8 | 9·0 |
| | | Ineffective | 53·7 | 37·3 | 91·0 |
| | 80 | Effective | 2·3 | 6·2 | 8·5 |
| | | Ineffective | 56·0 | 35·5 | 91·5 |

Based on the numerical experiment, irrespective of which time-point the interim analysis was performed, the probability that treatment would be effective was 75% to 90% if the median duration was 7 or 8 days in the camostat mesilate group. Moreover, if the median duration was 14 days, the probability that treatment was effective was controlled within 10%. From these data, we therefore considered it was possible to demonstrate superiority of camostat mesilate over placebo with a sample size of 100 patients (50 patients per group).

**Table S1. Discontinuation criteria**

| No. | Criterion |
|---|---|
| 1 | The patient requested to withdraw from the study |
| 2 | The patient's representative requested to withdraw the patient from the study |
| 3 | The patient did not meet the inclusion criteria or met the exclusion criteria |
| 4 | Continuation of the study was considered difficult because of adverse event(s), whether or not related to the study drug, in the opinion of the investigator or subinvestigator |
| 5 | The patient tested negative for SARS-CoV-2 on two consecutive occasions |
| 6 | If criteria *a* and *b* are both met, and continuation of the study was considered difficult:<br>a. Exacerbation of pneumonia was observed by chest imaging<br>b. Oxygen saturation was ≤93% despite oxygen therapy |
| 7 | The patient received a prohibited concomitant medication |
| 8 | The patient died |
| 9 | The patient was found to be pregnant |
| 10 | Continuation of the study was not appropriate for any other reason(s), in the opinion of the investigator or subinvestigator |

**Table S2. Ordinal scale of severity[1]**

| Patient state | Description | Grade |
|---|---|---|
| Uninfected | - No clinical or virological evidence of infection | 0 |
| Ambulatory | - No limitation of activities | 1 |
| | - Limitations of activities | 2 |
| Hospitalised | | |
| Mild disease | - Hospitalised, no oxygen therapy | 3 |
| | - Oxygen therapy with a mask or nasal prongs | 4 |
| Severe disease | - Non-invasive ventilation or high-flow oxygen therapy | 5 |
| | - Intubation and mechanical ventilation | 6 |
| | - Ventilation and additional organ support (e.g., pressors, renal replacement therapy, extracorporeal membrane oxygenation) | 7 |
| Dead | - Death | 8 |

1 World Health Organization R&D Blue Print Team. WHO R&D Blueprint: Novel Coronavirus - COVID-19 Therapeutic Trial Synopsis. Available at: https://www.who.int/publications/i/item/covid-19-therapeutic-trial-synopsis. Accessed: July 22, 2021

**Table S3. Events and reasons for censoring patients for the analysis of time to SARS-CoV-2 negativity**

| Status | Event or censored date | Result |
|---|---|---|
| SARS-CoV-2 negativity was determined | Sampling date of the sample tested negative for the first time for the SARS-CoV-2 test (measured at the study site) | Event |
| SARS-CoV-2 negativity was not determined | Sampling date of the last assessable SARS-CoV-2 test (measured at the study site) | Censored |
| Died | Day 14 | Censored |
| The patient met criteria for discontinuing the study drug (study drug discontinuation criteria 6 or 7) for worsening of COVID-19, and administration of the study drug was discontinued | Day 14 | Censored |
| Study was being discontinued | Sampling date of the last assessable SARS-CoV-2 test (measured at the study site) | Censored |
| SARS-CoV-2 test (measured at the study site) was not performed more than once after randomisation | Day 1 | Censored |

**Table S4. Events and reasons for censoring patients in the sensitivity analysis of the time to SARS-CoV-2 negativity**

| Status | Event or censored date | Result |
|---|---|---|
| Patient tested negative for SARS-CoV-2, and did not test positive for SARS-CoV-2 thereafter | Sampling date of the sample tested negative for the first time for the SARS-CoV-2 test (measured at the study site) | Event |
| There were several determinations of SARS-CoV-2 negativity | Sampling date of the sample tested negative for the first time for the SARS-CoV-2 test at the latest determination (measured at the study site). | Event |
| Patient tested positive for SARS-CoV-2 after a negative test for SARS-CoV-2 | Sampling date of the last assessable SARS-CoV-2 test (measured at the study site) | Censored |
| SARS-CoV-2 negativity was not determined | Sampling date of the last assessable SARS-CoV-2 test (measured at the study site) | Censored |
| Died | Day 14 | Censored |
| The patient met criteria for discontinuing the study drug (study drug discontinuation criteria 6 or 7) for worsening of COVID-19, and administration of the study drug was discontinued | Day 14 | Censored |
| Study was being discontinued | Sampling date of the last assessable SARS-CoV-2 test (measured at the study site) | Censored |
| SARS-CoV-2 test (measured at the study site) was not performed more than once after randomisation | Day 1 | Censored |

**Table S5. Changes to the statistical analyses plan from the protocol**

| Description in the protocol | Description in the statistical analysis plan | Reason for change |
|---|---|---|
| None | 9 Primary efficacy endpoint<br>9.3 Analysis methods<br>Addition of sensitivity analysis | To investigate the effect of patients who tested positive for SARS-CoV-2 test after their SARS-CoV-2 test became negative |
| 10.5.5 Secondary efficacy endpoint<br>10.5.5.2.1 Analysis item<br>Percentage of patient with a negative SARS-CoV-2 | 10 Secondary efficacy endpoints<br>10.2 Analysis items and data handling<br>Cumulative proportion of patient with a negative SARS-CoV-2 | Based on the definition of the determination of negative conversion and the status of data collection after the determination of negative conversion.<br>To evaluate effectiveness in more detail |
| None | 10 Secondary efficacy endpoints<br>10.3 Analysis methods<br>Addition of summary statistics to the ordinal scale of severity | To evaluate effectiveness in more detail |
| 10.5.5 Secondary efficacy endpoint<br>10.5.5.3 Analysis method<br>Analysis method for survival | 10 Secondary efficacy endpoints<br>10.3 Analysis methods<br>Only events, number of censored patients, reasons for censoring, and Kaplan–Meier curves were analysed for survival. | Data on survival status were confirmed in a blinded manner, and detailed analysis was considered unnecessary |
| None | 11 Exploratory efficacy endpoints<br>11.2 Analysis items and data handling<br>Time to onset of clinical symptoms and addition of the proportion of patients with clinical symptoms | To evaluate effectiveness in more detail |
| None | 11 Exploratory efficacy endpoints<br>11.3 Analysis methods<br>The addition of an analysis of the difference in the proportion between the treatment groups to the presence or absence of lung lesions in chest images, the proportion of patients in whom clinical symptoms disappeared, and the proportion of patients in whom clinical symptoms occurred | To evaluate effectiveness in more detail |
| 8.2.5.2 Laboratory tests<br>Haematological examination | 12.2 Analysis items and data handling<br>1) Analysis item<br>(2) Laboratory test: haematological examination<br>Add total neutrophil count | Because some facilities could not measure the segmented and stab cells separately, an additional item was added to sum the segmented and stab cells |

**Table S6. Patient disposition**

| | Camostat mesilate (N = 78) | Placebo (N = 77) | Total (N = 155) |
|---|---|---|---|
| At the end of the observation period | | | |
| Number of patients who discontinued or dropped out | 0 | 0 | 0 |
| At the end of the treatment period | | | |
| Number of patients who discontinued or dropped out | 68 (87·2) | 66 (85·7) | 134 (86·5) |
| Reason for discontinuation or dropout | | | |
| Withdrawn at the patient's request | 7 (9·0) | 6 (7·8) | 13 (8·4) |
| Withdrawn at the guardian's request | 0 | 0 | 0 |
| Adverse event | 2 (2·6) | 0 | 2 (1·3) |
| Physician's decision | 9 (11·5) | 11 (14·3) | 20 (12·9) |
| Protocol deviation | 0 | 0 | 0 |
| Pregnancy | 0 | 0 | 0 |
| Lost to follow-up | 0 | 0 | 0 |
| Non-compliance with study drug | 0 | 0 | 0 |
| Study terminated by sponsor | 0 | 0 | 0 |
| Death | 0 | 0 | 0 |
| Protocol-specified withdrawal criterion met | 47 (60·3) | 48 (62·3) | 95 (61·3) |
| Discontinuation criterion 5 | 45 (57·7) | 43 (55·8) | 88 (56·8) |
| Discontinuation criterion 6 | 2 (2·6) | 5 (6·5) | 7 (4·5) |
| Other | 3 (3·8) | 1 (1·3) | 4 (2·6) |
| At the end of the study | | | |
| Number of patients who discontinued or dropped out | 3 (3·8) | 5 (6·5) | 8 (5·2) |
| Reason for discontinuation or dropout | | | |
| Withdrawn at the patient's request | 1 (1·3) | 1 (1·3) | 2 (1·3) |
| Withdrawn at the guardian's request | 0 | 0 | 0 |
| Adverse event | 0 | 0 | 0 |
| Death | 0 | 1 (1·3) | 1 (0·6) |
| Pregnancy | 0 | 0 | 0 |
| Other | 2 (2·6) | 3 (3·9) | 5 (3·2) |

Values are n (%)

#### Table S7. Results of the sensitivity analysis of the primary endpoint

| | Camostat mesilate (N = 74) | Placebo (N = 74) |
|---|---|---|
| Negative conversion, n (%) | 42 (56·8) | 45 (60·8) |
| Number of censored patients, n (%) | 32 (43·2) | 29 (39·2) |
| Reasons for censoring patients, n (%) | | |
| The patient tested positive for SARS-CoV-2 after a negative test for SARS-CoV-2 | 3 (9·4) | 2 (6·9) |
| SARS-CoV-2 negativity was not confirmed | 27 (84·4) | 21 (72·4) |
| Death | 0 | 1 (3·4) |
| The patient met criteria for discontinuing the study drug (study drug discontinuation criteria 6 or 7) for worsening of COVID-19, and administration of the study drug was discontinued | 2 (6·3) | 4 (13·8) |
| Patient discontinued the study | 0 | 0 |
| SARS-CoV-2 test (measured at the study site) was not performed more than once after randomisation | 0 | 1 (3·4) |
| Analysis of time (in days) to a negative test | | |
| Median time (95% confidence interval)[a] | 12·0 (10·0–13·0) | 11·0 (10·0–13·0) |
| 25th to 75th percentile[a] | 8·0–n/c | 8·0–n/c |
| Range | 2–15+ | 1+–14+ |
| Main analysis (Bayesian analysis) | | |
| Posterior average of hazard ratio (two-sided 95% credible interval)[b] | 0·96 (0·57–1·36) | |
| Posterior probability that the hazard ratio exceeds 1.0[b] | 38·0 | |
| Secondary analysis (frequency analysis) | | |
| Stratified log-rank test[c] | p = 0·71 | |
| Hazard ratio (95% confidence interval)[b] | 0·94 (0·61–1·43) | |

+, censored; n/c, not calculable

[a]Estimated using the Kaplan–Meier method. The Brookmeyer and Crowley method with double log transformation was used to calculate the confidence interval

[b]Randomisation factors (age [≥65 years *vs* <65 years] and presence of underlying diseases [chronic respiratory disease, chronic kidney disease, diabetes mellitus, hypertension, cardiovascular disease, and obesity (BMI ≥30 kg/m$^2$)]) were estimated using a stratified Cox proportional hazards model with stratification factors and treatment group as covariates

[c]Stratified log-rank test with randomisation factors (age [≥65 years *vs* <65 years] and presence of underlying diseases [chronic respiratory disease, chronic kidney disease, diabetes mellitus, hypertension, cardiovascular disease, obesity (BMI ≥30 kg/m$^2$)]) as stratification factors

**Table S8. Time to negative SARS-CoV-2 status as measured at the central laboratory**

| | **Camostat mesilate (N = 73)** | **Placebo (N = 72)** |
|---|---|---|
| Negative conversion, n (%) | 30 (41·1) | 30 (41·7) |
| Number of censored patients, n (%) | 43 (58·9) | 42 (58·3) |
| Reasons for censoring patients, n (%) | | |
| SARS-CoV-2 negative status not confirmed | 39 (90·7) | 33 (78·6) |
| Death | 0 | 1 (2·4) |
| The patient met criteria for discontinuing the study drug (study drug discontinuation criteria 6 or 7) for worsening of COVID-19, and administration of the study drug was discontinued | 2 (4·7) | 3 (7·1) |
| Patient has discontinued the study | 0 | 0 |
| SARS-CoV-2 test (measured at the study site) was not performed more than once after randomisation | 2 (4·7) | 5 (11·9) |
| Analysis of time (in days) to a negative test | | |
| Median time (95% confidence interval) [a] | 12·0 (10·0–n/c) | 12·0 (10·0–n/c) |
| 25th to 75th percentile[a] | 9·0–n/c | 9·0–n/c |
| Range | 1+–15+ | 1+–14+ |
| Main analysis (Bayesian analysis) | | |
| Posterior average of hazard ratio (two-sided 95% credible interval)[b] | 0·86 (0·45–1·31) | |
| Posterior probability that the hazard ratio exceeds 1.0 [b] | 23·8 | |
| Secondary analysis (frequency analysis) | | |
| Stratified log-rank test[c] | p = 0·51 | |
| Hazard ratio (95% confidence interval)[b] | 0·83 (0·50–1·39) | |

+, censored; n/c, not calculable

[a]Estimated using the Kaplan–Meier method. The Brookmeyer and Crowley method with double log transformation was used to calculate the confidence interval

[b]Randomisation factors (age [≥65 years *vs* <65 years] and presence of underlying diseases [chronic respiratory disease, chronic kidney disease, diabetes mellitus, hypertension, cardiovascular disease, and obesity (BMI ≥30 kg/m$^2$)]) were estimated using a stratified Cox proportional hazards model with the stratification factors and treatment group as covariates

[c]Stratified log-rank test with randomisation factors (age [≥65 years *vs* <65 years] and presence of underlying diseases [chronic respiratory disease, chronic kidney disease, diabetes mellitus, hypertension, cardiovascular disease, and obesity (BMI ≥30 kg/m$^2$)]) as stratification factors

**Table S9. Results of the subgroup analyses of time to negative SARS-CoV-2 status**

| **Characteristic** | | | **Camostat mesilate (N = 74)** | **Placebo (N = 74)** |
|---|---|---|---|---|
| Age (years) | ≥65 | Number of events | 15 (51·7) | 15 (53·6) |
| | | Number censored | 14 (48·3) | 13 (46·4) |
| | | Median (95% CI)[a] | 12·0 (9·0–n/c) | 13.0 (10·0–n/c) |
| | | Hazard ratio (95% CI) | 1·05 (0·51–2·16) | |
| | <65 | Number of events | 30 (66·7) | 32 (69·6) |
| | | Number censored | 15 (33·3) | 14 (30·4) |
| | | Median (95% CI)[a] | 11·0 (8·0–12·0) | 11·0 (8·0–12·0) |
| | | Hazard ratio (95% CI) | 0·94 (0·57–1·55) | |
| Age (years) | ≥75 | Number of events | 7 (53·8) | 5 (38·5) |
| | | Number censored | 6 (46·2) | 8 (61·5) |
| | | Median (95% CI)[a] | 10·5 (8·0–n/c) | n/c (9·0–n/c) |
| | | Hazard ratio | 2·03 (0·64–6·46) | |
| | ≥65 to <75 | Number of events | 8 (50·0) | 10 (66·7) |
| | | Number censored | 8 (50·0) | 5 (33·3) |
| | | Median (95% CI)[a] | 12·0 (9·0–n/c) | 10·0 (8·0–13·0) |
| | | Hazard ratio | 0·64 (0·25–1·64) | |
| | <65 | Number of events | 30 (66·7) | 32 (69·6) |
| | | Number censored | 15 (33·3) | 14 (30·4) |
| | | Median (95% CI)[a] | 11·0 (8·0–12·0) | 11·0 (8·0–12·0) |
| | | Hazard ratio (95% CI) | 0·94 (0·57–1·55) | |
| BMI (kg/$m^2$) | ≥30 | Number of events | 6 (66·7) | 4 (66·7) |
| | | Number censored | 3 (33·3) | 2 (33·3) |
| | | Median (95% CI)[a] | 11·0 (2·0–n/c) | 13·0 (11·0–n/c) |
| | | Hazard ratio (95% CI) | 2·11 (0·55–8·15) | |
| | <30 | Number of events | 39 (60·0) | 43 (63·2) |
| | | Number censored | 26 (40·0) | 25 (36·8) |
| | | Median (95% CI)[a] | 11·0 (9·0–13·0) | 11·0 (9·0–13·0) |
| | | Hazard ratio (95% CI) | 0·91 (0·59–1·41) | |
| Chronic respiratory disease | Yes | Number of events | 7 (63·6) | 8 (61·5) |
| | | Number censored | 4 (36·4) | 5 (38·5) |
| | | Median (95% CI)[a] | 10·0 (3·0–n/c) | 12·0 (7·0–n/c) |
| | | Hazard ratio (95% CI) | 1·50 (0·49–4·62) | |
| | No | Number of events | 38 (60·3) | 39 (63·9) |
| | | Number censored | 25 (39·7) | 22 (36·1) |
| | | Median (95% CI)[a] | 11·0 (9·0–13·0) | 11·0 (9·0–13·0) |
| | | Hazard ratio (95% CI) | 0·93 (0·59–1·46) | |
| Chronic kidney disease | Yes | Number of events | 4 (80·0) | 2 (50·0) |
| | | Number censored | 1 (20·0) | 2 (50·0) |
| | | Median (95% CI)[a] | 9·0 (2·0–12·0) | n/c (9·0–n/c) |
| | | Hazard ratio (95% CI) | 71016179·23 (0·00–n/c) | |
| | No | Number of events | 41 (59·4) | 45 (64·3) |
| | | Number censored | 28 (40·6) | 25 (35·7) |
| | | Median (95% CI)[a] | 11·0 (10·0–13·0) | 11·0 (10·0–12·0) |
| | | Hazard ratio (95% CI) | 0·91 (0·59–1·39) | |
| Diabetes mellitus | Yes | Number of events | 10 (71·4) | 5 (41·7) |
| | | Number censored | 4 (28·6) | 7 (58·3) |
| | | Median (95% CI)[a] | 9·0 (5·0–10·0) | n/c (8·0–n/c) |
| | | Hazard ratio (95% CI) | 3·78 (1·02–14·04) | |
| | No | Number of events | 35 (58·3) | 42 (67·7) |
| | | Number censored | 25 (41·7) | 20 (32·3) |
| | | Median (95% CI)[a] | 12·0 (10·0–14·0) | 11·0 (10·0–13·0) |
| | | Hazard ratio(95% CI) | 0·78 (0·50–1·22) | |
| Hypertension | Yes | Number of events | 14 (60·9) | 9 (47·4) |
| | | Number censored | 9 (39·1) | 10 (52·6) |
| | | Median (95% CI)[a] | 11·0 (9·0–n/c) | 13·0 (9·0–n/c) |
| | | Hazard ratio (95% CI) | 1·16 (0·44–3·07) | |
| | No | Number of events | 31 (60·8) | 38 (69·1) |
| | | Number censored | 20 (39·2) | 17 (30·9) |
| | | Median (95% CI)[a] | 11·0 (9·0–13·0) | 11·0 (9·0–13·0) |
| | | Hazard ratio (95% CI) | 0·87 (0·53–1·42) | |
| Cardiovascular disease | Yes | Number of events | 1 (25·0) | 1 (25·0) |
| | | Number censored | 3 (75·0) | 3 (75·0) |
| | | Median (95% CI)[a] | n/c (12·0–n/c) | n/c (13·0–n/c) |

| Characteristic | | | Camostat mesilate (N = 74) | Placebo (N = 74) |
|---|---|---|---|---|
| | | Hazard ratio (95% CI) | 1·00 (1·00–1·00) | |
| | No | Number of events | 44 (62·9) | 46 (65·7) |
| | | Number censored | 26 (37·1) | 24 (34·3) |
| | | Median (95% CI)[a] | 10·0 (9·0–12·0) | 11·0 (10·0–12·0) |
| | | Hazard ratio (95% CI) | 0·93 (0·62–1·42) | |
| Time (days) from sample collection date to confirmation of SARS-CoV-2 positive status in patients who were asymptomatic at the registration date | <4 | Number of events | 29 (72·5) | 15 (46·9) |
| | | Number censored | 11 (27·5) | 17 (53·1) |
| | | Median (95% CI)[a] | 10·0 (9·0–12·0) | 13·0 (10·0–n/c) |
| | | Hazard ratio (95% CI) | 1·76 (0·93–3·32) | |
| | ≥4 | Number of events | 16 (47·1) | 32 (76·2) |
| | | Number censored | 18 (52·9) | 10 (23·8) |
| | | Median (95% CI)[a] | 13·0 (9·0–n/c) | 10·0 (9·0–12·0) |
| | | Hazard ratio (95% CI) | 0·56 (0·30–1·02) | |
| SARS-CoV-2 viral load (central testing) ($\log_{10}$ copies/mL) | <7 | Number of events | 26 (74·3) | 33 (76·7) |
| | | Number censored | 9 (25·7) | 10 (23·3) |
| | | Median (95% CI)[a] | 9·0 (8·0–9·0) | 10·0 (8·0–12·0) |
| | | Hazard ratio (95% CI) | 1·15 (0·67–1·98) | |
| | ≥7 | Number of events | 19 (48·7) | 14 (45·2) |
| | | Number censored | 20 (51·3) | 17 (54·8) |
| | | Median (95% CI)[a] | 13·0 (12·0–n/c) | 13·0 (10·0–n/c) |
| | | Hazard ratio (95% CI) | 0·92 (0·46–1·87) | |
| Presence of pulmonary lesions | Yes | Number of events | 23 (63·9) | 27 (67·5) |
| | | Number censored | 13 (36·1) | 13 (32·5) |
| | | Median (95% CI)[a] | 10·0 (8·0–12·0) | 11·0 (8·0–13·0) |
| | | Hazard ratio (95% CI) | 1·05 (0·60–1·85) | |
| | No | Number of events | 22 (57·9) | 20 (58·8) |
| | | Number censored | 16 (42·1) | 14 (41·2) |
| | | Median (95% CI)[a] | 12·0 (10·0–14·0) | 12·0 (10·0–13·0) |
| | | Hazard ratio (95% CI) | 0·86 (0·47–1·59) | |
| IgM antibody test (central testing) | Positive | Number of events | 1 (100·0) | 1 (50·0) |
| | | Number censored | 0 ( 0·0) | 1 (50·0) |
| | | Median (95% CI)[a] | 11·0 (n/c–n/c) | n/c (2·0–n/c) |
| | | Hazard ratio (95% CI) | 1·00 (1·00–1·00) | |
| | Negative | Number of events | 44 (60·3) | 46 (63·9) |
| | | Number censored | 29 (39·7) | 26 (36·1) |
| | | Median (95% CI)[a] | 11·0 (9·0–13·0) | 11·0 (10·0–13·0) |
| | | Hazard ratio (95% CI) | 0·96 (0·63–1·45) | |
| IgG antibody test (central testing) | Positive | Number of events | 1 (100·0) | 0 |
| | | Number censored | 0 | 0 |
| | | Median (95% CI)[a] | 2·0 (n/c–n/c) | n/c (n/c–n/c) |
| | | Hazard ratio (95% CI) | n/c (n/c–n/c) | |
| | Negative | Number of events | 44 (60·3) | 47 (63·5) |
| | | Number censored | 29 (39·7) | 27 (36·5) |
| | | Median (95% CI)[a] | 11·0 (9·0–12·0) | 11·0 (10·0–13·0) |
| | | Hazard ratio (95% CI) | 0·95 (0·63–1·44) | |
| Presence of clinical symptom(s) | Yes | Number of events | 37 (69·8) | 36 (69·2) |
| | | Number censored | 16 (30·2) | 16 (30·8) |
| | | Median (95% CI)[a] | 10·0 (9·0–12·0) | 11·0 (10·0–13·0) |
| | | Hazard ratio | 1·14 (0·72–1·83) | |
| | No | Number of events | 8 (38·1) | 11 (50·0) |
| | | Number censored | 13 (61·9) | 11 (50·0) |
| | | Median (95% CI)[a] | n/c (9·0–n/c) | 12·0 (8·0–n/c) |
| | | Hazard ratio (95% CI) | 0·37 (0·13–1·03) | |

CI, confidence interval; n/c, not calculable; BMI, body mass index; Ig, immunoglobulin
[a]Estimated using the Kaplan–Meier method

### CONSORT 2010 checklist of information to include when reporting a randomised trial*

| Section/Topic | Item No | Checklist item | Reported on page No |
|---|---|---|---|
| **Title and abstract** | | | |
| | 1a | Identification as a randomised trial in the title | 1 |
| | 1b | Structured summary of trial design, methods, results, and conclusions (for specific guidance see CONSORT for abstracts) | 5 |
| **Introduction** | | | |
| Background and objectives | 2a | Scientific background and explanation of rationale | 6-7 |
| | 2b | Specific objectives or hypotheses | 7 |
| **Methods** | | | |
| Trial design | 3a | Description of trial design (such as parallel, factorial) including allocation ratio | 8-9 |
| | 3b | Important changes to methods after trial commencement (such as eligibility criteria), with reasons | Protocol |
| Participants | 4a | Eligibility criteria for participants | 8, Protocol |
| | 4b | Settings and locations where the data were collected | 8 |
| Interventions | 5 | The interventions for each group with sufficient details to allow replication, including how and when they were actually administered | 8-10 |
| Outcomes | 6a | Completely defined pre-specified primary and secondary outcome measures, including how and when they were assessed | 10-11, Protocol |
| | 6b | Any changes to trial outcomes after the trial commenced, with reasons | Protocol |
| Sample size | 7a | How sample size was determined | 11-12, appendix sample size calculation |
| | 7b | When applicable, explanation of any interim analyses and stopping guidelines | 11, Protocol |
| Randomisation: | | | |
| Sequence generation | 8a | Method used to generate the random allocation sequence | 9, Protocol |
| | 8b | Type of randomisation; details of any restriction (such as blocking and block size) | 9, Protocol |
| Allocation concealment mechanism | 9 | Mechanism used to implement the random allocation sequence (such as sequentially numbered containers), describing any steps taken to conceal the sequence until interventions were assigned | 9, Protocol |

| | | | |
|---|---|---|---|
| Implementation | 10 | Who generated the random allocation sequence, who enrolled participants, and who assigned participants to interventions | 9, Protocol |
| Blinding | 11a | If done, who was blinded after assignment to interventions (for example, participants, care providers, those assessing outcomes) and how | 8-13, Protocol |
| | 11b | If relevant, description of the similarity of interventions | 9-10 |
| Statistical methods | 12a | Statistical methods used to compare groups for primary and secondary outcomes | 11-13, Protocol |
| | 12b | Methods for additional analyses, such as subgroup analyses and adjusted analyses | 11-13, Protocol |
| **Results** | | | |
| Participant flow (a diagram is strongly recommended) | 13a | For each group, the numbers of participants who were randomly assigned, received intended treatment, and were analysed for the primary outcome | 15-16, Table S6 |
| | 13b | For each group, losses and exclusions after randomisation, together with reasons | 15, figure 1 |
| Recruitment | 14a | Dates defining the periods of recruitment and follow-up | 15 |
| | 14b | Why the trial ended or was stopped | 15 |
| Baseline data | 15 | A table showing baseline demographic and clinical characteristics for each group | Table 1 |
| Numbers analysed | 16 | For each group, number of participants (denominator) included in each analysis and whether the analysis was by original assigned groups | 15-17, tables, figures |
| Outcomes and estimation | 17a | For each primary and secondary outcome, results for each group, and the estimated effect size and its precision (such as 95% confidence interval) | 15-17, tables, figures |
| | 17b | For binary outcomes, presentation of both absolute and relative effect sizes is recommended | 15-17, tables, figures |
| Ancillary analyses | 18 | Results of any other analyses performed, including subgroup analyses and adjusted analyses, distinguishing pre-specified from exploratory | 16-17, Appendix tables S7-S9 |
| Harms | 19 | All important harms or unintended effects in each group (for specific guidance see CONSORT for harms) | 17, table 3 |
| **Discussion** | | | |
| Limitations | 20 | Trial limitations, addressing sources of potential bias, imprecision, and, if relevant, multiplicity of analyses | 20 |
| Generalisability | 21 | Generalisability (external validity, applicability) of the trial findings | 19-21 |
| Interpretation | 22 | Interpretation consistent with results, balancing benefits and harms, and considering other relevant evidence | 21 |
| **Other information** | | | |
| Registration | 23 | Registration number and name of trial registry | 13 |
| Protocol | 24 | Where the full trial protocol can be accessed, if available | Journal website |
| Funding | 25 | Sources of funding and other support (such as supply of drugs), role of funders | 14 |

*We strongly recommend reading this statement in conjunction with the CONSORT 2010 Explanation and Elaboration for important clarifications on all the items. If relevant, we also recommend reading CONSORT extensions for cluster randomised trials, non-inferiority and equivalence trials, non-pharmacological treatments, herbal interventions, and pragmatic trials. Additional extensions are forthcoming: for those and for up to date references relevant to this checklist, see www.consort-statement.org.
