## Supplementary material for "Phase 3, multicentre, double-blind, randomised, parallel-group, placebo-controlled study of camostat mesilate (FOY-305) for the treatment of COVID-19 (CANDLE study)": Protocol

### FOY-305 Phase III Study

### A Placebo-controlled, Multicenter, Double-blind, Randomized, Parallel-group Comparative Study in SARS-CoV-2 Infection (COVID-19)

### Protocol

#### Ono Pharmaceutical Co., Ltd.

Protocol Number: FOY-305-01

Version Number: 2

Preparation date: 20 November 2020

**Emergency Contact Center (accessible 24 hours a day)**

• Contact center to use if the contact person designated for this clinical study is not available

• Contact in case emergency code breaking is requested

BellSystem24, Inc.

**Serious Adverse Event Report Forms, Pregnancy Report Forms, and Overdose Report Forms should be faxed to:**

Pharmacovigilance Division, Ono Pharmaceutical Co., Ltd.

*For contact for acknowledgment of receipt of Serious Adverse Event Report Forms and Pregnancy Report Forms

Statement on Confidentiality

This protocol is provided only to the head of each study site, investigators, subinvestigators, study collaborators, drug accountability managers, and Institutional Review Boards. Please treat this protocol as a confidential document and do not disclose its content to any third party.

### Ethics

This study will be conducted in compliance with the spirit of the Declaration of Helsinki and in compliance with this Protocol; the standards set forth in Article 14, Paragraph 3 and Article 80-2 of the "Act on Securing Quality, Efficacy and Safety of Pharmaceuticals, Medical Devices, Regenerative and Cellular Therapy Products, Gene Therapy Products, and Cosmetics" (hereinafter referred to as the "PMD Act") and "Guideline for Good Clinical Practice" (International Council for Harmonisation [ICH] E6 Guideline; and "Ministerial Ordinance Concerning Good Clinical Practice" (Ministry of Health and Welfare Ordinance No. 28 of 1997 and related amendments) (hereinafter referred to as "GCP") and related notifications.

Protocol amendment(s) shall be prepared if amendment is considered necessary to ensure that the clinical study is being conducted safely.

### PROTOCOL SYNOPSIS

#### 1 STUDY OBJECTIVES

To assess the efficacy and safety of FOY-305 in patients with SARS-CoV-2 infection (COVID-19) in a placebo-controlled, multicenter, double-blind, randomized, parallel-group comparative study.

#### 2 STUDY DESIGN

This study will be conducted as a multicenter, double-blind, randomized, parallel-group comparative study.

This study consists of the following phases: an observation phase of up to 3 days with no study treatment administered; a double-blind phase of up to 14 days in which subjects will receive double-blinded study treatment; and a follow-up phase of 2 weeks with no study treatment administered. Subjects will be randomized in a 1:1 ratio to receive FOY-305 or placebo in a double-blind fashion through a dynamic allocation procedure, with site, age (≥65 vs <65 years), and underlying disease (chronic respiratory disease, chronic kidney disease, diabetes mellitus, hypertension, cardiovascular diseases, obesity [body mass index; BMI ≥30 kg/m$^2$]) (present vs absent) as allocation factors.

It has been planned that an interim analysis will be performed to determine whether the treatment is effective or ineffective in the subjects randomized as of the time when further enrollment appears to be difficult because of convergence of the COVID-19 pandemic during the subject enrollment period.

#### 3 SUBJECTS

##### 3.1 Subjects

Infectious disease caused by SARS-CoV-2 (COVID-19)

##### 3.2 Inclusion Criteria

Patients will be enrolled only if they have agreed to participate in this study and meet all the following criteria at the time of enrollment:

1. Male or female
2. Aged ≥18 years (at the time of providing consent)
3. Inpatient or outpatient: Inpatient
4. Positive SARS-CoV-2 test by a method eligible for definitive diagnosis (e.g., reverse transcriptase-polymerase chain reaction [RT-PCR] test, loop-mediated isothermal amplification [LAMP] test, antigen test)

5. Enrollment in this study within 5 days after onset of symptoms of SARS-CoV-2 infection (COVID-19) (for asymptomatic patients, enrollment in this study within 5 days after collection of the sample that tested positive)
6. Women of childbearing potential (including patients with amenorrhea due to chemically induced menopause or for other medical reasons)[#1] should agree to use contraceptive method(s)[#2] from the time of consent to 1 month after last administration of the study drug
7. Male subjects should agree to use contraceptive method(s)[#2] from the time of initiation of study drug administration to 1 month after last administration of the study drug

[#1]: Women of childbearing potential include all women who have experienced menarche, have not undergone sterilization (hysterectomy, bilateral tubal ligation, or bilateral oophorectomy), and have not experienced menopause. Menopause is defined as the absence of menses for ≥12 months without any particular reason. Women using oral contraceptives, intrauterine devices, or diaphragms are considered to be of childbearing potential.

[#2]: Dual contraceptive method using 2 of the following methods should be employed: vasectomy or condom for male patient or male partner, and tubal ligation, diaphragm, intrauterine device, or oral contraceptive for female patient or partner.

#### 3.3 Exclusion Criteria

Patients who meet any of the following criteria at enrollment shall be excluded from the study:

1. Receiving oxygen therapy
2. Difficulty in swallowing oral medication
3. History of SARS-CoV-2 infection (COVID-19)
4. Serum potassium >5.5 mEq/L
5. Total bilirubin ≥3.0 mg/dL, or aspartate aminotransferase (AST) (glutamic oxaloacetic transaminase [GOT]) or alanine aminotransferase (ALT) (glutamic pyruvic transaminase [GPT]) ≥2.5 × upper limit of normal of the institution (or ≥100 IU/L)
6. Serum creatinine ≥2.0 mg/dL
7. History of vaccination (including unapproved ones) against SARS-CoV-2 infection (COVID-19)
8. History of treatment with camostat mesilate or nafamostat mesilate for SARS-CoV-2 infection (COVID-19)
9. Taking camostat mesilate
10. History of hypersensitivity to camostat mesilate

11. Patients who are pregnant, breast-feeding, or possibly pregnant, or who wish to be pregnant during the study
12. History of treatment specified as prohibited therapy or concomitant therapy before initiation of the study treatment
13. History of serious drug allergy
14. Judged as lacking the ability to consent, for example, because of comorbid dementia
15. Judged by the investigator or subinvestigator as inappropriate as a study subject for any other reason

- The eligibility of patients should be assessed on the basis of the information from the observation period.
- For items 4, 5, and 6, if multiple actual values are available from the observation period, use the value obtained on the day closest to the day of enrollment for judgment.

### 4 DOSAGE AND ADMINISTRATION AND DURATION OF TREATMENT

FOY-305 600 mg (six 100-mg tablets) or placebo (6 placebo tablets) will be orally administered 4 times daily, before breakfast, before lunch, before evening meal, and at bedtime. Patients should be fasted for at least 1 hour after administration. The treatment period is up to 14 days. Patients will discontinue the study treatment when tested negative for SARS-CoV-2 on 2 consecutive occasions (at an interval of ≥24 hours). Once the eligibility is confirmed, the study treatment may be initiated from the earliest dosing time point mentioned above. Although the treatment period is up to 14 days, patients taking the first dose before lunch or before evening meal on the first day will receive the study treatment over 15 days at maximum (up to 56 doses) following the specified dosing regimen thereafter.

### 5 TREATMENTS AND CONCOMITANT THERAPIES

#### 5.1 Prohibited Treatments and Concomitant Therapies

1. Drugs for the treatment of SARS-CoV-2 infection (COVID-19)

   Use of the following drugs is prohibited from the day of onset of symptoms of SARS-CoV-2 infection (COVID-19) (for asymptomatic patients, the day of collection of the sample that tested positive) until completion of the study treatment: However, continued use of the drug for a comorbidity is permitted (in principle, to be continued with the same dosing regimen during the period from the informed consent to completion of the study treatment).

   Drugs with antiviral effects (remdesivir, favipiravir, ciclesonide, nafamostat mesilate, hydroxychloroquine, ivermectin, combination drug of lopinavir and ritonavir, povidone-iodine [e.g., used in the oral and nasal cavities]) and drugs with anticytokine effects (e.g., tocilizumab, Janus kinase [JAK] inhibitors)

2. Any other unapproved drugs not intended for the treatment of SARS-CoV-2 infection (COVID-19) (including those administered in clinical studies, unapproved combination drugs, and drugs with new dosage form)

   Use of any other unapproved drugs is prohibited from 28 days before initiation of the study treatment to completion of the study treatment.

3. Camostat mesilate in the market

   Use of camostat mesilate in the market is prohibited from initiation of the study treatment to completion of the study treatment.

#### 5.2 PERMITTED THERAPIES

Of the drugs for the treatment of SARS-CoV-2 infection (COVID-19), those that do not fall under the category of 5.1 Prohibited Treatments and Concomitant Therapies will be permitted (e.g., antipyretics, antibiotics, corticosteroids, oxygen therapy).

### 6 CRITERIA FOR DISCONTINUATION OF STUDY TREATMENT

1. The subject has requested treatment discontinuation
2. The legally acceptable representative has requested treatment discontinuation
3. The subject is found not to meet the inclusion criteria or to meet the exclusion criteria
4. Continuation of the study is difficult because of adverse event(s), whether or not related to the study drug, in the opinion of the investigator or subinvestigator
5. The subject tests negative for SARS-CoV-2 on 2 consecutive occasions
6. When both of the criteria a. and b. are met, and continuation of the study is considered difficult:
   a. Exacerbation of pneumonia is observed by chest imaging
   b. Oxygen saturation ($SpO_2$) is ≤93% despite oxygen therapy
7. The subject receives any prohibited concomitant medication
8. The subject is dead
9. The subject is found to be pregnant
10. Continuation of the study is not appropriate for any other reasons, in the opinion of the investigator or subinvestigator

### 7 CRITERIA FOR DISCONTINUATION OF THE STUDY

1. The subject has requested for withdrawal from the study
2. The legally acceptable representative has requested for withdrawal of the subject from the study

3. Continuation of the study is difficult because of adverse event(s), whether or not related to the study drug, in the opinion of the investigator or subinvestigator
4. The subject is dead
5. The subject is found to be pregnant
6. Continuation of the study is not appropriate for any other reasons, in the opinion of the investigator or subinvestigator

### 8 STUDY SCHEDULE AND OBSERVATION ITEMS

**Table 8-1 Study schedule**

| Time | Observation phase | Treatment phase | | | | | | | | | | | | | | | Follow-up phase[10] |
|---|---|---|---|---|---|---|---|---|---|---|---|---|---|---|---|---|---|
| Day | | Day 1 | Day 2 | Day 3 | Day 4 | Day 5 | Day 6 | Day 7 | Day 8 | Day 9 | Day 10 | Day 11 | Day 12 | Day 13 | Day 14 | Treatment discontinuation[9] | 14 days after last dose (±3) |
| Reference day (acceptable window) | Day -1[4] (-3 to 1) | Day 1 | Day 2 | Day 3 | Day 4 | Day 5 | Day 6 | Day 7 | Day 8 | Day 9 | Day 10 | Day 11 | Day 12 | Day 13 | Day 14 | Treatment discontinuation (+1) | |
| Informed consent | X | | | | | | | | | | | | | | | | |
| Patient demographics | X | | | | | | | | | | | | | | | | |
| History/complication | X | | | | | | | | | | | | | | | | |
| Inclusion/exclusion criteria | X | | | | | | | | | | | | | | | | |
| Laboratory test[1] | | X[5] | | | | | | X[6] | | | | | | | X[7] | X | |
| Patient enrollment | | X | | | | | | | | | | | | | | | |
| Vital signs[1] | | X[5] | X | X | X | X | X | X | X | X | X | X | X | X | X[7] | X | |
| 12-lead ECG[1] | | X[5] | | | | | | X[6] | | | | | | | X[7] | X | |
| SARS-CoV-2 test[1] | | X[5] | X | X | X | X | X | X | X | X | X | X | X | X | X[8] | X | |
| Ordinal scale for severity[1] | | X[5] | X | X | X | X | X | X | X | X | X | X | X | X | X | X | |
| Presence of clinical manifestation[1] | | X[5] | X | X | X | X | X | X | X | X | X | X | X | X | X | X | |
| Chest imaging[1] | | X[5] | | | | | | | | | | | | | X[6] | X | |
| Antibody test[1] | | X[5] | | | | | | | | | | | | | X[6] | X | |
| Study drug administration[2] | | ◄ | — | — | — | — | — | — | — | — | — | — | — | — | ► X | | |
| Survival status | | X ◄ | — | — | — | — | — | — | — | — | — | — | — | — | ► X | X | |
| Use of mechanical ventilation | | X ◄ | — | — | — | — | — | — | — | — | — | — | — | — | ► X | X | |
| Adverse events | | X ◄ | — | — | — | — | — | — | — | — | — | — | — | — | — | — | ► X |
| Concomitant medication/therapy[3] | | X ◄ | — | — | — | — | — | — | — | — | — | — | — | — | — | — | ► X |

1) These assessments will be performed until the subject tests negative for SARS-CoV-2 on 2 consecutive occasions. Each test for SARS-CoV-2 should be separated by at least 24 hours, except for Days 1 to 2.
2) Once the eligibility is confirmed, the study treatment may be initiated. The treatment period is up to 14 days. Patients will discontinue the study treatment when tested negative for SARS-CoV-2 on 2 consecutive occasions. Patients taking the first dose before lunch or before evening meal on the first day will receive the study treatment over 15 days at maximum (up to 56 doses) following the specified dosing regimen thereafter. If the subject cannot start the study treatment on the day of enrollment, he/she will start the study treatment before breakfast on the day after enrollment as far as possible. If the dates of enrollment and treatment initiation are not the same, the date of enrollment will be used as Day 1.
3) From completion of the tests (other than the follow-up for adverse events) specified for Day 14 (or discontinuation of treatment) until 14 days after the last dose of the study treatment, only the concomitant medications and therapies used for adverse events should be collected.
4) To be completed within 3 days before enrollment.
5) To be performed within 24 hours before enrollment.
6) An acceptable window of ±1 day is included for laboratory tests and 12-lead ECG on Day 7, and chest imaging and antibody test on Day 14.
7) Laboratory tests, vital sign measurement, and 12-lead ECG should be performed after completion of the last dose of the study treatment, with an acceptable window of +1 day, starting at the time of last administration of the study drug.
8) If the patient tests negative for SARS-CoV-2 for the first time on Day 14, the patient should be re-tested for SARS-CoV-2 after an interval of at least 24 hours to confirm the negative SARS-CoV-2 test result.
9) The procedures should be performed after the study treatment is discontinued. For the SARS-CoV-2 tests, ordinal scale for severity, and clinical symptom assessment, however, results obtained on the same day before the study treatment is discontinued may be used. The study treatment should be discontinued when the patient tests negative for SARS-CoV-2 on 2 consecutive occasions as well as when any of the criteria for discontinuation of the study treatment is met. In such cases, the tests specified for the time of study treatment discontinuation should be performed.
10) During the study period, the patient may be discharged from the institution if there is no safety issue as determined by the investigator or subinvestigator. For patients discharged from the institution, the follow-up phase assessment may be performed via telephone.

### 9 ENDPOINTS

The efficacy analysis of this study will include the modified Intention-to-Treat set (mITT), consisting of subjects who have been randomized and tested positive for SARS-CoV-2 (assessed by the local laboratory) on Day 1. The safety analysis of this study will include the Safety Set (SAF) consisting of subjects who have received at least 1 dose of the study treatment.

#### 9.1 Efficacy Endpoint

##### 9.1.1 Primary Endpoint

###### 9.1.1.1 Primary Endpoint

Time to negative test for SARS-CoV-2 as assessed by the local laboratory

###### 9.1.1.2 Analysis Method

Using a stratified Cox proportional hazard model with age and underlying disease (chronic respiratory disease, chronic kidney disease, diabetes mellitus, hypertension, cardiovascular disease, obesity [BMI $\geq$30 kg/m$^2$]) as allocation factors and the SARS-CoV-2 viral load on Day 1 as the stratification factor, the posterior mean hazard ratio of the FOY-305 group to the placebo group will be calculated along with the two-sided 95% confidence interval. In addition, the Bayesian posterior probability for hazard ratio >1.0 and the Bayesian posterior probability for hazard ratio <1.4 (only for the interim analysis) will be calculated. The prior distribution of the regression coefficient is assumed to be uniform distribution.

##### 9.1.2 Secondary Endpoints

1. Time to negative test for SARS-CoV-2 as assessed by the central laboratory
2. Proportion of subjects who test negative for SARS-CoV-2 (as assessed by the local and central laboratories)
3. Ordinal scale for severity
4. Proportion of subjects on mechanical ventilator
5. Survival status

##### 9.1.3 Exploratory Endpoints

1. SARS-CoV-2 viral load (as measured by the central laboratory)
2. Presence or absence of lung lesion as observed by chest imaging

3. Time to resolution of clinical symptoms and proportion of subjects in whom clinical symptoms have resolved
4. Antibody response (IgM and IgG) (measured by the central laboratory)

##### 9.2 Safety Endpoints

1. Adverse events
2. Laboratory tests (hematology, blood biochemistry, qualitative urinalysis)
3. Vital signs (body temperature, $SpO_2$, systolic/diastolic blood pressure, pulse rate)
4. 12-lead electrocardiogram (ECG)

#### 10 PLANNED NUMBER OF SUBJECTS

110 subjects in total, with 55 per group

#### 11 PLANNED STUDY PERIOD

August 2020 to October 2021

#### Tables

Appendix 1 Ordinal Scale
Appendix 2 Severity of Adverse Events
Supplementary Volume 1 Study Organization

**Table 1 List of Abbreviations**

| Abbreviation | Term |
|---|---|
| ALP | Alkaline phosphatase |
| ALT (GPT) | Alanine aminotransferase (glutamic pyruvic transaminase) |
| AST (GOT) | Aspartate aminotransferase (glutamic oxaloacetic transaminase) |
| BUN | Blood urea nitrogen |
| CK (CPK) | Creatine kinase (creatine phosphokinase) |
| COVID-19 | SARS-CoV-2 infection |
| CT | Computed tomography |
| eCRF | Electronic case report form |
| EC50 | Half-maximal effective concentration |
| GCP | Good Clinical Practice |
| IP-GMP | Good Manufacturing Practice for Investigational Products |
| γ-GTP | Gamma-glutamyl transpeptidase |
| ICH | International Council for Harmonisation of Technical Requirements for Pharmaceuticals for Human Use |
| IDMC | Independent data monitoring committee |
| LDH | Lactate dehydrogenase |
| MERS | Middle East respiratory syndrome |
| mITT | Modified intention-to-treat |
| pH | Hydrogen ion exponent |
| PT | Preferred term |
| RNA | Ribonucleic acid |
| S protein | Spike protein |
| SAF | Safety set |
| SARS | Severe acute respiratory syndrome |
| SARS-CoV-2 | SARS coronavirus 2 |
| SARS-2-S | SARS-CoV-2-S protein |
| $SpO_2$ | Oxygen saturation |
| TMPRSS2 | Transmembrane serine protease 2 |

**Table 2 List of Terms**

| Term | Definition |
|---|---|
| Legally acceptable representative | In the context of participation in a clinical study, a person who can acknowledge, together with or in place of the subject, that consent was obtained properly when the subject is not fully able to provide consent. The legally acceptable representative may be a person with parental rights, a spouse, conservator, or the like and must consider the physical and emotional quality of life of both subject and legally acceptable representative and act in the best interests of the subject. |
| Impartial witness | A person who is independent from the clinical study, who cannot be unfairly influenced by individuals involved in the clinical study, and who attends the informed consent process. Investigators, subinvestigators, and clinical study collaborators are not qualified as impartial witnesses. |

### 1 BACKGROUND OF DEVELOPMENT

#### 1.1 Introduction

FOY-305 (non-proprietary name, camostat mesilate; trade name, Foipan Tablets) is an oral protease enzyme inhibitor developed by Ono Pharmaceutical Co., Ltd. FOY-305 is a serine protease inhibitor that shows inhibitory effects on trypsin, plasma kallikrein, plasmin, thrombin, $C_1$r, and $C_1$ esterase but not on α-chymotrypsin, pepsin, and pancreatin, among others. It is a low molecular weight compound represented by the chemical name dimethylcarbamoylmethyl 4-(4-guanidinobenzoyloxy) phenylacetate monomethanesulfonate. FOY-305 was approved for manufacturing on 31 January 1985 for the alleviation of acute symptoms associated with chronic pancreatitis on the basis of the results from a Japanese double-blind controlled study that demonstrated the usefulness of the drug in alleviating acute symptoms and improving amylase levels in the blood and urine of patients with chronic pancreatitis. The efficacy and safety of FOY-305 were established through the subsequent post-marketing surveillance, and the re-examination was completed on 03 June 1992. An additional indication of postoperative reflux esophagitis was approved on 01 July 1994, for which re-examination was completed on 17 January 2003.

SARS-CoV-2 (SARS coronavirus 2) infection (COVID-19) is an infectious disease caused by the novel coronavirus that was first identified in December 2019 in Wuhan, Hubei Province, the People's Republic of China. On 30 January 2020, the World Health Organization (WHO) declared a Public Health Emergency of International Concern (PHEIC) for COVID-19. In response to the subsequent situations, such as rapid global expansion of the infection and severity of the symptoms, the WHO announced on 11 March 2020 that the COVID-19 outbreak had turned into a pandemic, positioning COVID-19 as a threat to the entire human race.[1)] As of 15:00 on 29 July 2020, 16,643,683 and 659,079 people in the world had been infected and died, respectively, and 190 countries/regions were affected by the pandemic.[2)] In Japan, a total of 57,766 people have reportedly been affected (tested positive) by COVID-19, including 1143 deaths, as of 19 August 2020.[3)] COVID-19 is clinically characterized by an incubation period of 5 to 6 days on average (range: 1 to 14 days),[4)] followed by flu-like symptoms such as pyrexia, respiratory symptoms including cough, and general malaise, lasting around 1 week. Patients in whom the disease progresses further present with systemic inflammatory reactions due to a cytokine storm and life-threatening, serious symptoms such as dyspnea, with typical features of severe acute pneumonia observed by imaging, such as chest X-ray and computed tomography (CT) scan.

Viruses need to infect a host cell for replicating the viral genome by utilizing the gene replication mechanism of the host as they lack the ability of self-propagation. One of the known mechanisms of host cell infection is as follows: the spike protein on the viral surface selects the target cell for infection by recognizing a specific molecule on the host cell membrane as a receptor, following which the conformation of the spike protein is modified by the host cell, leading to activation of the infectivity.

SARS-CoV-2, a type of coronavirus classified as a positive-sense single-stranded RNA virus, has a spike protein (S protein) that binds to angiotensin-converting enzyme II (ACE2) on the host cell membrane as a functional receptor. Subsequently, the S protein is cleaved by the host-derived protease activity into S1 and S2: the S1 fragment binds to ACE2, while the S2 fragment is cleaved by transmembrane serine protease 2 (TMPRSS2) on the host cell membrane, facilitating fusion of the viral envelope (outer membrane) with the cell membrane.[5)] As such, TMPRSS2 plays an important role in viral entry into a host cell. TMPRSS2 is known to cleave the S protein not only in SARS-CoV-2 but also in the severe acute respiratory syndrome (SARS) and Middle East respiratory syndrome (MERS) viruses.[6)]

Given the clinical characteristics of COVID-19 and the propagation/replication mechanism of the SARS-CoV-2 virus, various therapeutic approaches are being tested for the treatment of COVID-19, including controlling the cytokine storm and controlling viral genome replication by RNA polymerase. In Japan, remdesivir, an RNA polymerase inhibitor, has been granted a fast-track approval for the indication of SARS-CoV-2 infection. Use of dexamethasone, a corticosteroid, is also recommended for patients requiring mechanical ventilation or oxygen administration.[7)]

It has been reported that FOY-305, a serine protease inhibitor, can control infections with viruses, including the coronaviruses such as SARS-CoV and MERS-CoV and the influenza viruses, by inhibiting cleavage of the spike protein by the host enzyme.[8)-10)] In February 2020, Hoffmann et al.[5)] from the German Primate Center reported that ACE2 and TMPRSS2 on the airway epithelial cells are essential for SARS-CoV-2 infection. Furthermore, they discovered that FOY-305 (camostat mesilate) inhibits the effect of TMPRSS2, thereby blocking SARS-CoV-2 from infecting a human airway epithelial cell-derived cell line (Calu-3 cell).

In addition, a Phase I repeat oral dose study in Japanese healthy adult males demonstrated favorable tolerability of FOY-305 when administered at doses up to 600 mg given 4 times daily for 7 days.

On the basis of these findings, Ono Pharmaceutical Co., Ltd. has planned a Japanese Phase III study of FOY-305, anticipating that FOY-305 can fulfill the purpose of treatment for COVID-19 by exerting its protease inhibitory effect on TMPRSS2.

#### 1.2 Summary of Important Findings from Preclinical and Clinical Studies

##### 1.2.1 Summary of Preclinical Data

###### Efficacy Pharmacology

Various *in vitro*, *ex vivo* and *in vivo* studies revealed that FOY-305 inhibited trypsin activity as well as various serine proteases of kinin-generating, fibrinolytic, coagulation, and complement cascades in living organisms, and showed anti-inflammatory activity in experimental pancreatitis and reflux esophagitis models. *In vitro* studies, FOY-305 also exhibited antiviral activity against severe acute respiratory syndrome coronavirus 2 (SARS-CoV-2), and prevented the virus from entering the human airway epithelial cell-derived cell line (Calu-3 cells). In an *in vivo* mouse infection experiment using SARS-CoV, which is known, like SARS-CoV-2, to be transmitted via TMPRSS2 activity of the host cell, oral administration of FOY-305 improved the survival rate.

###### Safety Pharmacology

Safety pharmacology studies that investigated the effects of oral administration of FOY-305 on the central nervous system showed that in mice oral FOY-305 was associated with decreased spontaneous locomotor activity at doses of 1000 mg/kg and higher and respiratory distress and overreaction to external stimuli at doses of 3000 mg/kg and higher, and in rats respiratory distress, decreased spontaneous locomotor activity, overreaction to external stimuli, convulsions, loss of righting reflex, and dyspnoea at doses of 3000 mg/kg and higher. FOY-305 showed no effect on the body temperature or autonomic nervous system. The investigation of the effects of FOY-305 on the cardiovascular and respiratory systems showed that oral administration of the drug had no effect on the blood pressure in rats at doses up to 1000 mg/kg. Intravenous administration of FOY-305 to dogs was associated with transiently increased blood pressure at 0.1 to 30 mg/kg, and increased blood pressure that then dropped to a level lower than the baseline value at 10 and 30 mg/kg. At 100 mg/kg, only a remarkable decrease in blood pressure was observed. At 0.1 mg/kg and higher, breath excitation or transient respiratory depression following excitation was observed, and a decrease in heart rate at 1 mg/kg and higher. No effect was observed on ECG QT interval at doses up to 30 mg/kg. The investigation of the effects of oral administration of FOY-305 on the urinary system in rats revealed that the drug was associated with increased sodium ion excretion at 100 mg/kg and higher and decreased urine output at 1000 mg/kg. The investigation of the effects of FOY-305 on the smooth muscle tissues isolated from various organs showed an inhibitory action on smooth muscle tissue contraction isolated from the gastrointestinal tract, blood vessels, and uterus.

The no-observed-effect dose level for central nervous toxicity of oral FOY-305 was 500 and 1500 mg/kg in mice and rats, respectively. The no-observed-effect dose level for cardiovascular and respiratory toxicities of oral FOY-305 (rats) and intravenous FOY-305 (dogs) was 1000 and 0.03 mg/kg in rats and dogs, respectively. The no-observed-effect dose level of oral FOY-305 for urinary toxicity was 30 mg/kg in rats.

##### Pharmacokinetics

Using male rats with a gastrointestinal loop, $^{14}$C-FOY-305 was infused into the loop at 2 mg/kg, and the radioactivity concentration in portal vein plasma and the in-loop residual rate were measured. The results showed that the radioactivity was absorbed throughout the duodenum and the upper large intestine, with 40% or more of the dose being absorbed. The protein binding rate of FOY-305 was 25.8% to 28.2% in human serum and 50.2% to 55.3% in dog and rat serum. The concentrations of radioactivity in various tissues of male rats treated with $^{14}$C-FOY-305 reached the maximum level at 1 hour post-dose, except for the blood and spleen, showing a particularly greater distribution in the liver, kidney, and bladder. The concentrations of radioactivity in various tissues at 72 hours post-dose were 0.01% or lower of the dose. Similarly, the concentrations of radioactivity in various tissues were 0.003% of the dose 72 hours after the final dose of 7-day once daily repeated oral administration. After administration of $^{14}$C-FOY-305 to pregnant and breastfeeding rats, a small amount of radioactivity penetrated the placenta and was transferred to milk. FOY-305 was rapidly hydrolyzed *in vivo*, and was below the lower limit of quantitation in human plasma. Based on the abundance ratio in plasma and inhibitory effect on protease enzyme, it was likely that FOY-251, an active metabolite of FOY-305, played a major role in the efficacy of FOY-305 *in vivo*. It was suggested that carboxylesterase and arylesterase were mainly involved in the hydrolysis of FOY-305 and FOY-251, respectively. Neither FOY-305 nor FOY-251 inhibited CYP1A2, CYP2C9, CYP2C19, CYP2D6, or CYP3A4.

##### Toxicity

In single-dose toxicity studies of FOY-305, the drug was orally administered to male and female mice and rats. The results showed that the approximate lethal dose was 3630 and 3300 mg/kg in male and female mice, respectively, and 3300 mg/kg in both male and female rats. With regard to dead mice and rats, respiratory distress and convulsions were observed at 10 minutes post-dose and thereafter, and many of the animals died within 1 hour post-dose. In repeated-dose toxicity studies of FOY-305, the drug was administered to rats and dogs for up to 180 and 30 days, respectively. In the results, suppressed weight gain, decreased or increased food consumption, gastrointestinal toxicities (suppressed digestion/absorption, vomiting, and gastrointestinal damage) were observed at 1300 mg/kg and higher in rats and at 300 mg/kg in dogs, and death occurred at 3000 and 300 mg/kg in rats and dogs, respectively. The no-observed-adverse-effect level was 550 and 100 mg/kg in rats and dogs, respectively.

Bacterial reverse mutation and Rec assays that investigated the potential genotoxicity of FOY-305 showed that FOY-305 had no genotoxicity.

In reproductive and developmental toxicity studies of FOY-305, in which FOY-305 was orally administered to rats before pregnancy and in early pregnancy, rats and rabbits during organogenesis, and perinatal and breastfeeding rats, suppressed fetal weight gain was observed at 400 mg/kg and higher in perinatal and breastfeeding rats.

FOY-305 showed no antigenicity or local irritation.

##### 1.2.2 Summary of Clinical Study Data

In clinical studies, including a double-blind controlled study in patients with chronic pancreatitis, an improvement was observed in symptoms, such as pain, tenderness, and serum and urine amylase levels, with an efficacy rate of 48.6% (155/319 patients). In clinical studies, including a double-blind controlled study in patients with postoperative reflux esophagitis, an endoscopic improvement was observed in erosion and haemorrhage, and an improvement was observed in subjective symptoms, such as heartburn, chest pain, and chest burning feeling, with an efficacy rate of 82.0% (132/161 patients).

As for the indication of the remission of acute symptoms in chronic pancreatitis, pre-approval investigations and post-marketing surveillance showed that the incidence of adverse drug reactions (including abnormal laboratory tests) was 1.8% (69/3806 patients , 83 reactions), and common adverse drug reactions included rash (0.3%, 13 reactions), pruritus (0.2%, 9 reactions), nausea (feeling queasy) (0.3%, 10 reactions), abdominal discomfort (0.1%, 4 reactions), and abdominal distension (0.2%, 6 reactions) (at the end of reexamination). As for the indication of postoperative reflux esophagitis, the incidence of adverse drug reactions (including abnormal laboratory tests) was 1.3% (57/4224 patients , 75 reactions), and common adverse drug reactions included hepatic function abnormal (0.1%, 5 reactions), AST (GOT) increased (0.1%, 4 reactions), diarrhoea (0.2%, 8 reactions), and nausea (feeling queasy) (0.1%, 5 reactions) (at the end of reexamination).

Clinically significant adverse drug reactions were (1) shock and anaphylaxis, (2) platelets decreased, (3) hepatic impairment and jaundice, and (4) hyperkalaemia.

In a clinical study of repeated doses of FOY-305 600 mg given 4 times a day to healthy adults, adverse drug reactions of hyperuricaemia and aphthous ulcer were observed in 1 each of 14 subjects. All the adverse drug reactions were Grade 1, and rapidly resolved after the end of the treatment period, with no action taken with the study drug. Plasma FOY-251 concentrations were lower after dosing in the fed state compared with dosing in the fasting state, but there were no major differences between dosing at 60 minutes before a meal and dosing in the fasting state.

###### 1.2.2.1 A Multiple-dose Study in Healthy Japanese Adults (Study FOY-305-02)

A total of 14 healthy Japanese adult males (7 subjects/cohort) received FOY-305 600 mg 4 times a day on Day 1 and Days 3 to 9 (for 7 days). Day 2 was a treatment-free day. Subjects in Cohort 1 received treatment after a prescribed (standard) meal in the morning, at noon, and in the evening (and in the fasting state in the morning on Day 1 only) and at a specified time at night. Subjects in Cohort 2 received treatment before a prescribed (standard) meal in the morning, at noon, and in the evening (30 and 60 minutes before starting the meal on Day 1 and Days 3 to 9, respectively) and at a specified time at night.

Pharmacokinetics

Plasma FOY-251 concentrations were lower after dosing in the fed state compared with dosing in the fasting state, but there were no major differences between dosing at 60 minutes before a meal and dosing in the fasting state.

Safety and Tolerability

In Cohort 1, Grade 1 hyperuricaemia was observed in 1/7 subjects (14.3%), and the event was considered to be an adverse drug reaction. In Cohort 2, Grade 1 aphthous ulcer was observed in 1/7 subjects (14.3%), and the event was considered to be an adverse drug reaction. There was no death, serious adverse event, or adverse event leading to treatment discontinuation/treatment interruption/dose reduction.

##### 1.2.3 Summary of Known and Potential Risks and Benefits to Subjects

According to the package insert for FOY-305 (version 1, revised in November 2019) and Interview Form (version 6, revised in December 2019), the approved indications, dosages, precautions, etc. are provided below. The information below is based on data from previous nonclinical and clinical studies and post-marketing surveillance of FOY-305, and should be taken into account in regard to the conduct of the study.

###### 1.2.3.1 Indications

- Remission of acute symptoms in chronic pancreatitis
- Postoperative reflux esophagitis

##### 1.2.3.2 Precautions for Indications

| **5. Precautions for Indications**<br>**<Remission of Acute Symptoms in Chronic Pancreatitis>**<br>**5.1** FOY-305 should not be administered to patients with severe chronic pancreatitis requiring gastric juice suction or dietary restriction (e.g., no food or liquid).<br>**<Postoperative Reflux Esophagitis>**<br>**5.2** FOY-305 should not be used for postoperative reflux esophagitis caused by gastric juice reflux, because it is unlikely that FOY-305 is beneficial. |
|---|

**(Rationale)**

**5.1** The precaution is given because basic treatment for patients with severe chronic pancreatitis includes gastric juice suction and restriction with no food or liquid to prevent the stimulation of pancreatic exocrine function, and the use of oral agents itself can be a physical stimulus that may promote the secretion of pancreatic juice.

**5.2** The precaution is given because although the reflux of gastric juice may be mainly attributed to a small number of patients with postgastrectomy reflux esophagitis, postoperative reflux esophagitis can also be caused by the reflux of alkaline digestive juice in patients receiving subtotal gastrectomy as well as in those who had total gastrectomy.

##### 1.2.3.3 Dosage and Administration

###### 1.2.3.3.1 Description of Dosage and Administration

**<Remission of Acute Symptoms in Chronic Pancreatitis>**

In general, camostat mesilate should be orally administered at a daily dose of 600 mg in three divided doses. The dose should be increased or reduced based on symptoms.

**<Postoperative Reflux Esophagitis>**

In general, camostat mesilate should be orally administered after a meal at a daily dose of 300 mg in three divided doses.

###### 1.2.3.3.2 Background and Rationale of the Dosage and Administration

**<Remission of Acute Symptoms in Chronic Pancreatitis>**

Based on results of a Phase IIa study in chronic pancreatitis, in which patients received FOY-305 at a daily dose of 300 mg or 600 mg, 600 mg/day was deemed more appropriate.

**<Postoperative Reflux Esophagitis>**

A Phase II double-blind controlled study [1] indicated the need to test a daily dose of 300 mg or lower as the optimal dose of FOY-305, and based on results of a Phase II double-blind controlled study [2] that compared the 300 mg/day and 90 mg/day groups, 300 mg/day was deemed more appropriate.

##### 1.2.3.4 Precautions for Dosage and Administration and Reasons

None

##### 1.2.3.5 Warnings and Reasons

None

##### 1.2.3.6 Contraindication and Reason

| |
|---|
| **2. Contraindications (FOY-305 should not be used in patients with any of the following conditions.)**<br>Having a history of hypersensitivity to any ingredient of FOY-305 |

**(Rationale)**

FOY-305 has not been associated with the production of specific antibodies in antigenicity studies in animals; however, the contraindication was selected because adverse drug reactions of shock and anaphylaxis have been reported.

##### 1.2.3.7 Important Precautions and Reasons

| |
|---|
| **8. Important Precautions**<br>**<For Both Indications>**<br>**8.1** Serious hyperkalaemia may occur. Serum electrolytes should be measured.<br>**<Postoperative Reflux Esophagitis>**<br>**8.2** If there is no symptomatic improvement, FOY-305 should not be used for a long period of time without a specific purpose. |

**(Rationale)**

**8.1** (See Section 1.2.3.10.1, "Clinically Significant Adverse Drug Reactions and Their Early Symptoms.")

**8.2** For patients receiving FOY-305 for postoperative reflux esophagitis, a decision should be made regarding whether to continue or discontinue treatment with FOY-305 based on the patient's symptoms. The precaution was selected for the following reason: the duration of treatment with FOY-305 in double-blind controlled studies was 8 weeks; therefore, if the patient shows no response to FOY-305 after 8 weeks of treatment, the use of FOY-305 should not be continued without a specific purpose, and switching to alternative therapy needs to be considered.

##### 1.2.3.8 Precautions for Patients with Certain Characteristics

###### 1.2.3.8.1 Patients Having a Concurrent Illness/Past History

| |
|---|
| **9. Precautions for Patients with Certain Characteristics**<br>**9.1 Patients Having a Concurrent Illness/Past History**<br>**9.1.1 Patients with Hypersensitivity**<br>Adverse drug reactions are likely to occur. |

**(Rationale)**

A special investigation on the safety and efficacy of the long-term use of FOY-305 in patients with postoperative reflux esophagitis showed that the incidence of adverse drug reactions by patient characteristic was higher in patients with predisposition to hypersensitivity (40%, 6/15 patients) than in patients with no predisposition to hypersensitivity (1.42%, 8/565 patients). Accordingly, the findings were reflected in the precautions (see Section 1.2.3.10.5, "Frequencies of Adverse Drug Reactions by Patient Characteristic (Extracted) <Remission of Acute Symptoms in Chronic Pancreatitis>").

##### 1.2.3.8.2 Patients with Renal Impairment

None

##### 1.2.3.8.3 Patients with Hepatic Impairment

None

##### 1.2.3.8.4 Patients of Reproductive Potential

None

##### 1.2.3.8.5 Pregnant Women

| **9.5 Pregnant Women**<br>The use of FOY-305 in women who are pregnant or possibly pregnant should be limited to cases in which treatment benefits are considered to outweigh the risks. The use of a high dose should be avoided. In an animal (rat) study, in which FOY-305 was administered at 40 times the clinical dose (400 mg/kg/day) or higher, suppressed fetal weight gain was reported.[11)] |
|---|

**(Rationale)**

The precaution was selected for the following reason: in an animal study, suppressed fetal weight gain was observed after administrating FOY-305 to rat dams at a dose of 400 mg/kg/day, although FOY-305 showed low fetal distribution, with no teratogenicity or fetal death. In a drug use investigation in patients with chronic pancreatitis and drug use and special investigations in patients with postoperative reflux esophagitis, there was no report on the use of FOY-305 in pregnant women.

##### 1.2.3.8.6 Breastfeeding Women

None

##### 1.2.3.8.7 Children

| **9.7 Children**<br>No clinical study of FOY-305 in children has been conducted. |
|---|

**(Rationale)**

The precaution was selected because the number of children (aged under 15 years) treated with FOY-305 is limited, and the safety of FOY-305 in children has not been established. A drug use investigation in patients with chronic pancreatitis revealed that there were 6 children treated with FOY-305, none of whom had any adverse drug reaction. Drug use and special investigations in patients with postoperative reflux esophagitis revealed that there was no child treated with FOY-305.

###### 1.2.3.8.8 Elderly Patients

None

**(Rationale)**

A Use in Elderly Patients section had been included, but it was then removed because a drug use investigation in patients with chronic pancreatitis and drug use and special investigations in patients with postoperative reflux esophagitis showed that the safety and efficacy of FOY-305 did not differ between elderly and non-elderly patients.

##### 1.2.3.9 Interactions

###### 1.2.3.9.1 Contraindications for Coadministration and Reasons

None

###### 1.2.3.9.2 Precautions for Coadministration and Reasons

None

##### 1.2.3.10 Adverse Drug Reactions

**11. Adverse Drug Reactions**
The adverse drug reactions below may occur. Patients should be duly monitored, and appropriate actions (e.g., treatment discontinuation) should be taken if any abnormality is noted.

###### 1.2.3.10.1 Clinically Significant Adverse Drug Reactions and Their Early Symptoms

**11.1 Clinically Significant Adverse Drug Reactions**
**11.1.1 Shock and Anaphylaxis** (frequency unknown for both)
In the event of decreased blood pressure, dyspnoea, feeling itchy, etc., the use of FOY-305 should be discontinued, and appropriate treatment should be provided.
**11.1.2 Decreased Platelets** (frequency unknown)
**11.1.3 Hepatic Impairment and Jaundice** (frequency unknown for both)
Hepatic impairment and jaundice with remarkable increases in AST, ALT, γ-GTP, and Al-P may occur.
**11.1.4 Hyperkalaemia** (frequency unknown)
Serious hyperkalaemia may occur.

**(Rationale)**

**11.1.1 Shock and Anaphylaxis**

In the post-marketing setting, shock and anaphylaxis have been reported. FOY-305 has not been associated with the production of specific antibodies in antigenicity studies in animals. However, the time course of manifestations (onset within 30 minutes after taking the drug in most patients), the earliest symptoms (e.g., skin symptoms including welts, feeling itchy, and erythema, dizziness/feeling bad/headache dull, sweating/cold sweat, oedema/swollenness, dyspnoea, and decreased blood pressure), and a past history of hypersensitivity symptoms (e.g., urticaria, feeling itchy, oedema) that occurred after taking FOY-305 in some patients suggest that these symptoms are related to anaphylactic reactions. There is no way to predict the occurrence of FOY-305-related shock and other events. Therefore, the past history (especially a past history of FOY-305-related allergy) should be carefully taken in advance, and the use of FOY-305 should be discontinued and appropriate treatment for shock (e.g., vasopressors and steroids) should be provided if such signs or symptoms are observed.

**11.1.2 Decreased Platelets**

In the post-marketing setting, serious decreased platelets has been reported. In some patients, symptoms such as subcutaneous, mouth, and nasal haemorrhages have been observed. The time to onset is often within 1 month of the start of treatment In some patients, reported decreased platelets may have been related to idiopathic thrombocytopenic purpura. If concomitant symptoms, such as subcutaneous haemorrhage, or remarkable decreased platelets are observed after treatment with FOY-305, the use of FOY-305 should be discontinued, and appropriate treatment (e.g., platelet transfusion) should be provided based on the severity of decreased platelets. No decreased platelet has been observed in subacute or chronic toxicity studies of FOY-305, and its pathogenesis is unknown.

**11.1.3 Hepatic Impairment and Jaundice**

In the post-marketing setting, serious hepatic impairment and jaundice have been reported. While the earliest symptoms included general malaise, nausea/vomiting, and jaundice, hepatic impairment and jaundice were found by laboratory testing with no subjective symptom in many patients. The time to onset after the first dose of FOY-305 was often 1 to 2 months, with some patients experiencing (found to have) hepatic impairment and/or jaundice after completing treatment with FOY-305. If marked abnormality in hepatic function is observed after treatment with FOY-305, the use of FOY-305 should be discontinued, and appropriate treatment (e.g., liver protectants) should be provided, with the patient being placed at rest. No abnormal hepatic function has been observed in subacute or chronic toxicity studies of FOY-305, and its pathogenesis is unknown.

**11.1.4 Hyperkalaemia**

In the post-marketing setting, serious hyperkalaemia has been reported. While hyperkalaemia was found by laboratory testing with no subjective symptom in many patients, some patients had abnormal ECG findings with or without clinical symptoms (general malaise and chest distress). The time to onset after the first dose of FOY-305 was often 2 months or shorter. Patients treated with FOY-305 should be monitored for any change in the potassium level through regular periodic examination. In the event of an abnormal change, the use of FOY-305 should be discontinued, and appropriate treatment (e.g., cation exchange resin, diuretics, and glucose-insulin therapy) should be provided. No abnormal change in the potassium level has been observed in subacute or chronic toxicity studies of FOY-305, and its pathogenesis is unknown (see Section 1.2.3.7, "Important Precautions and Reasons").

##### 1.2.3.10.2 Other Adverse Drug Reactions

**11.2 Other Adverse Drug Reactions**

| | **0.1% to <0.5%** | **<0.1%** | **Frequency unknown** |
|---|---|---|---|
| Blood disorders | | Decreased white blood cells and erythrocytes | Eosinophilia |
| Hypersensitivity Note 2) | Rash, itching, etc. | | |
| Gastrointestinal disorders | Nausea, abdominal discomfort/bloating, and diarrhoea | Anorexia, vomiting, thirst, heartburn, abdominal pain, and constipation | |
| Hepatic disorders | Increased AST/ALT, etc. | | |
| Renal disorders | | Increased BUN and creatinine | |
| Other | | Oedema and hypoglycaemia | |

Note 1) The frequency data include data from the drug use investigation.
Note 2) Treatment with FOY-305 should be discontinued if hypersensitivity occurs.

1.2.3.10.3 List of the Frequencies of Adverse Drug Reactions <Remission of Acute Symptoms in Chronic Pancreatitis>

**Table 1.2-1 Frequencies of Adverse Drug Reactions <Remission of Acute Symptoms in Chronic Pancreatitis> (At the End of Reexamination)**

| | Pre-approval investigations | Post-approval investigations | Total |
|---|---|---|---|
| N<br>Having adverse drug reactions (%)<br>Number of adverse drug reactions | 423 patients<br>12 patients (2.84)<br>13 | 3383 patients<br>57 patients (1.68)<br>70 | 3806 patients<br>69 patients (1.81)<br>83 |
| **Type of adverse drug reactions** | **Number of adverse drug reactions (%)** | | |
| Gastrointestinal disorders | | | |
| Nausea (feeling queasy) | — | 10 (0.30) | 10 (0.26) |
| Vomiting | — | 3 (0.09) | 3 (0.08) |
| Abdominal discomfort | 1 (0.24) | 3 (0.09) | 4 (0.11) |
| Stomach discomfort | — | 3 (0.09) | 3 (0.08) |
| Dyspepsia (heartburn) | 1 (0.24) | — | 1 (0.03) |
| Abdominal distension | — | 6 (0.18) | 6 (0.16) |
| Bowel sounds abnormal | — | 1 (0.03) | 1 (0.03) |
| (borborygmus) | — | 1 (0.03) | 1 (0.03) |
| Abdominal pain | — | 1 (0.03) | 1 (0.03) |
| Abdominal pain upper | — | 1 (0.03) | 1 (0.03) |
| Leukoplakia oral | — | 4 (0.12) | 4 (0.11) |
| Diarrhoea | 1 (0.24) | — | 1 (0.03) |
| Constipation | | | |
| General disorders and administration site conditions | | | |
| Generalised oedema | — | 1 (0.03) | 1 (0.03) |
| Fatigue | — | 2 (0.06) | 2 (0.05) |
| Thirst | 2 (0.47) | 2 (0.06) | 4 (0.11) |
| Hepatobiliary disorders | | | |
| Hepatic function abnormal | — | 2 (0.06) | 2 (0.05) |
| Infections and infestations | | | |
| Nasopharyngitis (common cold syndrome) | — | 1 (0.03) | 1 (0.03) |
| Investigations | | | |
| AST (GOT) increased | — | 1 (0.03) | 1 (0.03) |
| ALT (GPT) increased | — | 1 (0.03) | 1 (0.03) |
| Platelet count decreased | 1 (0.24) | — | 1 (0.03) |
| Metabolism and nutrition disorders | | | |
| Anorexia | 1 (0.24) | 3 (0.09) | 4 (0.11) |
| Nervous system disorders | | | |
| Headache (headache dull) | — | 3 (0.09) | 3 (0.08) |
| Dizziness (wooziness) | 1 (0.24) | — | 1 (0.03) |
| Renal and urinary disorders | | | |
| Pollakiuria | — | 1 (0.03) | 1 (0.03) |
| Skin and subcutaneous tissue | | | |
| disorders | 3 (0.71) | 10 (0.30) | 13 (0.34) |
| Rash | — | 1 (0.03) | 1 (0.03) |
| Drug eruption | — | 1 (0.03) | 1 (0.03) |
| Toxic skin eruption | 2 (0.47) | 7 (0.21) | 9 (0.24) |
| Pruritus | — | 1 (0.03) | 1 (0.03) |
| Hyperhidrosis | | | |

(Data tabulated at the end of reexamination)

MedDRA/J ver. 8.0 was used. Each adverse drug reaction term is presented as the preferred term (PT).

##### 1.2.3.10.4 List of the Frequencies of Adverse Drug Reactions <Postoperative Reflux Esophagitis>

**Table 1.2-2 Frequencies of Adverse Drug Reactions <Postoperative Reflux Esophagitis> (At the End of Reexamination)**

| | Pre-approval investigations | Post-approval investigations | Total |
|---|---|---|---|
| N<br>Having adverse drug reactions (%)<br>Number of adverse drug reactions | 416 patients<br>12 patients (2.88)<br>15 | 3808 patients<br>45 patients (1.18)<br>60 | 4224 patients<br>57 patients (1.35)<br>75 |
| **Type of adverse drug reactions** | **Number of adverse drug reactions (%)** | | |
| Blood and lymphatic system disorders | | | |
| Anaemia | — | 1 (0.03) | 1 (0.02) |
| Leukopenia | — | 1 (0.03) | 1 (0.02) |
| Cardiac disorders | | | |
| Palpitations | — | 1 (0.03) | 1 (0.02) |
| Gastrointestinal disorders | | | |
| Nausea (feeling queasy) | — | 5 (0.13) | 5 (0.12) |
| Vomiting | — | 1 (0.03) | 1 (0.02) |
| Abdominal discomfort | — | 1 (0.03) | 1 (0.02) |
| Epigastric discomfort | — | 1 (0.03) | 1 (0.02) |
| Dyspepsia (heartburn) | — | 1 (0.03) | 1 (0.02) |
| Gastrointestinal upset (stomach feeling heavy) | — | 1 (0.03) | 1 (0.02) |
| Abdominal distension | 1 (0.24) | 1 (0.03) | 2 (0.05) |
| Abdominal pain | — | 1 (0.03) | 1 (0.02) |
| Abdominal pain upper | 1 (0.24) | — | 1 (0.02) |
| Diarrhoea | 1 (0.24) | 7 (0.18) | 8 (0.19) |
| Constipation | — | 1 (0.03) | 1 (0.02) |
| General disorders and administration site conditions | | | |
| Chest pain | 1 (0.24) | — | 1 (0.02) |
| Feeling abnormal (feeling bad) | — | 2 (0.05) | 2 (0.05) |
| Oedema | 2 (0.48) | — | 2 (0.05) |
| Oedema peripheral (leg oedema) | 1 (0.24) | 1 (0.03) | 2 (0.05) |
| Feeling hot | 1 (0.24) | — | 1 (0.02) |
| Thirst | — | 1 (0.03) | 1 (0.02) |
| Hepatobiliary disorders | | | |
| Hepatic function abnormal | — | 5 (0.13) | 5 (0.12) |
| Liver disorder | — | 1 (0.03) | 1 (0.02) |

| | Pre-approval investigations | Post-approval investigations | Total |
|---|---|---|---|
| N | 416 patients | 3808 patients | 4224 patients |
| Having adverse drug reactions (%) | 12 patients (2.88) | 45 patients (1.18) | 57 patients (1.35) |
| Number of adverse drug reactions | 15 | 60 | 75 |
| **Type of adverse drug reactions** | **Number of adverse drug reactions (%)** | | |
| Laboratory test | | | |
| AST (GOT) increased | — | 4 (0.11) | 4 (0.09) |
| ALT (GPT) increased | — | 2 (0.05) | 2 (0.05) |
| γ-GTP increased | — | 1 (0.03) | 1 (0.02) |
| Blood alkaline phosphatase | — | 1 (0.03) | 1 (0.02) |
| increased | — | 1 (0.03) | 1 (0.02) |
| Blood potassium increased | — | 2 (0.05) | 2 (0.05) |
| Blood urea increased | — | 2 (0.05) | 2 (0.05) |
| Red blood cell count decreased | — | 2 (0.05) | 2 (0.05) |
| Protein total decreased | — | 1 (0.03) | 1 (0.02) |
| Zinc sulphate turbidity abnormal | | | |

**Table 1.2-2 Frequencies of Adverse Drug Reactions <Postoperative Reflux Esophagitis> (At the End of Reexamination) (continued)**

| | **Pre-approval investigations** | **Post-approval investigations** | **Total** |
|---|---|---|---|
| N<br>Having adverse drug reactions (%)<br>Number of adverse drug reactions | 416 patients<br>12 patients (2.88)<br>15 | 3808 patients<br>45 patients (1.18)<br>60 | 4224 patients<br>57 patients (1.35)<br>75 |
| **Type of adverse drug reactions** | **Number of adverse drug reactions (%)** | | |
| Metabolism and nutrition disorders | | | |
| Dehydration | — | 1 (0.03) | 1 (0.02) |
| Hypoglycaemia | 1 (0.24) | — | 1 (0.02) |
| Nervous system disorders | | | |
| Headache | — | 1 (0.03) | 1 (0.02) |
| Dizziness (wooziness) | — | 3 (0.08) | 3 (0.07) |
| Depressed level of consciousness | — | 1 (0.03) | 1 (0.02) |
| Hypoaesthesia (numbness) | — | 1 (0.03) | 1 (0.02) |
| Psychiatric disorders | | | |
| Irritability (frustration) | — | 1 (0.03) | 1 (0.02) |
| Renal and urinary disorders | | | |
| Renal impairment | 1 (0.24) | — | 1 (0.02) |
| Respiratory, thoracic and mediastinal disorders | | | |
| Nasal congestion | 1 (0.24) | — | 1 (0.02) |
| Skin and subcutaneous tissue disorders | 1 (0.24) | — | 1 (0.02) |
| Rash | 2 (0.48) | — | 2 (0.05) |
| Urticaria | — | 2 (0.05) | 2 (0.05) |
| Eczema | — | 1 (0.03) | 1 (0.02) |
| Drug eruption | 1 (0.24) | — | 1 (0.02) |
| Pruritus | | | |

(Data tabulated at the end of reexamination)
MedDRA/J ver. 8.0 was used. Each adverse drug reaction term is presented as the PT.

##### 1.2.3.10.5 Frequencies of Adverse Drug Reactions by Patient Characteristic (Extracted) <Remission of Acute Symptoms in Chronic Pancreatitis>

The incidences of adverse drug reactions by patient characteristic in 3383 patients in the post-marketing drug use investigation are shown below.

**Table 1.2-3 Frequencies of Adverse Drug Reactions by Patient Characteristic (Extracted) <Remission of Acute Symptoms in Chronic Pancreatitis> (Drug Use Investigation)**

| | | Number of patients | Number of patients with adverse drug reactions | Incidence of adverse drug reactions | $\chi^2$ test |
|---|---|---|---|---|---|
| Total | | 3383 | 57 | 1.7% | — |
| Sex | Male | 1881 | 26 | 1.4% | N.S. |
| | Female | 1497 | 31 | 2.1% | |
| | Unknown | 5 | 0 | 0.0% | — |
| Severity at baseline | Mild | 1772 | 32 | 1.8% | N.S. |
| | Moderate | 1336 | 21 | 1.6% | |
| | Severe | 147 | 2 | 1.4% | |
| | Unknown | 128 | 2 | 1.6% | — |
| Concurrent illness | No | 1431 | 31 | 2.2% | N.S. |
| | Yes | 1939 | 26 | 1.3% | |
| | Unknown | 13 | 0 | 0.0% | — |

| | | Number of patients (cumulative) | Number of patients with adverse drug reactions | Incidence of adverse drug reactions |
|---|---|---|---|---|
| Duration of treatment at the onset | <2 weeks | 3383 | 33 | 1.0% |
| | 2 to <4 weeks | 3285 | 8 | 0.2% |
| | 4 to <8 weeks | 2896 | 12 | 0.4% |
| | 8 to <12 weeks | 1892 | 2 | 0.1% |
| | ≥12 weeks | 1361 | 2 | 0.1% |
| | Unknown | 12 | 0 | 0.0% |
| Total number of tablets at the onset | <84 | 3383 | 37 | 1.1% |
| | 84 to <168 | 3226 | 5 | 0.2% |
| | 168 to <336 | 2780 | 12 | 0.4% |
| | 336 to <504 | 1755 | 1 | 0.1% |
| | ≥504 | 1208 | 1 | 0.1% |
| | Unknown | 26 | 1 | 3.8% |

N.S.: No significant difference

The incidences of adverse drug reactions were higher in patients with a treatment duration of <2 weeks and those with a total number of tablets of <84; however, the results reflect the large number of patients with adverse drug reactions observed early after treatment initiation.

1.2.3.10.6 Frequencies of Adverse Drug Reactions by Patient Characteristic (Extracted) <Postoperative Reflux Esophagitis>

The incidences of adverse drug reactions by patient characteristic in 3228 and 580 patients in the post-marketing drug use investigation and the special investigation on the long-term use, respectively, are shown below.[12)]

**Table 1.2-4 Frequencies of Adverse Drug Reactions by Patient Characteristic (Extracted) <Postoperative Reflux Esophagitis> (Drug Use Investigation)**

| | | | Drug use investigation | | | Special investigation on the long-term use | | |
|---|---|---|---|---|---|---|---|---|
| | | | Number of patients | Number of patients with adverse drug reactions | $\chi^2$ test | Number of patients | Number of patients with adverse drug reactions | $\chi^2$ test |
| Total | | | 3228 | 31 (1.0%) | — | 580 | 14 (2.4%) | — |
| Sex | Male | | 2050 | 20 (1.0%) | N.S. | 388 | 9 (2.3%) | N.S. |
| | Female | | 1129 | 11 (1.0%) | | 192 | 5 (2.6%) | |
| | Unknown/not provided | | 49 | 0 (0.0%) | — | 0 | — | — |
| Age | <15 years old | | 0 | — | N.S. | 0 | — | N.S. |
| | 15 to 64 years old | | 1493 | 18 (1.2%) | | 266 | 4 (1.5%) | |
| | ≥65 years old | | 1731 | 13 (0.8%) | | 314 | 10 (3.2%) | |
| | Unknown/not provided | | 4 | 0 (0.0%) | — | 0 | — | — |
| Predisposition to hypersensitivity | No | | 3194 | 30 (0.9%) | N.S. | 565 | 8 (1.4%) | *** |
| | Yes | | 31 | 1 (3.2%) | | 15 | 6 (40.0%) | |
| | Unknown/not provided | | 3 | 0 (0.0%) | — | 0 | — | — |
| Concurrent illness | No | | 2374 | 22 (0.9%) | N.S. | 423 | 7 (1.7%) | N.S. |
| | Yes | | 849 | 9 (1.1%) | | 141 | 7 (5.0%) | |
| | Unknown/not provided | | 5 | 0 (0.0%) | — | 16 | 0 (0.0%) | — |
| | Breakdown | Hepatic disorders | 177 | 2 (1.1%) | — | 17 | 0 (0.0%) | — |
| | | Renal disorders | 39 | 0 (0.0%) | | 5 | 0 (0.0%) | |
| | | Upper gastrointestinal tract disorders | 24 | 0 (0.0%) | | 5 | 0 (0.0%) | |
| | | Cancers/tumors | 55 | 0 (0.0%) | | 29 | 1 (3.4%) | |
| | | Other | 616 | 7 (1.1%) | | 117 | 6 (5.1%) | |
| Severity at baseline | Mild | | 1632 | 19 (1.2%) | N.S. | 289 | 7 (2.4%) | N.S. |
| | Moderate | | 1425 | 11 (0.8%) | | 264 | 5 (1.9%) | |
| | Severe | | 134 | 0 (0.0%) | | 22 | 2 (9.1%) | |
| | Unknown/not provided | | 37 | 1 (2.7%) | — | 5 | 0 (0.0%) | — |
| Type of gastrectomy | Total | | 1165 | 14 (1.2%) | N.S. | 211 | 4 (1.9%) | N.S. |
| | Cardiac | | 179 | 2 (1.1%) | | 38 | 0 (0.0%) | |
| | Pyloric | | 1662 | 14 (0.8%) | | 294 | 10 (3.4%) | |
| | Other | | 189 | 1 (0.5%) | | 35 | 0 (0.0%) | |
| Concomitant medication | No | | 386 | 4 (1.0%) | N.S. | 39 | 3 (7.7%) | N.S. |
| | Present | | 2841 | 27 (1.0%) | | 537 | 11 (2.0%) | |
| | Unknown/not provided | | 1 | 0 (0.0%) | | 4 | 0 (0.0%) | — |
| | Breakdown | Antiulcer | 1681 | 13 (0.8%) | — | 318 | 5 (1.6%) | — |
| | | drugs/antacids | 2186 | 24 (1.1%) | | 430 | 8 (1.9%) | |
| | | Other gastrointestinal | | | | | | |
| | | drugs | 655 | 7 (1.1%) | | 159 | 3 (1.9%) | |
| | | Anti-tumor drugs | 96 | 1 (1.0%) | | 23 | 1 (4.3%) | |
| | | Other | | | | | | |

N.S.: No significant difference; ***: p<0.001

Based on the higher incidence of adverse drug reactions in patients with predisposition to hypersensitivity in the special investigation, “Patients with Hypersensitivity” was included in Section 1.2.3.8, “Precautions for Patients with Certain Characteristics.”

##### 1.2.3.11 Effects on Laboratory Test Results

None

##### 1.2.3.12 Overdose

None

##### 1.2.3.13 Precautions for Application

**14. Precautions for Application**
**14.1 Precautions for Drug Preparation**
Single-dose packaging of FOY-305 and packaging together with an olmesartan medoxomil product should be avoided. The storage of single-dose packages under hot and humid conditions may result in FOY-305 discoloration.
**14.2 Precautions for Drug Prescription**
For blister packaged medications, patients should be instructed to take each dose out of the blister pack. If accidentally ingested, the hard edge of a blister pack may be inserted into the esophageal mucosa, which can cause perforation that may result in serious complications, such as mediastinitis.

**(Rationale)**

**14.1** Additional Relevant Document “Change Caused by Combining with an Olmesartan Medoxomil Product”

[Study methods]

One tablet of FOY-305 (100 mg) was packaged with 1 tablet of an olmesartan medoxomil product (20 mg) using polyethylene laminated cellophane paper, and changes in appearance were visually compared with FOY-305 at baseline under various conditions (away from light).

[Results]

Under 25°C and 60%RH conditions, no change in appearance was observed for 3 months. Under 25°C and 75%RH and 30°C and 75%RH conditions, the packaged FOY-305 turned to very pale red at 3 and 2 months, respectively, showing that the packaged FOY-305 discolored earlier under hotter and more humid conditions.

| Time / Conditions | Baseline | Week 1 | Week 2 | Week 3 | Month 1 | Month 2 | Month 3 |
|---|---|---|---|---|---|---|---|
| 25°C, 60%RH | White | (-) | (-) | (-) | (-) | (-) | (-) |
| 25°C, 75%RH | White | (-) | (-) | (-) | (-) | (-) | (±) |
| 30°C, 75%RH | White | (-) | (-) | (-) | (-) | (±) | (±) |

Criteria for the determination of appearance:
(-), no change; (±), turned to very pale red (difficult to discriminate from the control); (+), turned to pale red; (++), turned to slightly red; and (+++), turned to red.

**14.2** The precaution was selected as important and general for blister packaged medications, according to the Measures against Accidental Ingestion of Blister Packs (FPMAJ No. 240 issued on 27 March 1996 and FPMAJ No. 304 issued on 18 April 1996).

##### 1.2.4 Other Precautions

###### 1.2.4.1 Information based on Clinical Experience

For the indication of remission of acute symptoms in chronic pancreatitis, camostat mesilate should in general be orally administered at a daily dose of 600 mg in three divided doses. The dose should be increased or reduced based on symptoms. For clinical safety information on doses exceeding the approved dose, a single dose of FOY-305 50 to 600 mg administered to 6 healthy adult males aged 24 to 39 years after a meal in a Phase I study caused no abnormal finding in any of the subjects.[13)] In a double-blind controlled study in patients with postoperative reflux esophagitis, in which repeated doses of FOY-305 300 mg were administered 3 times a day after a meal, for 8 weeks as a general rule, 3 adverse drug reactions (oedema, feeling of nasal congestion, and urticaria) were observed in 2/62 patients ; however, none of these adverse drug reactions were reported to be clinically significant.[14)] A clinical research report on 6-month treatment with FOY-305 7.2 g/day in several patients with cancer reported no tolerability issue.[15)] In a study in which healthy adults received FOY-305 600 mg 4 times a day, 2 adverse events (hyperuricaemia and aphthous ulcer) were observed in 2 subjects, both of which were considered as adverse drug reactions. Both of these events were Grade 1 in severity, and there was no clinically significant adverse event. In a study in which healthy adults received repeated doses of FOY-305 600 mg 4 times a day for 7 days, 2 adverse events (hyperuricaemia and aphthous ulcer) were observed in 2 subjects, both of which were considered as adverse drug reactions. Both of these events were Grade 1 in severity, and there was no clinically significant adverse event.

Based on the above, it is unlikely that the safety risk of FOY-305 is significantly increased at doses exceeding the approved dose. As with administration at the approved dose, however, subjects should be carefully monitored for clinically significant adverse drug reactions (including shock, anaphylaxis, platelets decreased, hepatic impairment, jaundice, and hyperkalaemia) and commonly reported adverse drug reactions (including hypersensitivity [rash, itching, etc.], gastrointestinal disorders [nausea, abdominal discomfort/distension, and diarrhoea], and increased AST/ALT) during the present study.

###### 1.2.4.2 Information based on Non-Clinical Studies

Safety evaluation showed effects on general condition (including respiratory distress, decreased spontaneous locomotor activity, overreaction to external stimuli, and convulsions), heart rate, blood pressure, breathing, and the urinary system. Safety evaluation also showed malnutrition related to effects of overactivity of the pharmacological action of FOY-305 (protease enzyme inhibition) on digestion and absorption of foods, and gastrointestinal effects (including vomiting and gastrointestinal damage). Given that FOY-305 was well-tolerated at doses exceeding the approved dose in clinical studies and the clinical research report, as shown in Section 1.2.4.1, "Information based on Clinical Experience," it is unlikely that FOY-305 will have clinically significant adverse effects in the present clinical study; however, subjects should be monitored for these effects.

Based on suppressed fetal weight gain observed after administration of FOY-305 to perinatal and breastfeeding rats, it is recommended that the use of FOY-305 in women who are pregnant or possibly pregnant should be limited to cases in which treatment benefits are considered to outweigh the risks.

### 2 STUDY ORGANIZATION AND ROLES

Refer to Supplementary Volume 1, "Study Organization."

Supplementary Volume 1 shall be revised separately from the protocol.

### 3 STUDY OBJECTIVES

To assess the efficacy and safety of FOY-305 in patients with COVID-19 infection in a placebo-controlled, multicenter, double-blind, randomized, parallel-group comparative study.

### 4 SUBJECTS

#### 4.1 Subjects

COVID-19

#### 4.2 Inclusion Criteria

Patients will be enrolled only if they have agreed to participate in this study and meet all the following criteria at the time of enrollment:

1. Male or female
2. Aged ≥18 years (at the time of providing consent)
3. Inpatient or outpatient: Inpatient

4. Positive SARS-CoV-2 test by a method eligible for definitive diagnosis (e.g., RT-PCR test, LAMP test, antigen test)
5. Enrollment in this study within 5 days after onset of symptoms of SARS-CoV-2 infection (COVID-19) (for asymptomatic patients, enrollment in this study within 5 days after collection of the sample that tested positive)
6. Women of childbearing potential (including patients with amenorrhea due to chemically induced menopause or for other medical reasons)[#1] should agree to use contraceptive method(s)[#2] from the time of consent to 1 month after last administration of the study drug
7. Male subjects should agree to use contraceptive method(s)[#2] from the time of initiation of study drug administration to 1 month after last administration of the study drug

[#1]: Women of childbearing potential include all women who have experienced menarche, have not undergone sterilization (hysterectomy, bilateral tubal ligation, or bilateral oophorectomy), and have not experienced menopause. Menopause is defined as the absence of menses for ≥12 months without any particular reason. Women using oral contraceptives, intrauterine devices, or diaphragms are considered to be of childbearing potential.

[#2]: Dual contraceptive method using 2 of the following methods should be employed: vasectomy or condom for male patient or male partner, and tubal ligation, diaphragm, intrauterine device, or oral contraceptive for female patient or partner.

<Rationale>

1. On the basis of the judgment that restriction of study subjects by gender is not necessary.
2. To define the age at which the patient by himself/herself is able to provide consent to participate in the study.
3. To evaluate the drug efficacy appropriately.
4. To enroll patients with SARS-CoV-2 infection.
5. To enroll patients considered appropriate for evaluation of drug efficacy.

6. to 7. In consideration of the safety of pregnant women and fetuses.

##### 4.3 Exclusion Criteria

Patients who meet any of the following criteria at enrollment shall be excluded from the study:

1. Receiving oxygen therapy
2. Difficulty in swallowing oral medication
3. History of SARS-CoV-2 infection (COVID-19)

4. Serum potassium >5.5 mEq/L
5. Total bilirubin ≥3.0 mg/dL, or AST (GOT) or ALT (GPT) ≥2.5 × upper limit of normal of the institution (or ≥100 IU/L)
6. Serum creatinine ≥2.0 mg/dL
7. History of vaccination (including unapproved ones) against SARS-CoV-2 infection (COVID-19)
8. History of treatment with camostat mesilate or nafamostat mesilate for SARS-CoV-2 infection (COVID-19)
9. Taking camostat mesilate
10. History of hypersensitivity to camostat mesilate
11. Patients who are pregnant, breast-feeding, or possibly pregnant, or who wish to be pregnant during the study
12. History of treatment specified as prohibited therapy or concomitant therapy before initiation of the study treatment
13. History of serious drug allergy
14. Judged as lacking the ability to consent, for example, because of comorbid dementia
15. Judged by the investigator or subinvestigator as inappropriate as a study subject for any other reason

- The eligibility of patients should be assessed on the basis of the information from the observation period.
- For items 4, 5, and 6, if multiple actual values are available from the observation period, use the value obtained on the day closest to the day of enrollment for judgment.

<Rationale>

1. To exclude patients who are not appropriate for evaluation of drug efficacy.
2. To exclude patients who are not appropriate for evaluation of drug efficacy.
3. To exclude patients who are not appropriate for evaluation of drug efficacy.
4. In consideration of the safety of patients, because serious hyperkalemia has been reported with camostat mesilate in the post-marketing period.

5. to 6. In consideration of the safety of patients.

7. To exclude patients who are not appropriate for evaluation of drug efficacy.
8. To exclude patients who are not appropriate for evaluation of drug efficacy.

9. In consideration of the safety of patients, and to exclude patients who are not appropriate for evaluation of drug efficacy.
10. In consideration of the safety of patients.
11. In consideration of the safety of pregnant women and fetuses.
12. In consideration of the safety of patients, and to exclude patients who are not appropriate for evaluation of drug efficacy.
13. In consideration of the safety of patients.
14. To protect the human rights of patients.
15. In consideration of possible conditions that are not appropriate for the study other than 1 to 14 listed above.

### 5 STUDY METHODS

#### 5.1 Study Design

##### 5.1.1 Study Design

This study will be conducted as a multicenter, double-blind, randomized, parallel-group comparative study.

This study consists of the following phases: an observation phase of up to 3 days with no study treatment administered; a double-blind phase of up to 14 days in which subjects will receive double-blinded study treatment; and a follow-up phase of 2 weeks with no study treatment administered. Subjects will be randomized in a 1:1 ratio to receive FOY-305 or placebo in a double-blind fashion through a dynamic allocation procedure, with site, age (≥65 vs <65 years), and underlying disease (chronic respiratory disease, chronic kidney disease, diabetes mellitus, hypertension, cardiovascular diseases, obesity [BMI ≥30 $kg/m^2$]) (present vs absent) as allocation factors.

It has been planned that an interim analysis will be performed to determine whether the treatment is effective or ineffective in the subjects randomized as of the time when further enrollment appears to be difficult because of convergence of the COVID-19 pandemic during the subject enrollment period.

<Rationale>

The observation phase of this study has been included to confirm the eligibility as study subjects. The placebo-controlled, double-blind, comparative design has been adopted to evaluate the efficacy and safety of FOY-305 objectively by eliminating biases from evaluators and subjects. The double-blind phase is up to 14 days in duration because progression from onset of the symptoms, such as pyrexia and malaise, to development of severe pneumonia requiring oxygen therapy and mechanical ventilation is thought to be 1 to 2 weeks. A 2-week period following the final administration is designated as a follow-up phase to ensure the safety of subjects after a certain period after administration of FOY-305. A multicenter design has been adopted to allow for accumulation of the target number of subjects for the study, efficient evaluation of the efficacy and safety of FOY-305, and generalization of the results to be obtained in the study.

The interim analysis of this study has been included in anticipation of the possible end of the pandemic during the study period as the future status of the COVID-19 pandemic is difficult to predict.

###### 5.1.2 Planned Number of Subjects

110 subjects in total, with 55 per group

###### 5.1.3 Planned Study Period

August 2020 to October 2021

Enrollment may be ended early depending on the enrollment status of subjects.

###### 5.1.4 Definition of the End of Study for Individual Subjects

The end of study for individual subjects is defined as the time point at which all protocol-specified assessments are completed. For subjects discontinuing treatment, “treatment discontinuation” will be replaced by “end of study.” The date of study completion will be entered into the electronic case report form (eCRF).

In studies with follow-up, however, the follow-up phase will be handled as a post-study phase.

##### 5.2 Assignment of Subjects to Treatment Groups

###### 5.2.1 Study Treatment Allocation Procedure

The person responsible for study drug allocation will randomly allocate subjects into the FOY-305 or placebo group in a 1:1 ratio.

The study drug will be allocated to subjects at the time of enrollment. If a subject is eligible, the FOY-305-01 Enrollment Center will assign the subject to the FOY-305 or placebo group through the dynamic allocation procedure and issue the assignment result.

The dynamic allocation will be performed by the minimization method with the following allocation factors:

- Medical institution
- Age (≥65 vs <65 years)
- Presence or absence of underlying disease (chronic respiratory disease, chronic kidney disease, diabetes mellitus, hypertension, cardiovascular diseases, obesity [BMI ≥30 kg/$m^2$])

##### 5.2.2 Enrollment of Subjects

Patients will be enrolled as subjects in accordance with the procedures below.

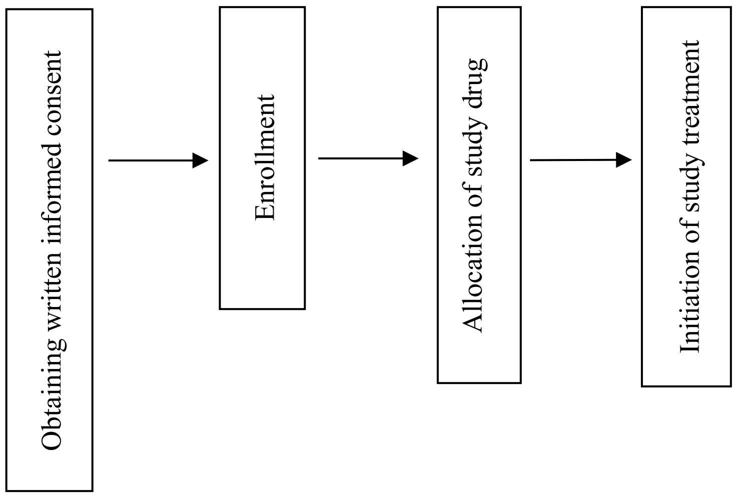

###### 5.2.2.1 Enrollment

After confirming the eligibility of the patient, the investigator or subinvestigator will enroll the patient at the FOY-305-01 Enrollment Center in accordance with the separately specified "Procedures for Patient Enrollment." If the patient is eligible, the Enrollment Center will dynamically assign the subject to the FOY-305 or placebo group (see Section 5.2.1, "Study Treatment Allocation Procedure"), and send the study treatment number to the investigator or subinvestigator via web or email. After confirming that the patient has been enrolled as a subject, the investigator or subinvestigator will initiate the study treatment. The study treatment will be initiated no later than the day after enrollment. If a subject assessed as eligible discontinues the study prior to the first dose of the study drug for any reason, the pre-dose discontinuation will be reported to the Enrollment Center. If the dates of enrollment (assignment) and treatment initiation are not the same, the date of enrollment will be used as Day 1.

| |
|---|
| <u>FOY-305-01 Enrollment Center</u><br>URL: https://e-medinfo.com/webrc/ono/foy-305-01/<br>TEL: 0120-203-541<br>Available via web: 24 hours per day<br>Helpdesk available: From 09:00 to 20:00 on Monday through Saturday<br>(Except for 13 to 15 August and 29 December to 5 January [public/national holidays]) |

#### 5.3 Blinding

##### 5.3.1 Preparation of Randomization Table

The person responsible for study drug allocation will prepare the randomization table. After allocation of the study drug, the person responsible for study drug allocation will seal the original copy of the randomization table. At the request of the sponsor, the person responsible for study drug allocation will prepare and seal a copy of the original randomization table. The storage of the randomization table will be specified in a separately prepared procedural document. After allocation of the study drug, the person responsible for study drug allocation will prepare/seal 1 copy of the breakable emergency code per subject for use in the case of an emergency such as the occurrence of a serious adverse event.

##### 5.3.2 Maintenance of Blindness

The study will be conducted in a double-blind manner. The sponsor will supply study drugs that are identical in appearance, and maintain the blindness using IWRS. At the time of study treatment allocation and at the end of study, the person responsible for study drug allocation will confirm that the package configuration, labeling, and other requirements are identical in appearance. The Emergency Contact Center will keep the emergency code securely up to the time of code breaking. The drug accountability manager should first seal any remaining study drugs before the sponsor collects them before code breaking.

##### 5.3.3 Code-Breaking Procedure

###### 5.3.3.1 Emergency Code-Breaking Procedure

1. If it becomes necessary to urgently know the allocation of the study drug due to a reason such as the occurrence of a serious adverse event, the investigator or subinvestigator shall contact the Emergency Contact Center (See page 2) to request code breaking of the emergency code for the subject.
2. When requested to break the emergency code, the Emergency Contact Center will break the emergency code of the applicable subject immediately, inform the investigator or subinvestigator

of the allocation of the study drug, and inform the sponsor of the code breaking. The sponsor, when receiving the information from the Emergency Contact Center, will report the result to the medical expert and the coordinating investigator.

3. After breaking the emergency code, the investigator will prepare an eCRF for the subject that contains data collected before breaking of the emergency code, provide an electronic signature, and submit it to the sponsor as immediately as possible. The date of code breaking will be entered into the eCRF.
4. If the regulatory authorities request reporting of the allocation of the study drug for the subject due to a reason such as the occurrence of a serious adverse event, the sponsor shall request the Emergency Contact Center to break the emergency code for the subject in accordance with a separately defined procedure. In this case, the result will not be disclosed to the investigator or subinvestigator.

###### 5.3.3.2 Code-Breaking Procedure for Randomization Table

The procedures below will be followed for code breaking after the study is completed. The procedures to be performed at the interim analysis will be specified in a separately prepared procedural document.

1. If there are any unexpected issues related to handling the subjects before code breaking, the sponsor shall discuss those details with the medical expert and decide the handling in the analysis.
2. The person responsible for study drug allocation shall reconfirm the handling in the analysis of all enrolled subjects in accordance with the criteria defined in the protocol and the discussion result in 1. by the time of code breaking.
3. The person responsible for study drug allocation shall confirm the unidentifiability of the study drug during the study period, and confirm the presence/absence of breaking of emergency code and that the original and copy of the randomization table are sealed. If the emergency code has been broken, it shall be reconfirmed that the code breaking for the subjects had been conducted appropriately.
4. The person responsible for study drug allocation shall break the randomization table.

#### 5.4 Endpoints

##### 5.4.1 Efficacy Endpoints

###### 5.4.1.1 Primary Endpoint

Time to negative test for SARS-CoV-2 as assessed by the local laboratory

SARS-CoV-2 negative test is defined as that confirmed by SARS-CoV-2 test performed on 2 consecutive occasions (after an interval of at least 24 hours), and the date of sample collection for the test that first shows a negative test result will be used as the date of confirmed SARS-CoV-2 negative test.

The time to negative test for SARS-CoV-2 is defined as in the following formula:

“Date of the first confirmation of SARS-CoV-2 negative test” − “Date of randomization” + 1

<Rationale>

The time to negative test for SARS-CoV-2 has been selected as the primary endpoint because achievement of negativity in an early phase can lead to cure and the primary goal of treatment for viral infection is viral clearance.

###### 5.4.1.2 Secondary Endpoints

1. Time to negative test for SARS-CoV-2 as assessed by the central laboratory
2. Proportion of subjects who test negative for SARS-CoV-2 (as assessed by the local and central laboratories)
3. Ordinal scale for severity
4. Proportion of subjects on mechanical ventilator
5. Survival status

<Rationale>

The endpoints have been selected for multilateral evaluation of the efficacy of FOY-305 for SARS-CoV-2 infection (COVID-19).

###### 5.4.1.3 Exploratory Endpoints

1. SARS-CoV-2 viral load (as measured by the central laboratory)
2. Presence or absence of lung lesion as observed by chest imaging
3. Time to resolution of clinical symptoms and proportion of subjects in whom clinical symptoms have resolved
4. Antibody response (IgM and IgG) (measured by the central laboratory)

<Rationale>

The endpoints have been selected for multilateral evaluation of the efficacy of FOY-305 for SARS-CoV-2 infection (COVID-19).

##### 5.4.2 Safety Endpoints

1. Adverse events
2. Laboratory tests (hematology, blood biochemistry, qualitative urinalysis)
3. Vital signs (body temperature, $SpO_2$, systolic/diastolic blood pressure, pulse rate)
4. 12-lead ECG

<Rationale>

The endpoints have been selected for multilateral evaluation of the safety of FOY-305 for SARS-CoV-2 infection (COVID-19).

### 6 STUDY DRUGS

All study drugs used in this study shall be manufactured, handled, stored, and managed in compliance with the "Good Manufacturing Practice for Investigational Products (IP-GMP)."

#### 6.1 Names of the Study Drug and Other Drugs to Be Used in the Study

##### 6.1.1 Study Drug

1. Study drug: FOY-305 100 mg Tablets (non-proprietary name: camostat mesilate)
2. Control drug: FOY-305-matched placebo

#### 6.2 Dosage Form and Content of Study Drug

The following formulations will be used in this study:

| Formulation | Dosage form | Content |
|---|---|---|
| FOY-305 100 mg tablets | Film-coated tablet | Each tablet contains FOY-305 100 mg. |
| FOY-305-matched placebo | Placebo tablets contain no FOY-305 but are identical in appearance with FOY-305 100 mg tablets. | |

#### 6.3 Packaging and Labeling of Study Drug

##### 6.3.1 Packaging

FOY-305 100 mg tablets or placebo: 50 tablets/bottle/box

For more information, see "Study Drug Accountability Procedure."

##### 6.4 Supply of Study Drug

After conclusion of the study contract, the study drug will be supplied to each study site.

##### 6.5 Procedures for Study Drug Management and Return of Unused Study Drug

###### 6.5.1 Storage and Management of Study Drug

The drug accountability manager at the study site shall appropriately store and manage the study drug, and prepare an study drug accountability log to keep track of the usage of study drug in accordance with the “Study drug Accountability Procedure.” The drug accountability manager and the sponsor shall check the study drug management record, the number of remaining vials, and entries in the eCRFs for consistency, and immediately investigate causes for discrepancies, if any, and make necessary corrections.

###### 6.5.2 Return of Unused Study Drug

The drug accountability manager will return unused study drugs to the sponsor in accordance with the “Study Drug Accountability Procedure.” At the time of return, individual identifying information such as patient names (e.g., initials) shall be made illegible. If these are returned to the sponsor before code breaking, the drug accountability manager shall count, record, seal, and return the unused drugs. If unused study drugs are lost, the content and the reason for the loss shall be recorded.

#### 7 TREATMENT PLAN

##### 7.1 Study Drug

###### 7.1.1 Dosage and Administration and Duration of Treatment

FOY-305 600 mg (six 100-mg tablets) or placebo (6 placebo tablets) will be orally administered 4 times daily, before breakfast, before lunch, before evening meal, and at bedtime. Patients should be fasted for at least 1 hour after administration. The treatment period is up to 14 days. Patients will discontinue the study treatment when tested negative for SARS-CoV-2 on 2 consecutive occasions (at an interval of ≥24 hours). Once the eligibility is confirmed, the study treatment may be initiated from the earliest dosing time point mentioned above. Although the treatment period is up to 14 days, patients taking the first dose before lunch or before evening meal on the first day will receive the study treatment over 15 days at maximum (up to 56 doses) following the specified dosing regimen thereafter.

<Rationale>

The plasma concentration (mean±standard deviation) of FOY-251, an active metabolite, was 88.9±36.7 ng/mL and 15.7±18.3 ng/mL at 3 and 5 hours, respectively, after administration of FOY-305 as a single dose of 600 mg to healthy adult males. On the basis of these data, the plasma FOY-251 concentration at 4 hours post-dose is expected to be around 50 ng/mL. However, Hoffmann et al. reported that FOY-305 inhibited the effects of TMPRSS2[5] and that FOY-305 inhibited penetration of SARS-CoV-2-S protein (SARS-2-S) into human airway epithelial cell-derived cells in a dose-dependent manner, with a half-maximal effective concentration (EC50) of 87 nmol/L (27.3 ng/mL).[16] Given that FOY-251 has an inhibitory activity on trypsin, plasma kallikrein, plasmin, and others that is almost similar to that of FOY-305, the EC50 of FOY-251 for inhibiting penetration of SARS-2-S into human airway epithelial cell-derived cells is expected to be similar to that of FOY-305. These findings suggest that, when FOY-305 600 mg is administered 4 times daily, with each dose separated by 4 hours, the plasma FOY-251 concentration is expected to remain above the EC50 for about 16 hours or longer. In a Japanese Phase I clinical study in healthy adult males (protocol number: FOY-305-02), FOY-305 was shown to be tolerated when administered at 600 mg/dose, 2400 mg per day; thus, the dosing regimen has been determined to be FOY-305 600 mg (six 100-mg tablets) orally taken 4 times daily. An evaluation of the effects of food on the pharmacokinetics of FOY-305 showed no marked difference in exposure between administration of FOY-305 1 hour before starting the meal and administration in the fasting state, while the exposure after administration of FOY-305 in the fed condition or 30 minutes before starting a meal was lower than that after administration in the fasted state. Thus, administration in the fasted state or at least 1 hour before starting a meal seemed necessary. The duration of treatment is up to 14 days because COVID-19 is clinically characterized by a relatively long incubation period ranging from 1 to 14 days (5 days is the most frequent) and flu-like symptoms such as pyrexia, respiratory symptoms including cough, and general malaise, lasting around 1 week and also because the SARS-CoV-2 viral load reportedly resolves spontaneously at about 20 days after the onset of symptoms.[17), 18]

#### 7.2 Treatments and Concomitant Therapies

##### 7.2.1 Prohibited Treatments and Permitted Therapies

###### 7.2.1.1 Prohibited Treatments and Concomitant Therapies

1. Drugs for the treatment of SARS-CoV-2 infection (COVID-19)

   Use of the following drugs is prohibited from the day of onset of symptoms of SARS-CoV-2 infection (COVID-19) (for asymptomatic patients, the day of collection of the sample that tested positive) until completion of the study treatment: However, continued use of the drug for a comorbidity is permitted (in principle, to be continued with the same dosing regimen during the period from the informed consent to completion of the study treatment).

Drugs with antiviral effects (remdesivir, favipiravir, ciclesonide, nafamostat mesilate, hydroxychloroquine, ivermectin, combination drug of lopinavir and ritonavir, povidone-iodine [e.g., used in the oral and nasal cavities]) and drugs with anticytokine effects (e.g., tocilizumab, JAK inhibitors)

2. Any other unapproved drugs not intended for the treatment of SARS-CoV-2 infection (COVID-19) (including those administered in clinical studies, unapproved combination drugs, and drugs with new dosage form)

   Use of any other unapproved drugs is prohibited from 28 days before initiation of the study treatment to completion of the study treatment.

3. Camostat mesilate in the market

   Use of camostat mesilate in the market is prohibited from initiation of the study treatment to completion of the study treatment.

<Rationale>

1. To eliminate the effect on drug efficacy evaluation.
2. To eliminate the effect of unapproved drugs with uncertain efficacy.
3. To eliminate the effect on drug efficacy evaluation.

###### 7.2.1.2 Permitted Therapies

Of the drugs for the treatment of SARS-CoV-2 infection (COVID-19), those that do not fall under the category of Section 7.2.1.1, “Prohibited Treatments and Concomitant Therapies” will be permitted (e.g., antipyretics, antibiotics, corticosteroids, oxygen therapy).

### 8 STUDY SCHEDULE AND OBSERVATION ITEMS

#### 8.1 Study Schedule

See Section 8, “STUDY SCHEDULE AND OBSERVATION ITEMS” in the Protocol Synopsis.

##### 8.1.1 Observation Phase (Days −3 to 1)

The investigator or subinvestigator will perform the protocol-specified tests to confirm the eligibility of subjects. Subjects meeting any of the criteria specified in Section 13.1, “Rules or Criteria for Treatment Discontinuation of Individual Patients” will not move to the treatment period.

##### 8.1.2 Treatment Phase [Days 1 to 14 or Treatment Discontinuation (+1) ]

The investigator or subinvestigator will enroll patients who are confirmed to meet all the criteria specified in Section 4.2, "Inclusion Criteria" and not meet any of the criteria specified in Section 4.3, "Exclusion Criteria" and are therefore considered appropriate for the study, and initiate the study treatment after treatment allocation. Subjects meeting any of the criteria specified in Section 13.1, "Rules or Criteria for Treatment Discontinuation of Individual Patients" should discontinue the study treatment and move to the follow-up period. During the study period, subjects may be discharged from the study site if they meet the current criteria on hospital discharge specified by the Ministry of Health, Labour and Welfare, and had no safety issue, as determined by the investigator or subinvestigator. For subjects discharged from the study site before SARS-CoV-2 negative test is confirmed on 2 consecutive occasions, protocol-specified assessments should be continued on an outpatient basis until the subjects test negative for SARS-CoV-2 on 2 consecutive occasions.

##### 8.1.3 Follow-up Phase [14 Days after Last Dose (±3)]

For all subjects receiving the study treatment, the investigator or subinvestigator will collect information on any new adverse events until 14 (±3) days after the last dose of the study drug. During the study period, the patient may be discharged from the institution if there is no safety issue as determined by the investigator or subinvestigator. For patients discharged from the institution, the follow-up phase assessment may be performed via telephone.

After the completion of the assessment at the time of discontinuation, the follow-up should be continued as far as possible until treatment-related adverse events are considered as resolved/resolving, or no further follow-up is considered necessary because of symptom stabilization/permanent events.

#### 8.2 Observation Items

##### 8.2.1 Patient Characteristics

The following information will be entered into the eCRF: patient identification code, randomization number, date of randomization, date of written informed consent (by the legally acceptable representative, if applicable), sex, race, age, height,* weight,* date of onset of COVID-19 symptoms** (date of collection of the sample with a positive result for symptom-free patients), date of hospital admission, past history,*** concurrent illnesses,**** and prior treatment for COVID-19 (name of drug/therapy, initiation and completion dates, daily dose, and reason for use).

*Data collected during the observation phase should be used.

**The date of onset of COVID-19 symptoms is defined as the earliest date of onset of COVID-19-related symptoms reported by the subject.

***In addition to all information on past history within 1 year prior to enrollment, past history within >1 year prior to enrollment should be collected for diseases/illnesses that are specified in the inclusion or exclusion criteria, are clinically significant as determined by the investigator or subinvestigator, or affect the assessment of the causal relationship between adverse events and the study drug.

****Clinically significant diseases/illnesses or symptoms at baseline

##### 8.2.2 Status of Administration of the Study Drug

The status of administration of the study drug (initiation and completion dates, dose, and reason for treatment discontinuation if applicable) will be entered into the eCRF.

##### 8.2.3 Combination Therapy

Information on the use of all drugs and concomitant therapies between the period from baseline to the completion of protocol-specified assessments at Day 14 or treatment discontinuation (except for assessments for follow-up of adverse events) will be collected, and names of drugs/therapies, daily dose of each drug, treatment initiation and completion dates, and reason for use will be entered into the eCRF. For 14 days (Day 28 [±3]) after the completion of protocol-specified assessments at Day 14 or treatment discontinuation (except for assessments for follow-up of adverse events), information on concomitant medications and therapies used for adverse events will be collected.

##### 8.2.4 Efficacy Variables

For subjects who discontinue the study treatment, the investigator or subinvestigator will perform the assessments as far as possible with due consideration for their safety.

###### 8.2.4.1 SARS-CoV-2 Test

###### 8.2.4.1.1 SARS-CoV-2 Test (Local and Central Qualitative Test)

From Days 1 to 14 (or treatment discontinuation), the investigator or subinvestigator will perform SARS-CoV-2 tests (qualitative) at the study site, and enter the date of sample collection, test method, type of the sample, and test results into the eCRF. Each test for SARS-CoV-2 should be separated by at least 24 hours, except for Days 1 to 2, and the same test method and type of sample should be used throughout the study period as a general rule. If the patient tests negative for SARS-CoV-2 for the first time on Day 14, the patient should be re-tested for SARS-CoV-2 after an interval of at least 24 hours to confirm the negative SARS-CoV-2 test result.

From Days 1 to 14 (or treatment discontinuation), the investigator or subinvestigator will collect a sample for central SARS-CoV-2 testing, and enter the date of sample collection into the eCRF. The collected sample will be submitted to the central laboratory. As a general rule, the sample for central testing will be collected on consecutive days from Days 1 to 14 (or treatment discontinuation). If it is difficult to collect the sample on consecutive days, samples on at least Day 1 and the days on which the first and second SARS-CoV-2 negative tests are confirmed (by local testing) should be collected. Separate written procedures should be followed for the collection and submission of the sample to the central laboratory.

###### 8.2.4.1.2 SARS-CoV-2 Test (Central Quantitative Test)

From Days 1 to 14 (or treatment discontinuation), the investigator or subinvestigator will collect a sample for SARS-CoV-2 testing, and enter the date of sample collection into the eCRF. If the patient tests negative for SARS-CoV-2 for the first time on Day 14, the patient should be re-tested for SARS-CoV-2 after an interval of at least 24 hours. The collected sample will be submitted to the central laboratory. As a general rule, the sample for central testing will be collected on consecutive days from Days 1 to 14 (or treatment discontinuation). If it is difficult to collect the sample on consecutive days, samples on at least Day 1 and the days on which the first and second SARS-CoV-2 negative tests are confirmed (by local testing) should be collected. Separate written procedures should be followed for the collection and submission of the samples to the central laboratory.

###### 8.2.4.2 Ordinal Scale for Severity

From Days 1 to 14 (or treatment discontinuation), the investigator or subinvestigator will assess the ordinal scale for severity (see Appendix 1). The investigator or subinvestigator will enter the date and results of assessment into the eCRF.

###### 8.2.4.3 Presence/Absence of Clinical Symptoms

From Days 1 to 14 (or treatment discontinuation), the investigator or subinvestigator will assess the presence/absence of clinical symptoms. The investigator or subinvestigator will enter the date of assessment and the presence/absence of clinical symptoms into the eCRF.

###### 8.2.4.4 Use of Mechanical Ventilation

The investigator or subinvestigator will collect information on the use of mechanical ventilation (use/non-use of mechanical ventilation, and initiation and completion dates of mechanical ventilation if applicable) during the period from enrollment to Day 14 (or treatment discontinuation), and enter the information into the eCRF.

###### 8.2.4.5 Survival Status

The investigator or subinvestigator will collect information on the survival status (date of collection, date and cause of death [if applicable], and reason for unknown outcome and last date of survival [if applicable]) during the period from enrollment to Day 14 (or treatment discontinuation), and enter the information into the eCRF.

###### 8.2.4.6 Chest Imaging Tests

From Days 1 to 14 (or treatment discontinuation), the investigator or subinvestigator will perform a chest X-ray test, and enter the date of completion and presence/absence of abnormal lung findings into the eCRF.

###### 8.2.4.7 Antibody Test (Central)

On Days 1 and 14 (or treatment discontinuation), the investigator or subinvestigator will collect a sample for central antibody testing, and enter the date of sample collection into the eCRF. The collected sample will be submitted to the central laboratory. Separate written procedures should be followed for the collection and analysis of the samples.

##### 8.2.5 Safety Variables

For subjects who discontinue the study treatment, the investigator or subinvestigator will perform the assessments as far as possible with due consideration for their safety.

###### 8.2.5.1 Vital Signs (Body Temperature, $SpO_2$, Systolic/Diastolic Blood Pressure, Pulse Rate)

From Days 1 to 14 (or treatment discontinuation), the investigator or subinvestigator will measure body temperature, $SpO_2$, systolic/diastolic blood pressure, and pulse rate, and enter the date and results of measurement into the eCRF. The measurement on Day 14 should be performed after the study treatment.

###### 8.2.5.2 Laboratory Test

During the observation phase and on Days 7 and 14 (or treatment discontinuation), the investigator or subinvestigator will perform laboratory testing for the parameters below, and enter the date of sample collection and test results into the eCRF. Each test result will be compared with the baseline value, while determining whether it is normal or abnormal, to determine whether there is a clinically significant abnormal change from baseline. Parameters determined to show adverse events shall be described in detail in the eCRF, and the causal relationship to the study drug shall be assessed. The testing on Day 14 should be performed after the study treatment.

| Hematology | Blood biochemistry | Qualitative urinalysis |
|---|---|---|
| Red blood cell count | Blood glucose | pH |
| Hemoglobin | Total cholesterol | Protein |
| Hematocrit | Triglyceride | Sugar |
| White blood cell count | Total protein | Occult blood |
| Differential white blood count | Albumin | |
| - Neutrophils (segmented/stab cells) | BUN | |
| | Creatinine | |
| - Lymphocytes | Total bilirubin | |
| - Eosinocytes | AST (GOT) | |
| - Basocytes | ALT (GPT) | |
| - Monocytes | γ-GTP | |
| Blood platelet count | LDH | |
| D dimer | ALP | |
| | CK (CPK) | |
| | Na | |
| | K | |
| | Cl | |
| | Ca | |
| | CRP | |

Clinical Laboratory Test Reference Values

- Clinical laboratory test reference values will be entered into the eCRF.
- If clinical laboratory test reference values have not been defined, the investigator shall define them.

###### 8.2.5.3 12-Lead ECG

On Days 1, 7, and 14 (or treatment discontinuation), the investigator or subinvestigator shall record 12-lead ECG, and enter the date of completion and overall findings into the eCRF. For each test result and abnormal finding, the investigator or subinvestigator shall perform adverse event assessment for any clinically significant abnormal changes during the period as compared to the baseline condition, and enter the assessment result into the eCRF. The testing on Day 14 should be performed after the study treatment.

###### 8.2.5.4 Adverse Events

Information on adverse events will be collected through unsolicited reports from subjects, open-ended questions, various types of testing (e.g., laboratory tests, vital signs, ECG) and clinical assessment. Subjects will not be asked questions regarding solicited adverse events.

For adverse events reported during the period from baseline to the end of the follow-up phase (see Section 9.1, "Definition of Adverse Event"), the following information will be entered into the eCRF: event term, date of onset, severity (see Section 9.3, "Severity of Adverse Events"), judgment about study treatment continuation (see Section 9.4, "Judgment about Study Treatment Continuation"), action taken with the study drug, seriousness, causal relationship to the study drug (see Section 9.5, "Criteria for Assessing Causal Relationship to the Study Drug"), and outcome at the final assessment (or the date of resolution if the event resolved before the final assessment) (see Section 9.6, "Outcome of Adverse Events"). For adverse events based on objective measures such as laboratory test values, the relevant value at the onset of the adverse events and the worst value observed during follow-up will be entered in the eCRF. Regardless of the causal relationship to study drug, all adverse events will be treated appropriately while observing clinical manifestations and performing laboratory tests until the event is assessed as resolved, resolving, or requiring no further follow-up because of symptom stability or persistence. Serious adverse events (see Section 9.2, "Definition of Serious Adverse Event") will be reported to the head of the study site and the sponsor in accordance with Section 9.7, "Actions to Be Taken in the Case of Serious Adverse Events."

In principle, adverse events representing subjective symptoms/objective findings that the investigator or subinvestigator considers to be inextricably linked to abnormal laboratory changes shall be reported as single adverse events (only as adverse events representing subjective symptoms/objective findings). Subjective symptoms/objective findings and abnormal laboratory changes shall be reported separately as individual adverse events unless determined to be inextricably linked to each other.

### 9 ADVERSE EVENTS AND PATIENT SAFETY ASSURANCE

#### 9.1 Definition of Adverse Event

An adverse event is defined as any unfavorable and unintended sign (including an abnormal laboratory finding), symptom, or disease in a patient administered a study drug, regardless of relationship to the study drug. Exacerbation of the primary disease or accompanying symptoms or complications of the primary disease, which is medically assessed to be beyond the natural course of the disease, is regarded as an adverse event.

##### 9.2 Definition of Serious Adverse Event

A serious adverse event is defined as an adverse event meeting one of the conditions listed below.

1. Results in death
2. Is life-threatening
3. Requires inpatient hospitalization or prolongation of existing hospitalization [#1]
4. Results in persistent or significant disability/incapacity
5. Is a congenital anomaly
6. Other event that is judged to be a medically important condition [#2]

[#1]Among hospitalizations, hospitalization for the conduct of this study or the following hospitalization is not considered as a serious adverse event.

- Inpatient hospitalization or prolongation of existing hospitalization for treatment planned before the study (surgery or examinations planned before the start of the study, etc.)
- Inpatient hospitalization or prolongation of existing hospitalization for examinations or education (e.g., hospitalization for examination/education)
- Inpatient hospitalization or prolongation of existing hospitalization not resulting from worsening of concurrent diseases or concomitant symptoms but for surgery, etc. due to changes in treatment policy of concurrent diseases or concomitant symptoms
- Inpatient hospitalization or prolongation of existing hospitalization merely for follow-up of adverse events that have resolved or are resolving
- Hospitalization/admission to hospice, nursing-care facility or rehabilitation facility
- Inpatient hospitalization or prolongation of existing hospitalization for new treatment of primary disease such as next treatment following completion of study treatment
- Inpatient hospitalization or prolongation of existing hospitalization for social reasons (e.g., rest for caretaker, absent of family member)

[#2]As for other medically important events, that is, although an event does not result in immediate life-threatening condition, death, or hospitalization, if measures are needed to prevent the subject from being placed at risk or result in an outcome of the above 1 to 5, it is necessary to determine whether immediate reporting is required for such events based on the medical and scientific evidence, and the events should be assessed as serious. Examples of such events are intensive treatment in an emergency room, etc. for bronchospasm, blood dyscrasias or convulsions that do not result in hospitalization, development of drug dependency or drug abuse, or development of malignant tumor.

##### 9.3 Severity of Adverse Events

In reference to "Severity of adverse events" (see Appendix 2) and the following criteria, severity shall be assessed as 1 of 3 grades of mild, moderate, or severe.

| Severity | Assessment criteria |
|---|---|
| Mild | Signs or symptoms are observed but are easily tolerated (Grade 1) |
| Moderate | Discomfort that disturbs the activities of daily living (Grade 2) |
| Severe | Disability that prevents work or the activities of daily living (Grade 3) |

##### 9.4 Judgment about Study Treatment Continuation

For each reported adverse event, the investigator or subinvestigator will select his/her judgment about study treatment continuation from the 4 options below, and record the judgment. The subject's own judgment will not be taken into account.

| Investigator's judgment | Assessment criteria |
|---|---|
| Dose Not Changed | The investigator or subinvestigator determines that the study treatment may be continued. |
| Drug Withdrawn | The investigator or subinvestigator determines that the study treatment should be discontinued. |
| Not Applicable | The event occurred prior to the first dose of the study drug or after the last dose of the study drug. |
| Unknown | The investigator or subinvestigator's judgment about study treatment continuation is unknown. |

##### 9.5 Criteria for Assessing Causal Relationship to the Study Drug

The causal relationship between each adverse event and the study drug will be assessed as either 1 or 2, below, based on possible causes other than the study drug (e.g., the subject's underlying diseases, concomitant therapies, and other risk factors) and the temporal relationship between each adverse event and the study drug. Adverse events for which the causal relationship is determined to be 1. shall be handled as "adverse drug reactions." For 1., below, the basis for the determination shall be entered in the comments section in the eCRF.

1. Related: A valid causal relationship exists between administration of the study drug and the adverse event.
2. Unrelated: A valid causal relationship does not exist between administration of the study drug and the adverse event.

A "valid causal relationship" refers to the presence of evidence that suggests a causal relationship.

##### 9.6 Outcome of Adverse Events

The outcome of adverse events (symptoms and abnormal changes in laboratory test data) shall be assessed in accordance with the criteria below. In addition, the date of recovery in the case of "recovered," or the outcome at the time of last observation in the case of other outcomes shall be assessed.

| Outcome | Assessment criteria (reference only) |
|---|---|
| Recovered/Resolved | Disappearance of symptoms, normalization of test values, or recovery to baseline level |
| Recovering/Resolving | Alleviation of severity or tendency toward improvement of symptoms |
| Not recovered/Not resolved | The symptom or test value has not improved. |
| Recovered/Resolved with sequelae | Recovered with remained symptoms such as functional disturbances |
| Fatal | Results in death |
| Unknown | Unable to assess the outcome in the same manner as above, such as "outcome was impossible to follow-up" |

##### 9.7 Actions to Be Taken in the Case of Serious Adverse Events

If any serious adverse event that meets one of the definitions in Section 9.2, "Definition of Serious Adverse Event" occurs during the period from enrollment to 14 days after the last dose of the study drug, whether it is related to the study drug or not, the investigator or subinvestigator will provide appropriate treatment, and report it immediately (within 24 hours after becoming aware of the event) to the sponsor to provide information equivalent to that provided on the Uniform Format "Serious Adverse Event Report Form." The investigator or subinvestigator will also prepare a Serious Adverse Event Report Form that provides detailed information about the event (information equivalent to that provided on the Uniform Format "Serious Adverse Event Report Form" and the format for detailed information) within 3 days after becoming aware of the event, and report it by fax to the sponsor's contact person for safety management (see the contact information below) and submit the form to the head of the study site. When the investigator is not available, the subinvestigator may prepare and submit the form on behalf of the investigator. If receiving follow-up information, including any addition or change that may affect the safety assessment of the previously reported serious adverse event, the investigator will prepare a report as a second Serious Adverse Event Report Form within 3 days after receiving the follow-up information, and report it by fax to the sponsor's contact person for safety management and submit the form to the head of the study site.

In the case of additional information not considered to affect the assessment of a serious adverse event, such as the outcome of a reported event changing from “resolving” to “resolved,” it is not mandatory for the investigator to prepare a follow-up “Serious Adverse Event Report Form.”

The sponsor will discuss the contents of the report with medical experts and/or the Coordinating Committee, as required, and if submission of test images, etc. is requested by medical experts or the coordinating investigator for safety evaluation, masking of the personal information shall be performed before submission.

<Sponsor’s contact person for safety management>

Pharmacovigilance Division, Ono Pharmaceutical Co., Ltd.

TEL: 06-6222-5507 (to confirm the receipt of the Serious Adverse Event Report Form)

<Contact center to use if the sponsor is not available>

Emergency Contact Center (accessible 24 hours a day): BellSystem24, Inc.

#### 9.8 Actions to Be Taken if Pregnancy or Overdose Is Found

##### 9.8.1 Actions to Be Taken if Pregnancy Is Found

If a female subject becomes pregnant during the period from enrollment to 1 month after the last dose of the study drug (or discontinuation), the investigator or subinvestigator will report it to the sponsor orally or via telephone or FAX within 24 hours, as a general rule, after becoming aware of the pregnancy. Further, the subsequent study treatment shall be discontinued. Furthermore, the investigator or subinvestigator shall prepare a “Pregnancy Report Form” within 3 days after becoming aware of the pregnancy, and report it to the sponsor’s contact person for safety management by fax (see the contact information below). When the investigator is not available, the subinvestigator may prepare and submit the form on behalf of the investigator.

The investigator or subinvestigator should follow up the pregnancy until the outcome is determined, and should promptly complete an additional “Pregnancy Report Form” and report to the sponsor’s contact person for safety management by fax if the outcome is determined. Subsequently, if the investigator, etc. obtains information on abnormal development of a newborn, he/she shall report it to the sponsor.

If the partner of a male subject becomes pregnant during the period from enrollment to 1 month after the last dose of the study drug, the investigator or subinvestigator will, as in the case of a pregnancy of a female subject, collect as much information as possible about the partner's pregnancy, and prepare and submit a "Pregnancy Report Form" to the sponsor's contact person for safety management by fax.

<Sponsor's contact person for safety management>
Pharmacovigilance Division, Ono Pharmaceutical Co., Ltd.

<Contact center to use if the sponsor is not available>
Emergency Contact Center (accessible 24 hours a day): BellSystem24, Inc.

###### 9.8.2 Actions to Be Taken if Overdose Is Found

If a subject is found to have received an overdose, the investigator or subinvestigator must take appropriate measures and promptly report it to the sponsor verbally, via telephone, or by fax.

##### 9.9 Provision of New Information

If information that may adversely affect the safety of patients, may affect the conduct of the study, or alter the approval status by the Institutional Review Board of the continuation of the study is obtained, or information on unexpected serious adverse drug reactions is obtained, the sponsor shall promptly notify all investigators and the heads of the study sites involved in this study in writing. The investigator should promptly amend the Informed Consent Form and Written Information as necessary. Further, the sponsor shall amend the protocol/Investigator's Brochure as needed.

##### 9.10 Emergency Code Breaking on Emergency

If it becomes necessary to know the allocation of the study drug, e.g., due to onset of a serious adverse event, the emergency code shall be opened in accordance with Section 5.3.3, "Code-Breaking Procedure."

#### 10 STATISTICAL ANALYSIS

Statistical analysis will be performed by Ono Pharmaceutical Co., Ltd. The basic policy on analyses is presented below. Detailed methods of analyses will be provided in a Statistical Analysis Plan prepared by the Biostatistics Director and the biostatistician. An Interim Analysis Plan will be prepared as appropriate.

The first versions of the interim and statistical analysis plans will be finalized prior to data lock for the interim analysis if the interim analysis is performed, or prior to data lock if the interim analysis is not performed. After the first versions are finalized, the interim and statistical analysis plans will be revised as appropriate. Any additional analysis not described in the Statistical Analysis Plan will be reported in a clinical study report.

#### 10.1 Determination of the Sample Size

110 subjects in total, with 55 per group

<Rationale>

The objective of this study is to establish the superiority of the FOY-305 group to the placebo group in patients with COVID-19 using the time to negative test for SARS-CoV-2 as assessed by the local laboratory as the primary endpoint. Subjects will be randomized in a 1:1 ratio to receive FOY-305 or placebo through a dynamic allocation procedure, with site, age (≥65 vs <65 years), and underlying disease (chronic respiratory disease, chronic kidney disease, diabetes mellitus, hypertension, cardiovascular diseases, obesity [BMI ≥30 kg/$m^2$]) (present vs absent) as allocation factors.

When further enrollment appears to be difficult because of convergence of the COVID-19 pandemic during the subject enrollment period of this study, an interim analysis will be performed to determine whether the treatment is effective or ineffective as of the time. Bayesian interim monitoring[19)] is applied to the interim analysis, and the criteria for assessment in the interim and final analyses are shown in the table below. Note that the criteria for assessment in the final analysis will remain the same even if no interim analysis takes place.

| Time of analysis | Criteria for effectiveness | Criteria for ineffectiveness |
|---|---|---|
| Interim analysis | Bayesian posterior probability ≥97.5% for hazard ratio >1.0 | Bayesian posterior probability ≤2.5% for hazard ratio >1.0 AND Bayesian posterior probability ≥80.0% for hazard ratio <1.4 |
| Final analysis | Bayesian posterior probability ≥92% for hazard ratio >1.0 | Bayesian posterior probability ≤8% for hazard ratio >1.0 |

The time to negative test for SARS-CoV-2 was assumed to follow an exponential distribution, with the median time assumed to be 14 days in the placebo group. The median time of 7 or 8 days was assumed in the FOY-305 group, and the probabilities of meeting the assessment of effectiveness or ineffectiveness with 50 subjects per group (100 in total) were calculated for various analysis time points through numerical simulations with the SAS statistical analysis software (version 9.4). As the timing of the interim analysis depends on the status of the COVID-19 outbreak, numerical simulations were performed, assuming the time points where 40 to 80 subjects have been randomized. The prior distribution of the regression coefficient was assumed to be uniform distribution.

Numerical experiments were repeated 1,000 times under the above-mentioned conditions, and the probabilities of meeting the assessment of effectiveness and ineffectiveness at various time points were estimated as shown below:

| Median value (days) in the FOY-305 group | Number of subjects at the time of interim analysis | Assessment | Interim analysis (%) | Final analysis (%) | Total (%) |
|---|---|---|---|---|---|
| 7 | 40 | Effective | 36.6 | 52.2 | 88.8 |
| | | Ineffective | 4.3 | 6.9 | 11.2 |
| | 50 | Effective | 44.1 | 46.2 | 90.3 |
| | | Ineffective | 2.4 | 7.3 | 9.7 |
| | 60 | Effective | 51.1 | 39.1 | 90.2 |
| | | Ineffective | 2.2 | 7.6 | 9.8 |
| | 70 | Effective | 57.5 | 32.5 | 90.0 |
| | | Ineffective | 1.5 | 8.5 | 10.0 |
| | 80 | Effective | 64.3 | 25.9 | 90.2 |
| | | Ineffective | 1.5 | 8.3 | 9.8 |
| 8 | 40 | Effective | 23.1 | 52.2 | 75.3 |
| | | Ineffective | 7.6 | 17.1 | 24.7 |
| | 50 | Effective | 29.8 | 45.5 | 75.3 |
| | | Ineffective | 6.5 | 18.2 | 24.7 |
| | 60 | Effective | 36.1 | 40.6 | 76.7 |
| | | Ineffective | 6.1 | 17.2 | 23.3 |
| | 70 | Effective | 39.1 | 38.0 | 77.1 |
| | | Ineffective | 4.6 | 18.3 | 22.9 |
| | 80 | Effective | 45.5 | 31.1 | 76.6 |
| | | Ineffective | 4.9 | 18.5 | 23.4 |
| 14 | 40 | Effective | 2.4 | 6.9 | 9.3 |
| | | Ineffective | 43.6 | 47.1 | 90.7 |
| | 50 | Effective | 2.6 | 6.6 | 9.2 |
| | | Ineffective | 47.2 | 43.6 | 90.8 |
| | 60 | Effective | 2.8 | 6.4 | 9.2 |
| | | Ineffective | 49.2 | 41.6 | 90.8 |
| | 70 | Effective | 2.2 | 6.8 | 9.0 |
| | | Ineffective | 53.7 | 37.3 | 91.0 |
| | 80 | Effective | 2.3 | 6.2 | 8.5 |
| | | Ineffective | 56.0 | 35.5 | 91.5 |

The results of the numerical simulations showed that the probability of the study treatment to be assessed as effective was around 75% to 90% in total when the median time to negative test in the FOY-305 group is 7 or 8 days, irrespective of which of the assumed time points the interim analysis is performed at. With the median time of 14 days in the FOY-305 group, the probability of the study treatment to be assessed as effective was generally controlled within 10%. It was therefore considered possible to demonstrate the superiority of the FOY-305 group to the placebo group with 50 subjects per group (100 subjects in total).

The target number of subjects was set at 110, taking into consideration the number of subjects to have negative SARS-CoV-2 test on Day 1 (as assessed by the local laboratory).

<Rationale for assuming the median time to negative test for SARS-CoV-2 in the placebo group to be 14 days>

Epidemiological studies on changes over time in the SARS-CoV-2 viral load in patients with COVID-19 have shown that the time from onset of COVID-19 symptoms to spontaneous resolution of SARS-CoV-2 viral load was 20 days on average.[17), 18)] In consideration of the time from onset of COVID-19 symptoms to enrollment in this study along with the above data, the median time to negative test for SARS-CoV-2 in the placebo group was assumed to be 14 days.

#### 10.2 Procedures for Statistical Analyses, Including the Timing of Planned Interim Analyses

When further enrollment appears to be difficult because of convergence of the COVID-19 pandemic during the subject enrollment period of this study, an interim analysis will be performed to determine whether the treatment is effective or ineffective based on the hazard ratio of the FOY-305 group to the placebo group. The hazard ratio will be estimated in a Cox proportional hazards model using treatment as a single covariate, stratified by allocation factors of age and underlying disease (chronic respiratory disease, chronic kidney disease, diabetes mellitus, hypertension, cardiovascular disease, obesity [BMI ≥30 kg/m$^2$]). Bayesian interim monitoring[19)] is applied to the interim analysis, and the criteria for assessment in the interim and final analyses are shown in the table below. Note that the criteria for assessment in the final analysis will remain the same even if no interim analysis takes place.

| Time of analysis | Criteria for effectiveness | Criteria for ineffectiveness |
|---|---|---|
| Interim analysis | Bayesian posterior probability ≥97.5% for hazard ratio >1.0 | Bayesian posterior probability ≤2.5% for hazard ratio >1.0 AND Bayesian posterior probability ≥80.0% for hazard ratio <1.4 |
| Final analysis | Bayesian posterior probability ≥92% for hazard ratio >1.0 | Bayesian posterior probability ≤8% for hazard ratio >1.0 |

An independent data monitoring committee (IDMC) will assess the treatment as effective or ineffective in the interim analysis. Details of the interim analysis are described in the separate written procedure for the IDMC and the interim analysis plan.

#### 10.3 Level of Significance to Be Used

Two-sided tests with a significance level of 5% shall be used. No significance level will be determined, since the primary analysis of the primary efficacy endpoint will be based on Bayesian interim monitoring. For other endpoints and time points, no multiplicity adjustment will be made.

#### 10.4 Inclusion of Subjects in Analysis Populations

##### 10.4.1 Definition of Analysis Sets

The Modified Intention-to-Treat (mITT) will be used for efficacy analysis.

The SAF will be used for safety analysis.

The definition of each analysis population is presented below.

###### 10.4.1.1 Informed Consent Set (ICS)

The Informed Consent set is defined as the group of patients who have provided informed consent.

###### 10.4.1.2 Randomized Set (RND)

The Randomized set is defined as the group of subjects who are randomized.

###### 10.4.1.3 Modified Intention-to-Treat (mITT)

The mITT is defined as the maximum group of subjects included in the RND who can be analyzed for efficacy. Subjects in the mITT must meet the following criterion:

1. Tested positive for SARS-CoV-2 on Day 1 (as assessed by the local laboratory)

###### 10.4.1.4 Safety Set (SAF)

The SAF is defined as the group of subjects who have received at least one dose of the study drug without GCP noncompliance specified in Section 10.4.2, “Criteria for Handling Subjects.”

##### 10.4.2 Criteria for Handling Subjects

Details of handling subjects are explained below.

###### 10.4.2.1 Pre-Randomization Dropouts

A pre-randomization dropout is defined as a subject who has never been randomized after providing informed consent.

###### 10.4.2.2 Subjects Testing Negative for SARS-CoV-2

A subject testing negative for SARS-CoV-2 is defined as a subject testing negative for SARS-CoV-2 on Day 1 (as assessed by the local laboratory).

###### 10.4.2.3 GCP-Noncompliant Subjects

A GCP-noncompliant subject is defined as a subject from whom data has not been reviewed by the Institutional Review Board, who has not concluded the clinical study contract, or who has not provided appropriate informed consent.

###### 10.4.2.4 Non-Treated Subjects

Non-treated subjects are defined as subjects who have not received any dose of the study drug.

The sponsor should discuss unexpected cases other than those listed above with the medical expert, and decide how to handle these subjects in analyses prior to data lock for each analysis.

##### 10.4.3 Criteria for Handling of Evaluation Timing

If the protocol-specified and actual dates of assessment differ, only data collected within the acceptable window specified in Table 8-1 in the Synopsis will be used for the evaluation of each test parameter.

##### 10.4.4 Handling of Missing Measurements

Explicit supplementation of missing measurements, e.g., substitution with another measurement, shall not be performed in any analysis set, unless particularly specified otherwise. During a blinded review, missing data status will be checked, and the handling of missing data will be discussed as appropriate for performing an exploratory analysis.

#### 10.5 Analytical Methods

##### 10.5.1 Examination of the Reliability of the Study

###### 10.5.1.1 Presence or Absence of Excluded Subjects

Frequencies of subjects included in or excluded from analysis sets and frequencies of subjects excluded from analysis sets by reason for exclusion will be summarized for each treatment group.

###### 10.5.1.2 Status of Discontinuation/Dropouts

Frequency of subjects who discontinue the study treatment and frequency by reason for discontinuation will be summarized for each treatment group. Frequency of subjects who discontinue the study and frequency by reason for study discontinuation will be summarized for each treatment group.

##### 10.5.2 Status of Administration of the Study Drug

The number of tablets taken will be tabulated for each treatment group.

##### 10.5.3 Distribution of Demographics and Baseline Characteristics

The frequency distribution and summary statistics for demographic variables and patient clinical characteristics will be calculated by treatment group, and intergroup comparability will be determined.

##### 10.5.4 Primary Efficacy Endpoint

###### 10.5.4.1 Objective

The objective of this study involving patients with COVID-19 is to demonstrate the superiority of the FOY-305 group over the placebo group in terms of the time to negative test for SARS-CoV-2 as assessed by the local laboratory (primary endpoint).

###### 10.5.4.2 Hypothesis

The time to negative test for SARS-CoV-2 is shorter in the FOY-305 group than in the placebo group.

###### 10.5.4.3 Analysis Variables and Data Handling

- Endpoint

  Time to negative test for SARS-CoV-2 as assessed by the local laboratory

- Handling of Data

  The time to negative test for SARS-CoV-2 will be calculated using the following formula:

Time to negative test for SARS-CoV-2 (days) = “Date of collection of the first sample testing negative for SARS-CoV-2” − “Date of randomization” + 1

The confirmation of SARS-CoV-2 negative test will require two consecutive negative results from local laboratory SARS-CoV-2 testing (obtained after an interval of at least 24 hours), and the date of collection of the sample for the first test will be used as the date of collection of the first sample testing negative for SARS-CoV-2.

Depending on the situation, the determination on an event or censoring will be made according to Table 10.5-1. For subjects for whom more than one event was censored, the longest period of time will be used as the duration for the subjects. Although it is not expected that there will be many subjects who test positive for SARS-CoV-2 after becoming negative for SARS-CoV-2 or whose symptoms become severe, a sensitivity analysis will be performed as appropriate to investigate the effects of such subjects.

**Table 10.5-1 Definition of an Event or Censoring in the Time to Negative Test for SARS-CoV-2**

| Status | Date of an event or censoring | Determination |
|---|---|---|
| SARS-CoV-2 negative test has been confirmed | Date of the collection of the first sample testing negative for SARS-CoV-2 (by local laboratory testing) | Event |
| SARS-CoV-2 negative test has not been confirmed | Date of the collection of the sample with the final evaluable SARS-CoV-2 test result (from local laboratory testing) | Censoring |
| The subject has died | Day 14 | Censoring |
| The subject meets Criterion 6 or 7 of the criteria for discontinuation of study treatment, both of which are related to increased severity of COVID-19, and therefore has discontinued the study treatment | Day 14 | Censoring |
| The subject has discontinued the study | Date of the collection of the sample with the final evaluable SARS-CoV-2 test result (from local laboratory testing) | Censoring |
| Local SARS-CoV-2 testing has not been performed at 2 or more time points after randomization | Day 1 | Censoring |

##### 10.5.4.4 Analysis Method

1. Primary Analysis Methods

The posterior mean hazard ratio and corresponding two-sided 95% credible interval will be estimated in a Cox proportional hazards model using treatment as a single covariate, stratified by allocation factors of age and underlying diseases (chronic respiratory disease, chronic kidney disease, diabetes mellitus, hypertension, cardiovascular disease, and obesity [BMI ≥30 kg/m$^2$]). In addition, the Bayesian posterior probability for hazard ratio >1.0 and the Bayesian posterior probability for hazard ratio <1.4 (only for the interim analysis) will be calculated. The prior distribution of the regression coefficient is assumed to be uniform distribution.

2. Secondary Analytical Methods

A stratified log-rank test using allocation factors, including age and underlying diseases (chronic respiratory disease, chronic kidney disease, diabetes mellitus, hypertension, cardiovascular diseases, and obesity [BMI ≥30 kg/m$^2$]), as stratification factors, will be performed to compare the treatment groups. The Kaplan-Meier curves for each treatment group will be presented, and the medians and their two-sided 95% confidence intervals will be calculated using the Kaplan-Meier method. For the calculation of the confidence intervals using the Kaplan-Meier method, the Brookmeyer and Crowley method with double-log transformation will be used.

##### 10.5.5 Secondary Efficacy Endpoints

###### 10.5.5.1 Objective

To analyze the non-primary endpoints in an exploratory manner so as to evaluate the pharmacological effect of FOY-305 from multiple aspects

###### 10.5.5.2 Analysis Variables and Data Handling

###### 10.5.5.2.1 Analysis Items

1. Time to negative test for SARS-CoV-2 as assessed by the central laboratory
2. Proportion of subjects who test negative for SARS-CoV-2 (as assessed by the local and central laboratories)
3. Ordinal scale for severity
4. Proportion of subjects on mechanical ventilator
5. Survival status

###### 10.5.5.2.2 Handling of Data

SARS-CoV-2 negative test will be handled in the same manner as for the primary efficacy endpoint.

The survival status (overall survival [days])will be calculated using the following formula: “Date of death by any cause” − “Date of randomization” + 1. For subjects lost to follow-up or who survived to Day 14, the date of final survival confirmation will be used as the date of censoring.

###### 10.5.5.3 Analysis Method

1. For Analysis Item 1, data will be analyzed in the same manner as for the primary efficacy endpoint.
2. For Analysis Item 2, the frequency distribution will be calculated for each assessment time point and treatment group, and the two-sided 95% confidence interval of the percentage will be calculated using the Clopper-Pearson method. The treatment group difference in the percentage and its two-sided 95% confidence interval will be calculated using the Mantel-Haenszel estimator stratified by the allocation factors, including age and underlying diseases (chronic respiratory disease, chronic kidney disease, diabetes mellitus, hypertension, cardiovascular diseases, and obesity [BMI ≥30 kg/$m^2$]).
3. For Analysis Item 3, the frequency distributions of the actual measurement and the change from Day 1 will be calculated for each assessment time point and treatment group. In addition, the treatment group difference in the percentage of each category and its two-sided 95% confidence interval will be calculated using the Mantel-Haenszel estimator stratified by the allocation factors, including age and underlying diseases (chronic respiratory disease, chronic kidney disease, diabetes mellitus, hypertension, cardiovascular diseases, and obesity [BMI ≥30 kg/$m^2$]). The actual measurement will be compared between the treatment groups (the odds ratio of the FOY-305 group to the placebo group and its two-sided 95% confidence interval), using a proportional odds model with treatment group and the allocation factors, including age and underlying diseases (chronic respiratory disease, chronic kidney disease, diabetes mellitus, hypertension, cardiovascular diseases, and obesity [BMI ≥30 kg/$m^2$]).
4. For Analysis Item 4, the frequency distribution will be calculated for each treatment group, and the two-sided 95% confidence interval of the percentage will be calculated using the Clopper-Pearson method. The treatment group difference in the percentage and its two-sided 95% confidence interval will be calculated using the Mantel-Haenszel estimator stratified by the allocation factors, including age and underlying diseases (chronic respiratory disease, chronic kidney disease, diabetes mellitus, hypertension, cardiovascular diseases, and obesity [BMI ≥30 kg/$m^2$]).
5. For Analysis Item 5, the hazard ratio and corresponding two-sided 95% confidence interval will be estimated in a Cox proportional hazards model using treatment as a single covariate, stratified by allocation factors of age and underlying disease (chronic respiratory disease, chronic kidney disease, diabetes mellitus, hypertension, cardiovascular disease, obesity [BMI ≥30 kg/$m^2$]).The Kaplan-Meier curves for each treatment group will be presented, and the medians and their two-sided 95% confidence intervals will be calculated using the Kaplan-Meier method. For the

calculation of the confidence intervals using the Kaplan-Meier method, the Brookmeyer and Crowley method with double-log transformation will be used.

##### 10.5.6 Exploratory Efficacy Endpoints

###### 10.5.6.1 Objective

To analyze the non-primary endpoints in an exploratory manner so as to evaluate the efficacy of FOY-305 from multiple aspects

###### 10.5.6.2 Analysis Variables and Data Handling

###### 10.5.6.2.1 Analysis Items

1. SARS-CoV-2 viral load (as measured by the central laboratory)
2. Presence or absence of lung lesion as observed by chest imaging
3. Time to resolution of clinical symptoms and proportion of subjects in whom clinical symptoms have resolved
4. Antibody response (IgM and IgG) (measured by the central laboratory)

###### 10.5.6.2.2 Handling of Data

The time to resolution of clinical symptoms (days) will be calculated using the following formula: "Date of the first confirmed resolution of clinical symptoms" − "Date of randomization" + 1. For the time to resolution of clinical symptoms, the detailed procedure for determining an event or censoring will be provided in the Statistical Analysis Plan.

###### 10.5.6.3 Analysis Method

1. For Analysis Items 1 and 4, summary statistics will be calculated by assessment time point and treatment group.
2. For Analysis Items 2 and 3 (percentage of subjects with clinical symptoms resolved), the frequency distribution will be calculated for each treatment group, and the two-sided 95% confidence interval of the percentage will be calculated using the Clopper-Pearson method.
3. For Analysis Item 3 (time to resolution of clinical symptoms), the Kaplan-Meier curves for each treatment group will be presented, and the medians and their two-sided 95% confidence intervals will be calculated using the Kaplan-Meier method. For the calculation of the confidence intervals using the Kaplan-Meier method, the Brookmeyer and Crowley method with double-log transformation will be used.

##### 10.5.7 Adjustment Analysis

If there are any background factors that indicate bias among treatment groups or may have an impact on the evaluation, adjusted analysis with regard to the primary endpoint will be conducted with these as adjustment factors.

##### 10.5.8 Safety Analysis

###### 10.5.8.1 Objective

The objective is to evaluate the safety of FOY-305.

###### 10.5.8.2 Analysis Method

1. The numbers of subjects with adverse events and drug-related adverse events will be tabulated by treatment group.
2. Adverse events and drug-related adverse events will be tabulated by system organ class (SOC), PT, severity, and treatment group.
3. Regarding laboratory test data, summary statistics will be calculated for items of continuous values by assessment timing and by treatment group, and frequency distribution will be calculated for items of ordinal scale. In addition, the proportion of abnormal laboratory test data will be calculated.
4. Regarding vital signs, summary statistics will be calculated by assessment timing and by treatment group.
5. Regarding ECG, the proportion of abnormal findings will be calculated by assessment timing and treatment group.

##### 10.5.9 Blinded Review

Once data is accumulated, a preliminary review of the distribution of demographics and baseline characteristics will be performed using blinded data before data lock (i.e., data from subjects whose treatment allocation has not been unblinded) to confirm the appropriateness of the planned analytical methods.

##### 10.5.10 Other Variables

Subgroup analyses shall be performed on efficacy and safety endpoints.

In addition, exploratory analyses will be performed for other items related to the efficacy and safety, as required.

### 11 ETHICAL CONSIDERATIONS FOR THE CLINICAL STUDY

#### 11.1 Institutional Review Board

Prior to the implementation of the study, the Institutional Review Board of each study site will review the contents of the protocol, the Investigator's Brochure, and the Informed Consent Form and Written Information for patients, as well as the appropriateness of the conduct of the study.

#### 11.2 Continuing Review

1. To facilitate the continuing review by the Institutional Review Board, the investigator should submit written summaries of the study status to the head of the study site annually, or more frequently as requested by the Institutional Review Board.
2. The head of the study site should seek the opinion of the Institutional Review Board on the continuation of the study when he/she is notified of the study status at the study site, or if informed of unexpected serious adverse drug reactions or other issues by the sponsor, of serious adverse events by the investigator, of amendments made to Written Information by the investigator based on information that may affect the patient's willingness to continue participation in the study, or if he/she considers it necessary.

#### 11.3 Preservation of Patient Confidentiality

Due consideration shall be given to the preservation of patients' confidentiality. A patient identification code shall be used instead of the patient's name to identify the patient in the eCRFs and other study records. The eCRFs prepared will not be used for purposes other than this study. Any information that the sponsor becomes aware of shall not be disclosed to a third party.

### 12 OBTAINING INFORMED CONSENT FROM PATIENTS

#### 12.1 Informed Consent Form and Written Information

The investigator shall prepare Written Information and Informed Consent Forms that will be used to obtain patients' consent for participating in the study, in cooperation with the sponsor, and shall amend these if necessary. These documents that are prepared or amended shall be submitted to the sponsor.

Prior to the use of these documents, the investigator must obtain notification of the approval for use of these documents from the head of the study site based on the approval of the Institutional Review Board.

#### 12.2 Timing and Method of Obtaining Consent and its Contents

##### 12.2.1 Informed Consent Procedures

In this study, enrolled subjects will be randomized to study treatment to start the treatment phase. The investigator or subinvestigator will hand deliver a separate written document that defines the contents shown in Section 12.3.1, "Written Information for Patients," (Informed Consent Form and Written Information approved by the Institutional Review Board of each study site in advance) to the patient him/herself or legally acceptable representative (see Table 2 List of Terms") prior to enrollment, explain sufficiently, and after confirming that the patient has understood its contents, obtain consent based on the free will of the patient him/herself to participate in this study in a written document.

The patient shall personally sign and enter the consent date on the Informed Consent Form. The investigator or subinvestigator who provided an explanation shall sign and enter date of consent confirmation on the Informed Consent Form. If a study collaborator provides a supplementary explanation, the study collaborator shall also sign and enter the date of consent confirmation on the form. The investigator or subinvestigator will provide the subject with a copy of the signed Informed Consent Form together with the Written Information, and appropriately store the original in accordance with the rules of the study site rule.

The Written Information shall state any possibility that data from tests or assessments performed prior to obtaining informed consent shall be used as data for this study.

#### 12.3 Description of Written Information

##### 12.3.1 Written Information for Patients

1. That the study is associated with research
2. The objectives of the clinical study
3. The name, title, and contact information of the investigator or subinvestigator
4. Study methods (including the research aspects of the study, patient inclusion criteria, and probability of being assigned to each treatment if random allocation is performed)
5. The expected clinical benefits and risks or inconveniences (if there is no intended clinical benefit to the patient, the patient should be made aware of this)
6. When asking patients to participate in the clinical study as subjects, availability of alternative treatments for the patient and important potential benefits and risks of these treatments

7. The expected duration of the patient's participation in the clinical study
8. That the patient's participation in the clinical study is voluntary and that the patient or legally acceptable representative may refuse to participate or withdraw from the clinical study at any time. That there will be no penalty or loss of benefits to which the patient is otherwise entitled for refusing to participate or withdrawing from the clinical study.
9. That the monitors, the auditors, the Institutional Review Board, and the regulatory authorities will have access to the source data related to medical care. That in such a case, the confidentiality of patients will be maintained. In addition, by writing his or her name and affixing a name seal on or signing the Informed Consent Form, the patient or legally acceptable representative is authorizing such access.
10. That the patient's privacy will remain confidential even if the results of the clinical study are published.
11. The contact information of the study site for further information regarding the clinical study and the rights of patients, and for inquiries or contact in the event of study-related injuries.
12. The compensation and/or treatment available to the patient in the event of study-related injuries
13. The planned number of patients involved in the clinical study
14. That the patient or legally acceptable representative will be informed in a timely manner if information becomes available that may be relevant to the patient's or legally acceptable representative's willingness to continue participation in the clinical study.
15. The foreseeable circumstances and/or reasons under which the patient's participation in the study may be terminated
16. The anticipated expenses, if any, to the patient for participating in the clinical study
17. The anticipated prorated payment, if any, to the patient for participating in the clinical study (arrangements for prorated payment)
18. The patient's responsibilities
19. The type of Institutional Review Board that investigates and reviews the appropriateness of the clinical study; the matters that are investigated and reviewed by each Institutional Review Board; and other matters regarding the Institutional Review Board in relation to the clinical study

#### 12.4 Considerations for Informed Consent

In principle, the patient shall write his or her name and affix their name seal on or sign the Informed Consent Form.

- If the patient or legally acceptable representative cannot read the Informed Consent Form, Written Information, or other documents

  If the patient or legally acceptable representative is not able to read the Written Information, but is able to understand information provided verbally or by other means of communication, an impartial witness (see Table 2 List of Terms”) must be present during the explanation. After the patient or legally acceptable representative is provided with the Written Information and given an explanation of the information verbally or by other means of communication, the patient or legally acceptable representative shall write his or her name and affix their name seal on or sign the Informed Consent Form, and date the form to indicate consent to participate in the study. Subsequently, the witness shall write his or her name and affix their name seal on or sign, and write the date in the margin of the Informed Consent Form.
- For patients who are underage (<20 years old):

  For patients who are underage, written informed consent will also be obtained from their legally acceptable representatives.

#### 12.5 When Any Information That May Affect the Willingness of Subjects Is Obtained

- If any information that may affect the patient’s or legally acceptable representative’s willingness to continue to participate in the study is obtained (information considered related to the participating patient’s willingness to participate, for instance, information about the course of the study, such as changes in laboratory test values of an individual patient), the investigator or subinvestigator shall immediately inform the patient or legally acceptable representative and confirm their willingness to continue to participate in the study. Furthermore, the date that the patient or legally acceptable representative is informed, the content of the information, and the confirmation result shall be recorded in a source document such as the medical record.
- If any new important information that may affect the patient’s consent (i.e., information considered important for all patients that may require amendment of the Informed Consent Form and Written Information) is obtained, the investigator shall immediately amend the Written Information based on the relevant information, and obtain notification of approval to use this document from the head of the study site, based on the prior approval by the Institutional Review Board. Furthermore, the investigator or subinvestigator shall inform patients already participating in the study (or their legally acceptable representatives) in a timely manner to confirm their willingness to continue participation in the study, and shall provide an explanation to these patients or their legally acceptable representatives using the amended Written Information to obtain the written voluntary

informed consent of the patients or legally acceptable representatives for continuing participation in the study. The date of informed re-consent will be entered into the eCRF. It is, however, not necessary to obtain informed consent from patients for whom assessments have been completed at the time of the amendment.

##### 12.6 Withdrawal of Consent

Subjects who request study treatment discontinuation will move to the protocol-specified follow-up phase. If the subject or legally acceptable representative withdraws consent to follow-up, the investigator or subinvestigator should be notified **in writing** whenever possible. With regard to whether the consent withdrawal is applied to subsequent study treatment only or to the follow-up after the completion of study procedures and/or study treatment, the investigator or subinvestigator will record it in the source documents (e.g., medical records) and enter it into the eCRF. If the survival status (whether the subject is alive or dead) is confirmed based on official information, local guidelines should be followed in compliance with local laws.

#### 13 RULES OR CRITERIA FOR DISCONTINUATION OF INDIVIDUAL PATIENTS, PARTS OF THE STUDY, AND THE ENTIRE STUDY

##### 13.1 Rules or Criteria for Treatment Discontinuation of Individual Patients

###### 13.1.1 Observation Period

If any of the following conditions is met, the subject shall not be transferred to the treatment period:

1. The subject has requested for withdrawal from the study
2. The legally acceptable representative has requested for withdrawal of the subject from the study
3. The subject is found not to meet the inclusion criteria or to meet the exclusion criteria
4. Continuation of the study is not appropriate for any other reasons, in the opinion of the investigator or subinvestigator

###### 13.1.2 After the Initiation of the Treatment Phase

###### 13.1.2.1 Discontinuation of Study Treatment

If any one of the criteria listed below is met, the investigator or subinvestigator should immediately discontinue study treatment, take appropriate action, and observe clinical symptoms and perform clinical laboratory tests for the time of treatment discontinuation whenever possible. The investigator or subinvestigator should enter the date of completion of the treatment phase (i.e., date of completion of assessments for the treatment phase) and the reason for discontinuation into the eCRF, and report them to the sponsor promptly.

1. The subject has requested treatment discontinuation
2. The legally acceptable representative has requested treatment discontinuation
3. The subject is found not to meet the inclusion criteria or to meet the exclusion criteria
4. Continuation of the study treatment is difficult because of adverse event(s), whether or not related to the study drug, in the opinion of the investigator or subinvestigator
5. The subject tests negative for SARS-CoV-2 on 2 consecutive occasions
6. When both of the criteria a. and b. are met, and continuation of the study is considered difficult:
   a. Exacerbation of pneumonia is observed by chest imaging
   b. $SpO_2$ is ≤93% despite oxygen therapy
7. The subject receives any prohibited concomitant medication
8. The subject is dead
9. The subject is found to be pregnant
10. Continuation of the study is not appropriate for any other reasons, in the opinion of the investigator or subinvestigator

###### 13.1.2.2 Study Withdrawal

If any one of the criteria listed below is met, the investigator or subinvestigator should immediately withdraw the subject from the study, take appropriate action, and observe clinical symptoms and perform clinical laboratory tests for the time of treatment discontinuation whenever possible. The investigator or subinvestigator should enter the date of completion of the treatment phase (i.e., date of completion of assessments for the treatment phase), the date of study completion and the reason for discontinuation into the eCRF, and report them to the sponsor promptly.

1. The subject has requested for withdrawal from the study
2. The legally acceptable representative has requested for withdrawal of the subject from the study
3. Continuation of the study is difficult because of adverse event(s), whether or not related to the study drug, in the opinion of the investigator or subinvestigator
4. The subject is dead
5. The subject is found to be pregnant
6. Continuation of the study is not appropriate for any other reasons, in the opinion of the investigator or subinvestigator

###### 13.1.2.3 Lost to Follow up

If the subject repeatedly misses study visits as scheduled, and the study site cannot reach the subject, the subject will be considered as lost to follow up.

If the subject fails to return to the study site as scheduled, the following actions need to be taken:

- The study site will contact the subject as promptly as possible to reschedule the missed visit, explain the importance of following the visit schedule, and determine whether the subject wishes to and should continue the study.
- Before considering the subject as lost to follow up, the investigator or subinvestigator will again make the best effort to reach the subject (by making 3 telephone calls if possible, and sending a notice by certified mail to the most recently confirmed address as appropriate). The measures taken to reach the subject should be recorded in the source documents (e.g., medical records).
- If the subject still cannot be reached, the subject will be considered as dropped out of the study.

#### 13.2 Premature Termination or Suspension of the Study at a Study Site

1. If the investigator has prematurely terminated or suspended the study, the investigator should promptly notify the head of the study site in writing and explain the reason in detail.
2. If the head of the study site is notified by the investigator of the premature termination or suspension of the study, the head of the study site should promptly notify the sponsor and the Institutional Review Board in writing and explain the reason in detail.

#### 13.3 Rules and Criteria for Discontinuation of Part of the Study or the Entire Study

1. If the sponsor obtains information that may adversely affect the safety of subjects or the conduct of the study, or alter the approval status by the Institutional Review Board of the continuation of the study, the sponsor should discuss with the medical expert, and if necessary, with the coordinating investigator, to determine whether to discontinue the study partially or entirely.
2. The sponsor may be forced to discontinue the study at its discretion for any reason.
3. If the sponsor makes a determination to discontinue the study partially or entirely, the sponsor should promptly notify all investigators, the heads of the study sites involved in this study, and regulatory authorities in writing of the discontinuation of the study and detailed reasons for the discontinuation.

#### 14 COMPLETION OF THE STUDY

After completion of the study, the investigator shall notify the head of each study site with regard to the completion of the study in writing and submit a summary report on the study results. After receipt of notification from the study site on completion of the study, the sponsor shall report it to the regulatory authorities in compliance with the relevant standard operating procedure.

#### 15 STUDY MANAGEMENT

##### 15.1 Protocol Compliance, Deviations, and Modifications

1. The investigator or subinvestigator should not implement any deviation from or changes in the protocol without prior written agreement with the sponsor and prior review and documented approval of the Institutional Review Board.
2. The investigator or subinvestigator should document any deviation from the protocol.
3. The investigator or subinvestigator may implement a deviation from or a change in the protocol for medically unavoidable reasons, such as to eliminate an immediate hazard(s) to subjects, without prior agreement from the sponsor and prior approval of the Institutional Review Board. The investigator shall, as promptly as possible, submit a report on the details and reasons for the deviation or change, and, if amendment of the protocol is deemed necessary, a draft of the amended protocol to the sponsor, the head of the study site, and the Institutional Review Board, and shall obtain the approval of the Institutional Review Board for the draft of the amended protocol. The investigator shall then obtain the approval of the head of the study site, and the agreement of the sponsor via the head of the study site, in writing, regarding the amendment of the protocol.
4. The investigator should promptly provide written reports to the sponsor, the head of the study site, and the Institutional Review Board regarding any change that may significantly affect the conduct of the study and/or increase risks to subjects.

##### 15.2 Protocol Amendment

1. If amending the protocol, the sponsor should provide the investigator with the protocol amendment, the latest Investigator's Brochure, and other necessary documents and information.
2. The sponsor should provide ample time for the investigator to fully review the documents and information, such as the protocol amendment described above, and to discuss the documents and information with the sponsor.

3. After discussion with the investigator, the sponsor should agree with the investigator on the contents of the protocol amendment and the adherence to the protocol amendment. To confirm this agreement, the sponsor and the investigator should write their name and affix their name seal on or sign, and date the protocol or an alternative document. The same rule shall apply when the protocol is amended under the direction of the head of the study site based on the opinion of the Institutional Review Board.
4. The sponsor should promptly submit the protocol amendment to the Institutional Review Board.

##### 15.3 Amendments of the Investigator's Brochure

The sponsor shall amend the Investigator's Brochure if necessary, for instance, when new information is obtained.

##### 15.4 Compensation for Health Injury

###### 15.4.1 Principle of Compensation

1) The sponsor shall compensate for any health injury occurring in a study subject in accordance with the clinical trial compensation regulations prepared by the sponsor, at the request of the patient.
2) The sponsor shall prepare a summary of the clinical trial compensation regulations (hereinafter referred to as compensation summary) to make the regulations more understandable to subjects. The sponsor shall instruct the investigator or designee to specify in the Written Information, which will be used by the investigator or designee to provide an explanation to subjects, that any health injury will be compensated for as specified in the clinical trial compensation regulations and the protocol. Of note, a compensation summary will be provided by the investigator or designee to the subjects together with the Informed Consent Form and Written Information document.
3) Subject s may exercise the right to seek compensation from the sponsor, the study site, or a third party even after being compensated based on the clinical trial compensation regulations.
4) The same compensation shall be provided in a clinical trial using the same protocol in Japan.

###### 15.4.2 Coverage

1) A health injury for which a causal relationship to a study drug or protocol-specified procedures cannot be reasonably denied may be compensated for. This judgment must not be arbitrary.
   In assessing the causal relationship to a study drug the GCP guidance concerning Article 2 of the Ministerial Ordinance on GCP, which stipulates that an adverse event may be assessed as an

adverse drug reaction if a causal relationship between a medicinal product and an adverse event is at least a reasonable possibility, i.e., the relationship cannot be ruled out, can be referred to. The sponsor shall assess the causal relationship reasonably based on the case-by-case assessment criteria described in the GCP guidance as well as data accumulated up to the time.

2) Any health injury attributed to the sponsor, the study site, or a third party will not be compensated for. A health injury that has not been definitely attributed to the sponsor, the study site, or a third party within a reasonable period from a request for compensation may be compensated for.
3) A failure to provide expected drug effects or other benefits will not be compensated for.
4) A failure to provide treatment benefit to subjects receiving placebo will not be compensated for.
5) Any health injury resulting from the subject's intentional action will not be compensated for.
6) If a health injury results from the subject's gross negligence, compensation may be reduced or may not be paid.

### 16 DATA ENTRY INTO ELECTRONIC CASE REPORT FORMS (eCRFs)

#### 16.1 eCRF

The investigator or designee should enter data from each subject who has provided written informed consent in the eCRFs promptly after the completion of each assessment as specified in the protocol. For subjects who have provided informed consent but have never been randomized (screen failure), patient identification code, date of informed consent, date of withdrawal, age, sex, race, and reason for withdrawal (including an inclusion criterion that is not met or an exclusion criterion that is met, if appropriate) should be promptly entered in the eCRF. If a study collaborator enters the data on behalf of the investigator, his/her duty will be limited to the transcription of data from the source documents. The investigator shall assure, by checking the eCRF, that all data entered (including audit trail and contents for query) are accurate and complete, and provide an electronic signature in the eCRF. Since an electronic signature is legally equal to a handwritten signature, the contents endorsed with an electronic signature cannot be disaffirmed later. The procedures for entering, changing, or correcting data, providing an electronic signature in the eCRFs, and other relevant procedures should follow separately specified procedures.

#### 16.2 Inconsistency between eCRF and Source Document Such as Medical Record

The investigator or subinvestigator shall leave a record of confirmation within the source documents in the case of any inconsistencies in the description in the eCRF from source documents such as the medical record. If it is difficult to leave a record within the source documents, the investigator or subinvestigator shall prepare a "Report on Inconsistency from Source Document." The investigator shall submit the prepared "Report on Inconsistency from Source Document" to the sponsor and keep a copy of the report.

### 17 DIRECT ACCESS TO SOURCE DOCUMENTS

The investigator or the study site shall permit representatives of the sponsor (monitors and auditors), the Institutional Review Board, and regulatory authorities to conduct inspections of or have direct access to source documents for the purposes of investigation and confirmation that records are retained as required.

In addition, data that shall be directly entered in eCRFs and regarded as source documents are described below.

1. Eligibility of a study patient

2. Reasons for (concomitant) use of drugs and therapies, and reasons for changes if any
3. Presence/absence of abnormal changes
4. Severity and seriousness of adverse events, and causal relationship with the study drug
5. Reasons for study discontinuation for individual subjects and the subsequent courses
6. Comments

### 18 QUALITY CONTROL AND QUALITY ASSURANCE OF THE STUDY

#### 18.1 Sponsor

The quality control and quality assurance for the study should be implemented in accordance with the Ono Pharmaceutical Co., Ltd. standard operating procedures for the conduction of clinical studies. Prior to the initiation of the study, the sponsor will perform risk assessment using the Risk Assessment and Categorization Tool (RACT) to prepare a monitoring plan after reviewing the risk-based monitoring, and perform monitoring according to the monitoring plan.

#### 18.2 Study Sites

The quality control and quality assurance for the study shall be implemented in accordance with operating procedures for the conduction of clinical studies specified by the head of each study site.

#### 18.3 Contract Research Organization

The quality control and quality assurance for the study shall be implemented in accordance with operating procedures for the conduction of clinical studies specified by the contract research organization.

### 19 PROCEDURE FOR MONITORING SUBJECT COMPLIANCE WITH TREATMENT AND OTHER AGREEMENTS

#### 19.1 Investigator or Subinvestigator

The investigator or subinvestigator shall confirm whether the study drug is administered appropriately and whether blood and urine are collected appropriately at the study site.

##### 19.2 Drug Accountability Manager

The drug accountability manager shall check whether the study drugs are prescribed and dispensed appropriately at the study site in accordance with prescriptions (or source data) and the study drug accountability log.

##### 19.3 Sponsor (Monitor)

The sponsor (monitors) shall confirm that the study is conducted appropriately at the study site in accordance with the approved protocol.

#### 20 ARCHIVING OF ESSENTIAL DOCUMENTS AND RECORDS

##### 20.1 Sponsor

The sponsor shall archive documents and records related to the study in a suitable manner in accordance with the Ono Pharmaceutical Co., Ltd. standard operating procedures for the conduction of clinical studies until the day specified in 1 to 4 below, whichever occurs the latest:

1. The day on which 5 years have elapsed after the date of approval (in the case of termination of the development, the day on which 3 years have elapsed after the decision of development termination).

   However, for medicinal products that must be reexamined after approval in compliance with the PMD Act and for which the reexamination is to complete later than 5 years after approval, essential documents should be retained until the day of completion of the re-examination.
2. The day on which 3 years have elapsed since premature termination or completion of the study.

   If study-related documents or records are no longer required to be archived, the sponsor shall inform the head of the study site and the founder of the Institutional Review Board via the head of the study site that the study-related records are no longer required to be archived.
3. Until 2 years after the date of the final approval of the study drug in the ICH regions AND until the study drug is no longer in the process of evaluation for marketing approval or pending for marketing approval application in the ICH regions
4. Until 2 years after the date of the official discontinuation of the clinical development of the study drug in the ICH regions (the date of notification of the clinical development discontinuation to the regulatory authority of the last country in which discontinuation procedures were performed)

##### 20.2 Study Sites

Essential records and documents required to be preserved by the head of the study site or the founder of the Institutional Review Board shall be appropriately archived by an archive manager who is designated by the head of the study site. The essential documents and records will be retained until the day specified in 1 to 4 below, whichever occurs latest. If retention of these documents at the study site for the required period of time is difficult or if the sponsor requests that these documents be retained for a longer period of time, the study site should discuss the appropriate handling of the situation with the sponsor.

1. Date of approval of the study drug (or day on which 3 years have elapsed since the date of decision to discontinue development, if development is discontinued)
2. Date on which 3 years have elapsed since premature termination or completion of the study.
3. Until 2 years after the date of the final approval of the study drug in the ICH regions AND until the study drug is no longer in the process of evaluation for marketing approval or pending for marketing approval application in the ICH regions
4. Until 2 years after the date of the official discontinuation of the clinical development of the study drug in the ICH regions (the date of notification of the clinical development discontinuation to the regulatory authority of the last country in which discontinuation procedures were performed)

If study-related documents or records are no longer required to be archived, the sponsor shall inform the head of the study site and the founder of the Institutional Review Board via the head of the study site that the study-related records are no longer required to be archived. The head of the study site and the founder of the Institutional Review Board shall be responsible for archiving document of study-related records until informed by the sponsor that archiving is no longer required.

##### 20.3 Investigator

The investigator shall retain essential documents related to the conduction of the study as directed by the head of the study site.

#### 21 PAYMENT OF MONEY AND INSURANCE

##### 21.1 Payment of Money

The sponsor should discuss financing separately with each study site.

##### 21.2 Insurance

The sponsor should take measures such as securing insurance to ensure that the cost of treatment for study-related health injuries occurring in subjects or other losses will be compensated.

##### 21.3 Reduction of Burden on Subjects

The sponsor should discuss decisions on measures to be taken to reduce subject burden associated with participation in the study with each study site separately.

#### 22 PUBLICATION OF STUDY RESULTS

Prior confirmation by the sponsor is necessary before the results of the study conducted in accordance with this protocol may be published. The sponsor should decide matters related to the publication of study results through discussion with the coordinating investigator.

**Appendix 1 Ordinal Scale** [20)]

| Patient State | Descriptor | Score |
|---|---|---|
| ***Uninfected*** | No clinical or virological evidence of infection | 0 |
| ***Ambulatory*** | No limitation of activities | 1 |
| | Limitation of activities | 2 |
| ***Hospitalized Mild disease*** | Hospitalized, no oxygen therapy | 3 |
| | Oxygen by mask or nasal prongs | 4 |
| ***Hospitalized Severe Disease*** | Non-invasive ventilation or high-flow oxygen | 5 |
| | Intubation and mechanical ventilation | 6 |
| | Ventilation + additional organ support – pressors, RRT, ECMO | 7 |
| ***Dead*** | Death | 8 |

#### Appendix 2 Severity of Adverse Events

Source: The Standards for Classification of Serious Adverse Drug Reactions

Notification of the Safety Division Chief, PAB, Ministry of Health and Welfare (dated 29 June 1992, No. 80, SD-PAB)

1. These standards are to categorize the seriousness of adverse reactions into the following 3 Grades of 1 to 3:

   Grade 1: Considered to be a mild adverse reaction

   Grade 2: Not a serious adverse reaction but not a mild adverse reaction

   Grade 3: Considered to be a serious adverse reaction Specifically, depending on the patient's physical constitution and the condition at the time of onset, the event may result in death or persistent dysfunction on the level of disturbing the activities of daily living.

2. While these standards have been prepared as specific and easy criteria on judging the seriousness of adverse reactions, it should be considered that the seriousness of individual adverse reactions should be comprehensively assessed not only by the class of adverse reaction symptoms but also by considering the patient's general condition, status of underlying disease/concurrent diseases, and outcome, etc.

3. Although these standards were not prepared for interpretation of the reportability range for adverse reaction cases in compliance with Article 69 of the law (hereinafter, "case to be reported under Article 69"), an adverse reaction case for which Grade 3 of these standards is applicable roughly applies to a "case that may result in death or disability," which is stated in the Ordinance Article No. 62-2-1-1 among cases to be reported under Article 69. Thus, it is recommended to utilize these standards as criteria for judging whether a case qualifies as a case to be reported under Article 69.

4. Even if a case is an adverse reaction that does not qualify as a case to be reported under Article 69, to assure the safety measures from the viewpoint of healthcare are carried out, the actions to be taken for the adverse reactions on the levels described below are as follows:

(i) An adverse reaction that is suspected of being Grade 1 and not described in Precautions:

Such cases should be periodically accumulated and reported in compliance with "Amendment of reporting form for adverse reactions of pharmaceutical products"-3 (Reporting of unexpected and mild adverse reactions) in No. 24, SD-PAB dated on 26 February 1992.

(ii) An adverse reaction that is suspected of being Grade 2 and not described in Precautions:

Such cases should be reported immediately.

(iii)An adverse reaction that is suspected of being Grade 3:

Such cases should be reported immediately.

**The Standards for Classification of Serious Adverse Drug Reactions**

**Liver**

The seriousness of liver disorder is graded according to laboratory test data, symptoms, etc. shown in the table below, in principle. In addition, if liver disorder is suspected according to clinical symptoms such as having general malaise, anorexia, nausea, pyrexia and rash, GOT, GPT, etc. of the patient should be checked and also categorized according to the table below. In addition, if a result of liver biopsy has been obtained, this should be considered for the assessment.

| Grade for adverse reaction | Grade 1 | Grade 2 | Grade 3 |
|---|---|---|---|
| Total bilirubin (mg/dL) | ≥1.6 to <3.0 | ≥3.0 to <10 | ≥10 |
| GOT, GPT (U) | ≥1.25×N to <2.5×N<br>≥50 to <100 | ≥2.5×N to <12×N<br>≥100 to <500 | ≥12×N<br>≥500 |
| Al-P | ≥1.25×N to <2.5×N | ≥2.5×N to <5×N | ≥5×N |
| γ-GTP | ≥1.5×N | — | — |
| LDH | ≥1.5×N | — | — |
| PT | — | — | ≤40% |
| Symptoms etc. | — | Jaundice<br>Hepatomegaly<br>Hypochondrium pain right<br>Hepatic steatosis | Hepatic failure symptoms, e.g., bleeding tendency, consciousness disturbed (hepatitis fulminant)<br>Hepatic cirrhosis<br>Liver tumor<br>Jaundice prolonged for ≥6 months |

N: upper limit of normal range for each site

**Kidney**

The seriousness of renal disorder is graded according to laboratory test data, symptoms, etc. shown in the table below, in principle. In addition, if renal disorder is suspected according to clinical symptoms such as general malaise, anorexia, nausea, edema, hypertension, headache dull or urinary findings, BUN, creatinine, etc. of the patient should be checked and also categorized according to the table below. In addition, if a result of renal biopsy has been obtained, this should be considered for the assessment.

| Grade for adverse reaction | Grade 1 | Grade 2 | Grade 3 |
|---|---|---|---|
| BUN (mg/dL) | >1×N to <25 | ≥25 to <40 | ≥40 |
| Creatinine (mg/dL) | >1×N to <2 | ≥2 to <4 | ≥4 |
| Proteinuria | 1+ | 2+ to 3+ | >3+ |
| Hematuria | Microscopic | Macroscopic | Macroscopic, clotting blood |
| Urinary output | — | ≤500 mL/24 hr<br>or oliguria<br>Polyuria [Note] | ≤100 mL/24 hr<br>or anuria |
| Serum potassium level (mEq/L) | — | ≥5.0 to <5.5 | ≥5.5 |
| Other symptoms, etc. | — | — | Nephrotic syndrome<br>Renal failure acute (nephritis interstitial, tubular necrosis, renal necrosis, renal papillary necrosis, renal cortical necrosis)<br>Renal failure chronic (nephritis interstitial tubular necrosis, renal necrosis, renal papillary necrosis, renal cortical necrosis)<br>Uraemia<br>Hydronephrosis |

N: upper limit of normal range for each site
Note) Renal diabetes insipidus.

**Blood**

The seriousness of blood disorder is graded according to laboratory test data, symptoms, etc. shown in the table below, in principle.

| Grade for adverse reaction | Grade 1 | Grade 2 | Grade 3 |
|---|---|---|---|
| Red blood cells | <3500000 to ≥3000000 | <3000000 to ≥2500000 | <2500000 |
| Hb (g/dL) | <11 to ≥9.5 | <9.5 to ≥8 | <8 |
| White blood cells | <4000 to ≥3000 | <3000 to ≥2000 | <2000 |
| Granulocytes | <2000 to ≥1500 | <1500 to ≥1000 | <1000 |
| Platelets | <100000 to ≥75000 | <75000 to ≥50000 | <50000 |
| Bleeding tendency | Mild bleeding (haemorrhage subcutaneous) | Moderate bleeding (mucosal haemorrhage) [Note 1] | Severe bleeding (intraorgan bleeding) [Note 2] |
| Other symptoms, etc. | — | — | Pancytopenia<br>(e.g., aplastic anaemia)<br>Aplasia pure red cell<br>Agranulocytosis |

Note 1) Mucosal haemorrhage — gingival bleeding, epistaxis

Note 2) Intraorgan bleeding — haemorrhage intracranial, haemorrhage of digestive tract, pulmonary haemorrhage, renal haemorrhage, genital haemorrhage, muscle haemorrhage, intraarticular haemorrhage

**Hypersensitive symptoms**

The seriousness of hypersensitive symptoms is graded according to symptoms, etc. shown in the table below, in principle.

| Grade for adverse reaction | | Grade 1 | Grade 2 | Grade 3 |
|---|---|---|---|---|
| Skin symptoms | | Local rash<br>(e.g., local erythema/ rash papular)<br>Itching | Widespread rash<br>(e.g., general erythema, purpura, blister) | Oculomucocutaneous syndrome<br>Toxic epidermal necrolysis<br>Erythroderma (dermatitis exfoliative)<br>Weber-Christian syndrome<br>SLE-like symptoms [Note 1]<br>Scleroderma<br>Pemphigus-like lesion |
| | | (e.g., photosensitivity, fixed eruption, erosion/ulcer, pigmentation) | | |
| General symptoms | Pyrexia | Pyrexia [Note 2, Note 3] | | — |
| | Allergy | — | — | Shock<br>Anaphylactic symptoms [Note 4] |
| | | Angioedema (except for laryngeal part e.g., face oedema, eyelid oedema) [Note 3] | | Angioedema (laryngeal oedema) |
| | Vasculitis | — | Hypersensitivity vasculitis [Note 5] | |
| Local symptoms | | Arthralgia [Note 3]<br>Swollen lymph nodes [Note 3] | | —<br>— |

Note 1) As for SLE-like symptoms, general symptoms should also be considered.

Note 2) Pyrexia indicates so-called drug fever.

Note 3) The current physician, etc. should judge Grade 1 or Grade 2.

Note 4) Anaphylactic symptoms are acute and serious dyspnoea without hypotension, which are considered to be general and serious symptoms or allergic reactions associated with multiple symptoms among dyspnoea, generalized flushing, angioedema (e.g., face oedema, laryngeal oedema), and urticaria.

Note 5) The current physician, etc. should judge Grade 2 or Grade 3.

**Respiratory**

The seriousness of respiratory system disorder is graded according to laboratory test data, symptoms, etc. shown in the table below, in principle.

| Grade for adverse reaction | | Grade 1 | Grade 2 | Grade 3 |
|---|---|---|---|---|
| Respiratory condition | Dyspnoea | Shortness of breath<br>HJ classification Grade II [Note] | Dyspnoea on exertion<br>HJ classification Grade III to IV [Note 1] | Dyspnoea at rest<br>HJ classification Grade V [Note 1] |
| | Respiratory rhythm disorder | — | Transient hyperventilation<br><br>Sleep apnoea without clinical symptoms and hypoxaemia [Note 2] | Respiratory arrest (apnoea)<br>Respiratory depression (hypoventilation, carbon dioxide narcosis)<br>Persistent hyperventilation (respiratory distress, hyperpnoea)<br>Cheyne-Stokes respiration<br>Sleep apnoea with clinical symptoms and hypoxaemia [Note 2] |
| Partial pressure of oxygen in arterial blood $PaO_2$ (mmHg) | | <70 to ≥60 | <60 to ≥50 | <50<br>Decrease by ≥20 from baseline |
| Partial pressure of carbon dioxide in arterial blood $PaCO_2$ (mmHg) | | — | — | ≥50 (hypoventilation)<br>≤30 (hyperventilation) |
| % vital capacity<br>FEV1% | | —<br>— | <70% to ≥50%<br><70% to ≥50% | <50%<br><50% |
| Chest X-ray findings | Infiltrative shadow | — | <1/3 of one lung [Note 3] | ≥1/3 of one lung [Note 3] |
| | Interstitial shadow | — | — | Appearance of erosive interstitial shadow |
| | Pleural effusion | — | <1/3 of one lung [Note 3] | ≥1/3 of one lung [Note 3] |
| Asthma attack | | — | Wheezing,<br>mild attack [Note 4] | Moderate attack, severe attack [Note 4]<br>Asthmaticus Status |
| Haemoptysis | | — | Sputum bloody | Haemoptysis |
| Other symptoms etc. | | Hiccups<br>Yawning<br>Hoarseness<br>Sneezing<br>Nasal congestion/nasal cavity strange sensation<br>Cough<br>Sputum increased/ sputum expectoration difficult<br>Pharyngolaryngeal discomfort<br>Pharyngeal pain<br>Respiratory tract irritation<br>Chest pressure sensation | — | ARDS (adult respiratory distress syndrome)<br>Interstitial pneumonia<br>PIE syndrome<br>Pulmonary fibrosis<br>Hypersensitivity pneumonia<br>Pulmonary oedema<br>Pulmonary embolism<br>Pulmonary vasculitis<br>Glossoptosis<br>Laryngospasm<br>Oedema glottis<br>Pulmonary hypertension [Note] |
| | | Chest pain, feeling of pharyngeal stenosis (laryngopharyngeal paraesthesia) [Note 5] | | |

Note 1) HJ classification for dyspnoea

Grade I Capable of walking or climbing up hills and stairs at the same speed as people of the same age. Shortness of breath (-)

Grade II Capable of walking, but incapable of climbing up hills and stairs at the same speed as people of the same age.

Grade III Capable of walking more than 1600 m but not at the same speed as people of the same age.

Grade IV Incapable of walking around 45 m without a break

Grade V Have shortness of breath on changing clothes and conversation, incapable of going out because of shortness of breath

Note 2) Sleep apnoea is defined as around 5 times of ≥10 seconds respiratory arrest per hour while asleep. Clinical symptoms for this status are headache, sexual impotence, hypertension, cardiac failure, daytime narcoleptic tendency, etc.

Note 3) If findings indicating infiltrative shadow and pleural effusion have not been observed, classification should be Grade 3.

Note 4) Category of asthma attack is generally according to the following:

Mild attack Painful but capable of lying down. Normal conversation, normal mobility.

Moderate attack Painful and incapable of lying down. Slight difficulty in having conversation, much difficulty in mobility.

Severe attack Painful and incapable of moving. Incapable of having conversation and mobility.

Meanwhile, in the case of children, "The Standards of Severity Assessment Committee of Pediatric Allergy Research Group" with regard to grading of pediatric asthma attack (see next section) should be referred to.

Note 5) The current physician, etc. should judge Grade 1 or Grade 2.

Note 6) The seriousness classification of pulmonary capillary pressure under "Cardiovascular" should also be referred to for the level of pulmonary arterial pressure.

(Reference)

The Standards of Severity Assessment Committee of Pediatric Allergy Research Group

Level of pediatric asthma attack

| | Respiratory status | Life status | | | |
|---|---|---|---|---|---|
| | | Playing | Sleeping | Mood (conversation) | Eating |
| Mild attack | Having mild wheezing without dyspnoea, sometimes accompanied by mild retractive breathing | Normal | Normal | Normal Normal conversation | Normal |
| Moderate attack | Having obvious wheezing and retractive breathing, with dyspnoea | Slightly difficult | Awake sometimes | Slightly poor Capable of answering when spoken to | Slightly poor |
| Severe attack | Having marked wheezing, dyspnoea and orthopnoea, and sometimes having cyanosis. | Incapable or nearly incapable | Incapable or nearly incapable | Poor Incapable of answering when spoken to | Poor or nearly poor |

1. The level of attack shall be assessed mainly according to the respiratory status, and other items are reference information.
2. Breathing sounds decreased and consciousness disturbed (e.g., agitation, depressed consciousness, response decreased due to pain) are signs for risks.

**Gastrointestinal**

The seriousness of gastrointestinal disorder is graded according to laboratory test data, symptoms, etc. shown in the table below, in principle.

<table>
<tr><th>Grade for adverse reaction</th><th>Grade 1</th><th>Grade 2</th><th>Grade 3</th></tr>
<tr><td>Nausea, vomiting</td><td colspan="2">Nausea (feeling queasy)<br>Vomiting [Note 1]</td><td>—</td></tr>
<tr><td>Diarrhoea</td><td>Loose stools, mushy stool</td><td>Watery stool for which Grade 3 does not apply</td><td>Watery stool accompanied by dehydration and electrolyte abnormality</td></tr>
<tr><td>Haemorrhage of digestive tract</td><td>Occult blood (+)</td><td>Bloody stool, haematemesis, melena not accompanied by shock and hemoglobin decreased (≤8.0 g/dL)</td><td>Bloody stool, haematemesis, melena accompanied by shock or hemoglobin decreased (≤8.0 g/dL)</td></tr>
<tr><td rowspan="2">Abnormality in mouth</td><td>Subjective oral discomfort<br>(e.g.) lip dry feeling, oral discomfort, oral numbness, oral bitterness, numbness of tongue, abnormal sensation of tongue</td><td>Ulcerative stomatitis</td><td rowspan="2">—</td></tr>
<tr><td colspan="2">Abnormality in mouth accompanied by objective inflammation, etc. [Note 1]<br>(e.g.) angular stomatitis, cheilitis (lip vesicle), stomatitis (oral rough, gingival pain), glossitis (tongue eruption, tongue rough, glossodynia), tongue coated, tongue black, gingival hypertrophy</td></tr>
<tr><td>Abnormality in oesophagus</td><td>Subjective oesophageal discomfort<br>(e.g.) loading feeling, esophageal closed sensation</td><td colspan="2">Abnormality in esophagus accompanied by objective inflammation, ulcer, etc. [Note 2]<br>(e.g.) oesophagitis, oesophageal ulcer</td></tr>
<tr><td>Dysphagia</td><td>—</td><td>Swallowing difficult</td><td>Aphagia</td></tr>
<tr><td>Abnormality in stomach and intestines</td><td>Subjective gastrointestinal discomfort<br>(e.g.) heartburn, dyspepsia, stomach feeling heavy, gastric discomfort, abdominal discomfort, borborygmus, anorexia</td><td>—</td><td>—</td></tr>
<tr><td>Pain</td><td>Gastralgia or abdominal pain for which Grade 2 does not apply and of tolerable level or a level that does not require treatment</td><td>Colicky pain (stomach cramps, abdominal cramps, bowel cramps)</td><td>—</td></tr>
<tr><td rowspan="3">Inflammation</td><td colspan="3">Gastritis, enterocolitis, colitis [Note 3]</td></tr>
<tr><td colspan="2">Proctitis (rectal mucosal edema, rectal mucosal stimulation) [Note 1]</td><td>—</td></tr>
<tr><td>—</td><td colspan="2">Hemorrhagic colitis, pseudomembranous colitis [Note 2]</td></tr>
<tr><td>Ulcer</td><td>Erosion</td><td>Gastric ulcer, duodenal ulcer, haemorrhagic ulcer, small intestine ulcer, large intestinal ulcer [Note 2]</td><td>Gastrointestinal perforation</td></tr>
<tr><td>Intestinal paralysis</td><td colspan="2">Constipation [Note 1]</td><td>Ileus paralytic</td></tr>
<tr><td rowspan="2">Abnormality in anus</td><td>Subjective anal discomfort<br>(e.g.) anal pain, anal discomfort, strange feeling in the anus of, anal itching</td><td>—</td><td rowspan="2">—</td></tr>
<tr><td colspan="2">Anal abnormality accompanied by objective inflammation [Note 1]<br>(e.g.) periproctitis (anal sore, anal erosion), haemorrhoidal haemorrhage, prolapsed haemorrhoid</td></tr>
<tr><td>Pancreatic disorder</td><td>Only with abnormal amylase level</td><td>Pancreatitis for which Grade 3 does not apply</td><td>Pancreatic necrosis, pancreatitis haemorrhagic</td></tr>
<tr><td>Other symptoms, etc.</td><td>Hiccups, thirst (mouth feeling dry), belching (eructation), pigmentation of colon mucosa, flatulence, flatus, sulfurous odor, frequent bowel movements (desire to defecate, defecation urgency, tenesmus)</td><td>—</td><td>—</td></tr>
</table>

| Grade for adverse reaction | Grade 1 | Grade 2 | Grade 3 |
|---|---|---|---|
| | Sialoadenitis, faecal incontinence [Note 1] | | |

Note 1) The current physician, etc. should judge Grade 1 or Grade 2.

Note 2) It should be judged to categorize into Grade 2 or Grade 3 according to the level of concomitant clinical symptoms such as diarrhoea, haemorrhage of digestive tract, and dysphagia.

Note 3) Expressions of gastritis, enterocolitis, and colitis are often used by summarizing clinical symptoms such as vomiting, gastralgia, abdominal pain, and diarrhoea, regardless of having objective inflammation. The seriousness of these is graded according to the level of clinical symptoms such as vomiting.

**Cardiovascular**

The seriousness of cardiovascular disorder is graded according to laboratory test data, symptoms, etc. shown in the table below, in principle.

| **Grade for adverse reaction** | | | **Grade 1** | **Grade 2** | **Grade 3** |
|---|---|---|---|---|---|
| Abnormal blood pressure | Decrease | Systolic blood pressure (mmHg) | — | <90 to ≥80 | <80 |
| | | Symptoms | Dizziness on standing up, dizziness upon standing, orthostatic hypotension | | Impalpable pulse |
| | Increase | | Blood pressure increased (abnormal increased blood pressure, rapidly increased blood pressure), hypertension | | — |
| Circulatory disorder | | | — | — | Shock<br>Cyanosis<br>Peripheral circulatory failure |
| Heart rate (/min) | Tachycardia | | — | ≥110 to <130 | ≥130 |
| | Bradycardia | | — | <50 to ≥40 | <40 |
| Arrhythmia | | | Palpitations, arrhythmia (ECG untested) | | |
| | | | Supraventricular extrasystoles | Supraventricular tachycardia | |
| | | | Ventricular extrasystoles (single) | Ventricular extrasystoles (couplet)<br>Bigeminal pulse | Ventricular extrasystoles (multifocal) (≥ triplet)<br>Ventricular tachycardia (≥ sextuplet)<br>Ventricular fibrillation<br>Torsades de pointes |
| | | | | Atrial fibrillation (including paroxysmal episodes)<br>Atrial flutter | |
| | | | | Tachycardia paroxysmal | |
| | | | Grade 1 atrioventricular block (Atrioventricular conduction time prolonged) | Grade 2 atrioventricular block<br>Atrioventricular dissociation<br>Sinus arrest<br>Bundle branch block (intraventricular block) (intraventricular conduction defect)<br>Nodal rhythm<br>Ventricular rhythm | Grade 3 atrioventricular block (complete atrioventricular block)<br>Cardiac arrest (heart stroke arrest)<br>Adams-Stokes syndrome |
| Abnormal ECG findings | | | P wave disappearance<br>PR/PQ prolongation | ST increased<br>ST decreased<br>T wave inverted<br>T wave flattening<br>U wave appearance<br>QT prolonged<br>QRS widened | — |
| Heart failure-like symptoms | | | — | Edema (general/peripheral) | Cardiac failure (cardiac failure congestive)<br>Right heart failure<br>Left heart failure (cardiac asthma)<br>Cardiac failure acute<br>Cardiomegaly (cardiothoracic index increased) |
| Reference | Myocardial contractile force | | 60%≥ left ventricular ejection fraction >50% | 50%≥ left ventricular ejection fraction >40% | 40% ≥ left ventricular ejection fraction |
| | Cardiac output (cardiac index) | | — | 2.5 L/min/m$^2$ ≥ | 2.2 L/min/m$^2$ ≥ |
| | Pulmonary capillary pressure (pulmonary arterial systolic pressure) (mmHg) | | ≥20 to <30 | ≥30 to <40 | ≥40 |

| Grade for adverse reaction | Grade 1 | Grade 2 | Grade 3 |
|---|---|---|---|
| Dyspnoea (see seriousness classification for “Respiratory”) | Shortness of breath<br>HJ classification Grade II | Dyspnoea on exertion<br>HJ classification Grade III to IV | Dyspnoea at rest<br>HJ classification Grade V |
| Ischemic heart disease-like symptoms | Chest discomfort<br>Chest distressed feeling<br>Chest pressure sensation | — | Angina pectoris aggravated<br>Anginal attack (induced anginal attack)<br>Myocardial infarction (coronary artery thrombosis)<br>Myocardial necrosis |
| | Chest pain, anginal pain (angina-like pain), myocardial ischaemia, coronary insufficiency [Note] | | |

Note) The current physician, etc. should judge Grade 1 or Grade 2.

<table>
<tr><th>Grade for adverse reaction</th><th>Grade 1</th><th>Grade 2</th><th>Grade 3</th></tr>
<tr><td rowspan="2">Myocardial/pericardial/<br>endocardial disorder</td><td>—</td><td>Pericarditis<br>Pericardial exudate retention<br>Endocarditis</td><td>Myocarditis<br>Myocardial fibrosis</td></tr>
<tr><td colspan="2">Myocardial disorder [Note]</td><td></td></tr>
<tr><td rowspan="2">Vascular disorder</td><td>Vascular pain</td><td>Angiospasm<br>Claudicatio intermittens<br>Arteriosclerosis</td><td rowspan="2">Gangrene<br>Vasculitis<br>Thrombophlebitis<br>Thrombosis<br>Arterial thrombus/venous thrombus<br>Thromboembolism<br>Pulmonary embolism (infarction)<br>Cerebral embolism (infarction)<br>Mesenteric embolism</td></tr>
<tr><td colspan="2">Raynaud's-like syndrome [Note]<br>(without gangrene)</td></tr>
<tr><td>Other symptoms</td><td>Flushed face (hot flush)<br>Feeling hot<br>Burning sensation<br>Hot flush</td><td>—</td><td>—</td></tr>
</table>

Note) The current physician, etc. should judge Grade 1 or Grade 2.

**Psychoneurotic**

The seriousness of psychoneurotic disorder is graded according to the status, etc. shown in the table below, in principle, in consideration of the status such as subjective/objective, controllable/uncontrollable by others, support required/not required, transient/persistent, and reversible/irreversible.

| Grade for adverse reaction | | Grade 1 | Grade 2 | Grade 3 |
|---|---|---|---|---|
| Psychological activities and abnormal behavior | Mood elevation or unstable mood | Subjective mood elevation or unstable mood | Grade 1 status is observed objectively as well, accompanied by abnormal behavior | Events of Grade 2 with severe and uncontrollable symptoms |
| | | (e.g.) unstable temper, mood swings, labile affect, nervousness, hypersensitivity, irritable feeling, bad mood, anxiety (feeling), feeling irritated, talkativeness, hyperthimic, mania, euphoria (euphoric feeling) | (e.g.) manic depression/manic state, change to manic state, aggression, agitation with stimulation, agitation, irritability, unrest, irritable hyperactivity, prowling, impulsive behavior, lack of suppression, affective incontinence | |
| | | Insomnia (sleep disorder) | | |
| | Mood/ motivation/ behavior decreased | Subjective mood and behavior decreased | Grade 1 status is observed objectively as well | Events of Grade 2 with severe and uncontrollable symptoms |
| | | (e.g.) hypobulia, dullness, lack of motivation, feeling lack of motivation, listless status, unselfish status, fuzzy head, mental dullness, dreamy state, mental concentration decreased, depression state, depression (status), feeling blue, melancholia | | (e.g.) suicidal ideation/attempt Depressive stupor |
| | Psychotic-like symptoms | — | Transient illusion/hallucination/deliria (e.g., delirium nocturnal) | Persistent illusion/hallucination/deliria, confusion, delusion |
| | Intellectual mental function disturbance | Subjective intellectual capacity decreased | Intellectual capacity decreased that is observed objectively | Events of Grade 2 with severe and persistent symptoms |
| | | (e.g.) memory loss, decreased memory/ capacity to register | (e.g.) anterograde amnesia, retrograde amnesia | (e.g.) dementia |
| Consciousness disorder | | Subjective consciousness disorder | Consciousness disorder that is observed objectively as well | Events of Grade 2 with severe and persistent symptoms |
| | | (e.g.) sleepiness, twilight state, arousal difficult, delayed awakening, feeling drunk, feeling of residual sleepiness, after sleeping, sedation, excessive sedation, nightmare, excessive dreaming | (e.g.) somnolence, lethargy, drowsy state, twilight state, consciousness clouding, transient loss of consciousness, syncope, orientation disturbed, loss of orientation | (e.g.) coma, persistent loss of consciousness |
| Movement disorder | Coordinated movement | Subjective coordinated movement disorder | Coordinated movement disorder that is observed objectively as well | Events of Grade 2 with severe symptoms that disturb the activities of daily living significantly and require support |
| | | (e.g.) wobble, dizziness, vertigo, wooziness (feeling) | (e.g.) ataxia, coordinated movement disorder | |
| | Walking | — | Walking disorder that is observed objectively | Events of Grade 2 with severe symptoms that disturb the activities of daily living significantly and require support |
| | | | (e.g.) frozen gait, gait disturbance, difficulty in walking, atactic gait, abnormal gait | (e.g.) abasia |
| | Muscular force/ paralysis | — | Muscle weakness and disorder that is observed objectively | Events of Grade 2 with severe symptoms that disturb the activities of daily living significantly and require support |

| | | | | |
|---|---|---|---|---|
| | | | (e.g.) hypotonia, muscle faintness, muscular weakness, paresis | (e.g.) facial paralysis, quadriplegia, hemiplegia, monoplegia |
| | Myalgic pain/ arthralgia | Events of tolerable level or a level that does not require treatment | Events with severe and persistent symptoms | — |
| | | (e.g.) arthralgia, myalgia, back pain, low back pain, nuchal pain, neck pain | | |

| Grade for adverse reaction | | Grade 1 | Grade 2 | Grade 3 |
|---|---|---|---|---|
| Movement disorder (continued) | Extrapyramidal symptoms: Involuntary movements | Transient and mild involuntary movements | Persistent involuntary movements that can be recognized as neural symptoms | Events of Grade 2 with severe symptoms that disturb the activities of daily living significantly and require support |
| | | (e.g.) transient tremor (tremor limb, tremor finger), tremble hands, tremble | (e.g.) rough or persistent tremor, involuntary movements around the mouth, mimic tic, protrusion tongue, mask-like facies, dyskinesia, hyperkinesia, akathisia, hyperkinesis, Parkinson's syndrome (those symptoms of Parkinson's syndrome, Parkinson's syndrome-like symptoms, aggravation of symptoms of Parkinson's syndrome) | |
| | Extrapyramidal symptoms: Muscle tightness | Subjective muscle tension abnormal | Severe level of muscle tension that can be recognized as neural symptoms | Events of Grade 2 with severe symptoms that disturb the activities of daily living significantly and require support |
| | | (e.g.) bradykinesia, slow movement, stiff shoulder, anteversion-anteflexion posture, stiffness feeling of legs | (e.g.) facial/perioral tension, hypertonia, rigidity, muscle rigidity, muscle ankylosis, muscle stiffness, muscle spasticity, ankylosis of neck (extremities), body stiffness | |
| | Language disorder | Subjective language disorder | Language disorder that is observed objectively as well | Events of Grade 2 with severe symptoms that disturb the activities of daily living significantly and require support |
| | | (e.g.) lisp (oral lisp), tongue movement disturbance | (e.g.) dyslalia, dysarthria | (e.g.) aphasia |
| | Eye movement disorder | — | Transient eye movement disorder | Events of Grade 2 with severe and persistent symptoms |
| | | | (e.g.) ocular deviation, oculogyric crisis, ocular lateral crisis, eyeballs raise upward, nystagmus, diplopia | |
| | Reflex | Reflex decreased | Morbid escalation of reflex<br>Disappearance of reflex | Appearance of morbid reflex |
| | | (e.g.) tendon reflex decreased,<br>reduced ability of reflex movement | | (e.g.) Babinski's reflex |
| Convulsions | | Subjective convulsions | Local convulsions | General convulsions |
| | | (e.g.) shakiness | (e.g.) twitching, muscle twitching, neck/facial twitching, extension of upper extremities, muscle cramp | (e.g.) generalized convulsion, epileptic seizure, epileptiform seizure, clonic convulsion, tonic convulsion, seizure, induction of convulsion, opisthotonos |
| Sensory function disorder | Hearing impaired | Subjective hearing impaired | Transient hearing impaired that is observed objectively | Irreversible hearing impaired |
| | | (e.g.) tinnitus, closed sensation of ears | (e.g.) hearing reduced, hypoacusis | (e.g.) irreversible hearing loss, deaf (completely incapable status of hearing) |
| | Visual disturbance | Subjective abnormal vision | Transient visual disturbance that is observed objectively | Irreversible visual disturbance |
| | | (e.g.) photophobia, feeling of vision decreased, feeling of visual flash, vision blurred, visual adjustment disorder | (e.g.) visual acuity reduced transiently,<br>transient color blindness | (e.g.) optic neuritis, blindness, visual field disturbance |

| | | | | |
|---|---|---|---|---|
| | Smell disorder | Transient smell disorder [Note] | | Irreversible smell disorder |
| | | (e.g.) dysosmia, strange smell sensation | | (e.g.) loss of smell |
| | Taste disturbance | Transient taste disturbance [Note] | | Irreversible taste disturbance |
| | | (e.g.) abnormal feeling of tongue, dysgeusia, hypogeusia | | (e.g.) loss of taste |
| | Sensation (feeling) disorder | Transient sensation (feeling) disorder [Note] | | Irreversible sensation (feeling) disorder |
| | | (e.g.) numbness of extremities, etc., numbness of tongue, perioral numbness, ear pain, sensation (feeling) change, sensation (feeling) decreased | | (e.g.) loss of sensation (feeling) |
| Peripheral nerve (nervous disorder) | | Transient neuralgia | Persistent neuralgia | Events of Grade 2 with severe symptoms that disturb the activities of daily living significantly and require support |
| | | | | (e.g.) Guillain-Barre syndrome, multiple neuritis, peripheral neuritis, myopathy |

Note) The current physician, etc. should judge Grade 1 or Grade 2.

| Grade for adverse reaction | Grade 1 | Grade 2 | Grade 3 |
|---|---|---|---|
| Dependency | — | Events with mild mental dependency that are observed with a dose-dependent tendency (tendency of resistance appeared) | Events with physically-dependency, withdrawal symptoms (abstinence symptoms) |
| Others | Yawning, cerebral anemia-like symptoms, floating feeling, unstable feeling, headache, heaviness of head (feeling), head pressure, feeling strange, abnormal physical sensation, feeling of fatigue, general malaise, feelings of weakness, discomfort, poor feeling | Swallowing difficult (reduced swallowing capability)<br>Salivation | Aphagia<br>Neuroleptic malignant syndrome<br>Hyperthermia malignant<br>Encephalopathy/leukoencephalopathy<br>Meningitis/meningitis-like symptoms<br>Cerebrovascular disorder (e.g., cerebral haemorrhage, cerebral infarction) |

Note) The current physician, etc. should judge Grade 1 or Grade 2.

**Metabolism/electrolyte abnormality**

The seriousness of metabolism/electrolyte abnormality is graded according to laboratory test data, symptoms, etc. shown in the table below, in principle.

| Grade for adverse reaction | | Grade 1 | Grade 2 | Grade 3 |
|---|---|---|---|---|
| Abnormal blood sugar (mg/dL) | Sugar blood level increased | Blood sugar 120 to 200<br>or<br>Fasting 120 to 140<br>After meals 160 to 200 | Blood sugar 201 to 300<br>or<br>Fasting 141 to 200<br>After meals 201 to 300 | Blood sugar ≥301 |
| | Symptoms | — | — | Diabetic coma |
| | Sugar blood level decreased | 69 to 60 | 59 to 51 | ≤50 |
| | Symptoms | — | Hypoglycemic symptoms such as dizziness, headache, feeling hungry, irritable feeling, significant sweating | Hypoglycemic coma, convulsions |
| Metabolic acidosis | Arterial blood pH | <7.35 to ≥7.20 | <7.20 to ≥7.15 | <7.15 |
| | Symptoms | — | — | Consciousness disturbed, blood pressure decreased, convulsions, respiratory disorder (Kussmaul type) |
| Metabolic alkalosis | Arterial blood pH | ≥7.46 to <7.50 | ≥7.50 to <7.60 | ≥7.60 |
| | Symptoms | — | — | Convulsions, tetany, hypertension, arrhythmia |
| Blood calcium abnormal (mg/dL) | Increase | ≥10.6 to <12.1 | ≥12.1 to <15.0 | ≥15.0 |
| | Symptoms | — | — | Consciousness disturbed |
| | Decrease | <8.5 to ≥8.0 | <8.0 to ≥6.5 | <6.5 |
| | Symptoms | — | — | Tetany, blood pressure decreased, arrhythmia, psychiatric symptom |
| Serum potassium abnormal (mEq/L) | Increase Note | ≥5.0 to <5.5 | ≥5.5 to <6.0 | ≥6.0 |
| | Symptoms | — | — | Arrhythmia, muscle paralysis |
| | Decrease | <3.5 to ≥3.1 | <3.1 to ≥2.5 | <2.5 |
| | Symptoms | — | — | Hyposthenia, muscle paralysis, arrhythmia |
| Serum sodium abnormal (mEq/L) | Increase | ≥150 to <155 | ≥155 to <160 | ≥160 |
| | Symptoms | — | — | Central nerve symptoms (consciousness disturbed, convulsions) |
| | Decrease | <135 to ≥125 | <125 to ≥115 | <115 |
| | Symptoms | — | — | Mental disorder, convulsions, consciousness disturbed, morbid reflex |

Note) Increase of serum potassium level associated with renal disorder is graded according to the seriousness classification for “Kidney.”

#### Signatures for the Protocol

Protocol Number: FOY-305-01

Version No.: Ver. 2

I agree to comply with the content of the protocol described above and all the references in the protocol described above.

Investigator

Day/Month/Year

Name of the institution ______________________________

Name of the department ______________________________

Investigator ______________________________

I have reviewed and approved the content of the protocol described above. I will assume the responsibility for managing and supervising the study in accordance with the protocol.

Study Representative

Day/Month/Year

Clinical Development Planning Division, Ono Pharmaceutical Co., Ltd.

______________________________

Background of Protocol (Protocol Number: FOY-305-01)

Version 1: Prepared on 28 August 2020

Version 2: Prepared on 20 November 2020
